# SARS-CoV-2 is associated with changes in brain structure in UK Biobank

**DOI:** 10.1101/2021.06.11.21258690

**Authors:** Gwenaëlle Douaud, Soojin Lee, Fidel Alfaro-Almagro, Christoph Arthofer, Chaoyue Wang, Paul McCarthy, Frederik Lange, Jesper L.R. Andersson, Ludovica Griffanti, Eugene Duff, Saad Jbabdi, Bernd Taschler, Peter Keating, Anderson M. Winkler, Rory Collins, Paul M. Matthews, Naomi Allen, Karla L. Miller, Thomas E. Nichols, Stephen M. Smith

**Author notes:** Correspondence to: Prof. Gwenaëlle Douaud, FMRIB Centre, Wellcome Centre for Integrative Neuroimaging, Nuffield Department of Clinical Neurosciences, John Radcliffe Hospital, Oxford OX3 9DU, UK.

## Abstract

There is strong evidence for brain-related abnormalities in COVID-19^1–13^. It remains unknown however whether the impact of SARS-CoV-2 infection can be detected in milder cases, and whether this can reveal possible mechanisms contributing to brain pathology. Here, we investigated brain changes in 785 UK Biobank participants (aged 51–81) imaged twice, including 401 cases who tested positive for infection with SARS-CoV-2 between their two scans, with 141 days on average separating their diagnosis and second scan, and 384 controls. The availability of pre-infection imaging data reduces the likelihood of pre-existing risk factors being misinterpreted as disease effects. We identified significant longitudinal effects when comparing the two groups, including: (i) greater reduction in grey matter thickness and tissue-contrast in the orbitofrontal cortex and parahippocampal gyrus, (ii) greater changes in markers of tissue damage in regions functionally-connected to the primary olfactory cortex, and (iii) greater reduction in global brain size. The infected participants also showed on average larger cognitive decline between the two timepoints. Importantly, these imaging and cognitive longitudinal effects were still seen after excluding the 15 cases who had been hospitalised. These mainly limbic brain imaging results may be the in vivo hallmarks of a degenerative spread of the disease via olfactory pathways, of neuroinflammatory events, or of the loss of sensory input due to anosmia. Whether this deleterious impact can be partially reversed, or whether these effects will persist in the long term, remains to be investigated with additional follow up.

---

While the global pandemic of severe acute respiratory syndrome coronavirus 2 (SARS-CoV-2) has now claimed millions of lives across the world, there has been increased focus by the scientific and medical community on the effects of mild-to-moderate COVID-19 in the longer term. There is strong evidence for brain-related pathologies, some of which could be a consequence of viral neurotropism^1,2^(doi.org/10.21203/rs.3.rs-1139035/v1), or of virus-induced neuroinflammation^3–5^(doi.org/10.1101/2021.02.23.432474): neurological and cognitive deficits demonstrated by patients^6,7^, with an incidence of neurological symptoms in more than 80% of the severe cases^8^, radiological and *post mortem* tissue analyses demonstrating the impact of COVID-19 on the brain^9,10^, and the possible presence of the coronavirus in the central nervous system found in some studies^11–13^.

In particular, one consistent clinical feature, which can appear before the onset of respiratory symptoms, is the disturbance in olfaction and gustation in COVID-19 patients^14,15^. In a recent study, 100% of the patients in the subacute stage of the disease were displaying signs of gustatory impairment (hypogeusia), and 86% either hyposmia or anosmia^16^. Such loss of sensory olfactory inputs to the brain could lead to a loss of grey matter in olfactory-related brain regions^17^. Olfactory — whether neuronal or supporting — cells concentrated in the olfactory epithelium are also particularly vulnerable to coronavirus invasion, and this seems to be also the case specifically with SARS-CoV-2^15,18–20^. Within the olfactory system, direct neuronal connections from and to the olfactory bulb encompass regions of the piriform cortex (the primary olfactory cortex), parahippocampal gyrus, entorhinal cortex, and orbitofrontal areas^21,22^.

Most brain imaging studies of COVID-19 to date have focused on acute cases and radiological reports of single cases or case series based on CT, PET or MRI scans, revealing a broad array of gross cerebral abnormalities ranging from white matter hyperintensities, hypoperfusion and signs of ischaemic events spread throughout the brain, but found more consistently in the cerebrum^9^. Of the few larger studies focusing on cerebrovascular damage using CT or MRI, some have either found no clear marker of abnormalities in the majority of their patients, or importantly no spatially consistent pattern for the distribution of white matter hyperintensities or microhaemorrhages, except perhaps in the middle or posterior cerebral artery territories and the basal ganglia^9^. Imaging cohort studies of COVID-19, quantitatively comparing data across subjects through automated preprocessing and co-alignment of images, are much rarer. For instance, a recent PET cohort study focusing on correlates of cognitive impairment has demonstrated, in 29 COVID-19 patients at a subacute stage, the involvement of fronto-parietal areas revealed as ^18^F-FDG hypometabolism^16^. Another glucose PET study has shown bilateral hypometabolism in the bilateral orbital gyrus rectus and the right medial temporal lobe^23^. One multi-organ imaging study^24^ (and its brain-focused follow-up^25^) in over 50 previously hospitalised COVID-19 patients suggested modest abnormalities in T2* of the left and right thalami compared with matched controls. It remains unknown however whether any of these abnormalities *predates* the infection by SARS-CoV-2. These effects could be associated with a pre-existing increased brain vulnerability to the deleterious effects of COVID-19 and/or a higher probability to show more pronounced symptoms, rather than being a consequence of the COVID-19 disease process.

UK Biobank offers a unique resource to elucidate these questions. With the data from this large, multi-modal brain imaging study, we use for the first time a longitudinal design whereby participants had been already scanned as part of UK Biobank *before* getting infected by SARS-CoV-2. They were then imaged again, on average 38 months later, after some had either medical and public health records for COVID-19, or had had two positive rapid antibody tests. Those participants were then matched with controls who had undergone the same longitudinal imaging protocol but had tested negative to the rapid antibody test or had no medical record of COVID-19. In total, 401 SARS-CoV-2 infected participants with usable imaging data at both timepoints were included in this study, as well as 384 controls, matched for age, sex, ethnicity and time elapsed between the two scans. These large numbers may allow us to detect subtle, but consistent spatially distributed sites of damage associated with the infection, thus underlining *in vivo* the possible spreading pathways of the effects of the disease within the brain (whether such effects relate to the invasion of the virus itself^11,18^ doi.org/10.21203/rs.3.rs-1139035/v1, inflammatory reactions^3,4^ doi.org/10.1101/2021. 02.23.432474, possible anterograde degeneration starting with the olfactory neurons in the nose, or through sensory deprivation^17,26,27^). The longitudinal aspect of the study aims to help tease apart which of the observed effects between first and second scans are likely related to the infection, rather than due to pre-existing risk factors between the two groups.

Our general approach in this study was therefore as follows: (i) use brain imaging data from 785 participants who visited the UK Biobank imaging centres for two scanning sessions, on average 3 years apart, with 401 of these having been infected with SARS-CoV-2 in between their two scans; (ii) estimate — from each subject’s multimodal brain imaging data — hundreds of distinct brain imaging-derived phenotypes (IDPs), each IDP being a measure of one aspect of brain structure or function; (iii) model confounding effects, and estimate the longitudinal change in IDPs between the two scans; and (iv) identify significant SARS-CoV-2 vs control group differences in these longitudinal effects, correcting for multiple comparisons across IDPs. We did this for both a focussed set of *a priori* defined IDPs, testing the hypothesis that the olfactory system is particularly vulnerable in COVID-19, as well as an exploratory set of analyses considering a much larger set of IDPs. In both cases we identified significant effects associated with SARS-CoV-2 infection primarily relating to greater atrophy and increased tissue damage in cortical areas directly connected to primary olfactory cortex, as well as to changes in global measures of brain and cerebrospinal fluid volume.

## Participants

UK Biobank has been releasing data from the COVID-19 re-imaging study on a rolling basis. As of the 31^st^ of May 2021, 449 adult participants met the re-imaging study inclusion criteria (see **Methods: Study Design**) and were identified as having been infected with SARS-CoV-2 based on either their primary care (GP) data, hospital records, results of their diagnostic antigen tests identified through record linkage to the Public Health datasets in England, Wales and Scotland, or two concordant antibody-based home lateral flow kit positive results. Of these 449 SARS-CoV-2 positive adult participants, a total of 401 had usable brain scans at both timepoints (**Tables 1 and 2**). For the 351 for whom we had a diagnosis date based on their medical records or antigen tests, the time between diagnosis (a proxy for infection) and their second imaging scan was on average 141 days (**Table 2**, **Supplementary Fig. 1**).

**Table 1.** Main demographics for the 401 SARS-CoV-2 positive cases and 384 controls. We used the ‘Last Observation Carried Forward’ (LOCF) imputation method (**Methods: Additional analyses — Baseline group comparisons**). Non-parametric tests were used whenever a variable for each group was not normally distributed (Lilliefors P < 0.05). Two-sample Kolmogorov-Smirnov test was used for age at Scan 1 or Scan 2, years between Scan 1 and Scan 2, alcohol intake frequency, and tobacco smoking; chi-square test for sex, ethnicity, and diagnosed diabetes; and Mann-Whitney U-test was used for the systolic and diastolic blood pressures, weight, waist/hip ratio, BMI and Townsend deprivation index.

|  | SARS-CoV-2 Positive Cases | Controls | P<sub>uncorr</sub> |
| --- | --- | --- | --- |
| Number of subjects | 401 | 384 | - |
| Age at Scan 1, mean±SD (range) | 58.9 ± 7.0 (46.9–80.2) | 60.2 ± 7.4 (47.1–79.8) | 0.15 |
| Age at Scan 2, mean±SD (range) | 62.1 ± 6.7 (51.3–81.4) | 63.3 ± 7.1 (51.3–81.3) | 0.08 |
| Sex, male/female | 172 (42.9%) / 229 (57.1%) | 164 (42.7%) / 220 (57.3%) | 0.96 |
| Ethnicity, white/non-white* | 388 (96.8%) / 13 (3.2%) | 373 (97.1%) / 11 (2.9%) | 0.76 |
| Years between Scans 1 and 2, mean±SD (range) | 3.2 ± 1.6 (1.0–7.0) | 3.2 ± 1.6 (1.0–6.9) | 0.98 |
| Systolic blood pressure [mmHg] | 130.3 ± 17.3 | 132.1 ± 17.6 | 0.16 |
| Diastolic blood pressure [mmHg] | 78.7 ± 10.6 | 79.0 ± 10.2 | 0.63 |
| Diagnosed diabetes | 18 (4.5%) | 16 (4.2%) | 0.82 |
| Weight [kg] | 76.4 ± 15.8 | 75.2 ± 14.4 | 0.65 |
| Waist/Hip ratio | 0.87 ± 0.09 | 0.86 ± 0.09 | 0.37 |
| BMI [kg/m<sup>2</sup>] | 26.7 ± 4.4 | 26.6 ± 4.3 | 0.61 |
| Alcohol intake frequency | 3.1 ± 1.3 | 3.0 ± 1.4 | 1.00 |
| Tobacco smoking | 0.61 ± 0.92 | 0.65 ± 0.89 | 0.87 |
| Townsend deprivation index | -1.5 ± 2.9 | -1.6 ± 2.9 | 0.65 |

\*The white/non-white distinction was made as numbers were too low to allow for a finer distinction

**Table 2.** Main clinical information available for the SARS-CoV-2 positive cases. Of note, of the 401 participants in our SARS-CoV-2 positive group in our main analyses, 50 were identified as cases via two different antibody home-based lateral flow kits and do not have date of diagnosis in their primary care or hospital records.

| | N, or mean ± SD (range) |
| --- | --- |
| **Total number of positive cases** | **401** |
| **Origin of diagnosis** |  |
| - GP | 11 |
| - Hospital | 2 |
| - Diagnostic antigen test from Public Health records | 338 |
| - Antibody home-based lateral flow kits | 50 |
| **Number of infected participants with available information on date of diagnoses** | **351** |
| **ΔDays of SARS-CoV-2 infection before Scan 2, mean±SD (range)** | **141±79 (35–407)** |
| **Total number of hospitalised patients** | **15** |
| - COVID-19 as primary cause | 11 |
| - COVID-19 as secondary cause | 4 |
| - Days of hospitalization, mean±SD (range) | 11.1 ± 11.0 (1–40) |
| - Critical care unit | 2 |
| - Invasive ventilation | 1 |
| - Continuous positive airway pressure | 1 |
| - Non-invasive ventilation | 1 |
| - Unspecified oxygen therapy | 1 |

In total, 384 adult controls met the inclusion criteria (see **Methods: Study Design**) and had usable brain scans at both timepoints (**Table 1**). SARS-CoV-2 positive or negative status was identified using UK Biobank Showcase variable 41000.

Despite the original matched-pairing of the COVID-19 patients and controls, their age distributions were slightly — though not statistically significantly — different, due to different patterns of missing/usable data (**Extended Data Fig. 1**). Note that the control group is on average slightly (not significantly) older than the SARS-CoV-2 positive group, which would be expected to make any change between the two timepoints harder to detect in the group comparisons, rather than easier. For histograms of interval of time between the two scans in the two groups, see **Extended Data Fig. 2**.

The two groups showed no statistical differences across all 6,301 non-imaging phenotypes after FDR or FWE correction for multiple comparisons (lowest P_fwe_=0.12, and no uncorrected P values survived FDR correction). However, due to the stringent correction for multiple comparisons that this analysis imposes, we investigated further whether subtle patterns of baseline differences could be seen using dimension reduction with principal component analysis on all 6,301 variables, and using a separate principal component analysis focused on baseline cognition (see **Supplementary Analysis 1**). We found no principal component that differed significantly between the two groups when exploring all the non-imaging variables. With respect to cognitive tests, while no single cognitive score was significantly different at baseline between controls and future cases, we identified two cognitive principal components that were different (**Supplementary Analysis 1**). These subtle baseline cognitive differences suggest slightly lower cognitive abilities for the future cases when compared with the controls. Importantly, none of these principal components—cognitive or otherwise— could statistically account for the longitudinal imaging results (see below, **Additional baseline investigations**).

Through hospital records available for participants, we identified 15 of the SARS-CoV-2 positive group who were hospitalised with COVID-19, including 2 who received critical care (**Tables 2 and 3**). These hospitalised patients were on average older, had higher blood pressure and weight, and were more likely to have diabetes and to be men, compared with non-hospitalised cases (**Table 3**).

**Table 3.** Comparison between hospitalised vs non-hospitalised SARS-CoV-2 positive cases. For statistical procedures, please refer to Table 1.

|  | Hospitalised | Non-hospitalised | P<sub>uncorr</sub> |
| --- | --- | --- | --- |
| Number of subjects | 15 | 386 | - |
| Age at Scan 1, mean±SD (range) | 65.4 ± 8.9 (51.6–80.2) | 58.7 ± 6.8 (46.9–77.0) | 0.0028 |
| Age at Scan 2, mean±SD (range) | 68.1 ± 8.4 (54.9–81.4) | 61.9 ± 6.5 (51.3–80.0) | 0.0058 |
| Sex, male/female | 10 (66.7%) / 5 (33.3%) | 162 (42.0%) / 224 (58.0%) | 0.058 |
| Ethnicity, white/non-white* | 15 (100%) / 0 (0%) | 373 (96.6%) / 13 (3.4%) | 0.47 |
| Years between Scan 1 and 2, mean±SD (range) | 2.7 ± 1.4 (1.0–5.8) | 3.2 ± 1.6 (1.1–7.0) | 0.50 |
| Systolic blood pressure [mmHg] | 140.6 ± 16.6 | 129.9 ± 17.2 | 0.022 |
| Diastolic blood pressure [mmHg] | 85.0 ± 10.5 | 78.4 ± 10.5 | 0.028 |
| Diagnosed diabetes | 4 (26.7%) | 14 (3.6%) | < 0.001 |
| Weight [kg] | 85.9 ± 12.0 | 76.0 ± 15.8 | 0.0072 |
| Waist/Hip ratio | 0.94 ± 0.07 | 0.87 ± 0.09 | 0.0015 |
| BMI [kg/m<sup>2</sup>] | 29.3 ± 3.7 | 26.6 ± 4.4 | 0.0076 |
| Alcohol intake frequency | 3.1 ± 1.7 | 3.1 ± 1.3 | 1.00 |
| Tobacco smoking | 0.80 ± 1.0 | 0.60 ± 0.91 | 0.75 |
| Townsend deprivation index | -2.1 ± 2.6 | -1.5 ± 2.9 | 0.42 |

\*The white/non-white distinction was made as numbers were too low to allow for a finer distinction

### Hypothesis-driven results

The main case-vs-control analysis between the 401 SARS-CoV-2 positive cases and 384 controls (Model 1) on 297 olfactory-related cerebral IDPs yielded 68 significant results after FDR correction for multiple comparisons, including 6 further surviving FWE correction (**Table 4**, **Fig. 1, Supplementary Table 1** for full list of results). Focusing on the top 10 most significant associations, 8 of these IDPs covered similar brain regions functionally-connected to the primary olfactory cortex (see **Methods: Hypothesis-driven approach**), showing overlap especially in the anterior cingulate cortex, orbito-frontal cortex and insula, as well as in the ventral striatum, amygdala, hippocampus and parahippocampal gyrus^28^. We found greater longitudinal increase in diffusion indices for the SARS-CoV-2 group in these tailored IDPs defining the functional connections with the frontal and temporal piriform cortex, as well as the olfactory tubercle and anterior olfactory nucleus (**Table 4**, **Fig. 1, Supplementary Table 1**). The other two of the top 10 IDPs encompassed the left lateral orbitofrontal cortex and parahippocampal gyrus, both showing greater reduction of grey matter thickness or intensity contrast over time in the cases compared with controls (**Table 4**, **Fig. 1, Supplementary Table 1**). For those significant IDPs, average percentage change differences between the two groups was moderate, ranging from ∼0.2 to ∼2%, with the largest differences seen in the volume of the parahippocampal gyrus and entorhinal cortex (**Supplementary Table 1**). Scatter and box plots, as well as plots showing percentage longitudinal differences with age are available for the top 10 longitudinal IDPs as **Supplementary Longitudinal Plots**.

**Fig. 1:**
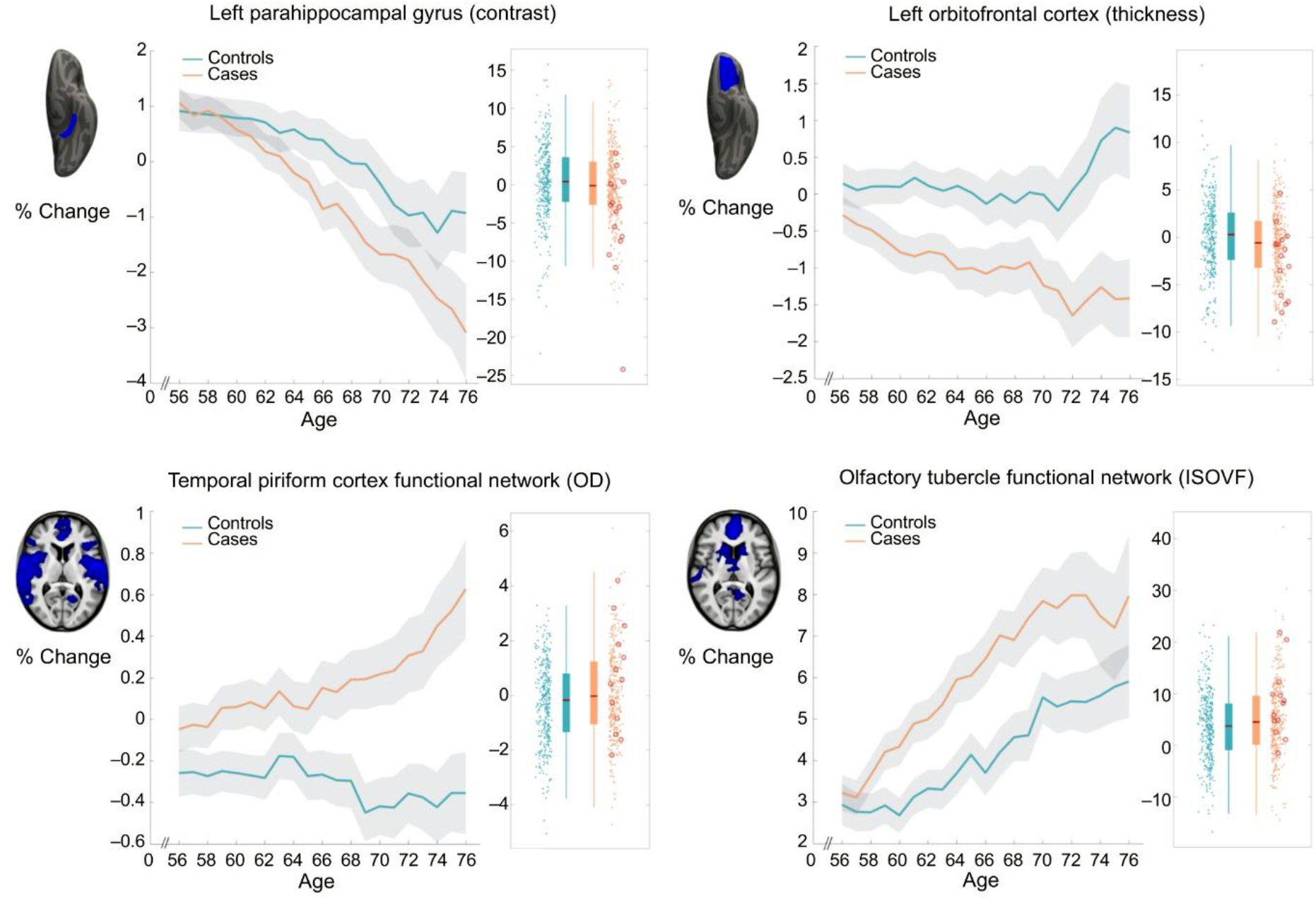
Most significant longitudinal group comparison results - hypothesis-driven approach. The top four regions consistently showing longitudinal differences across the three models comparing SARS-CoV-2 cases and controls demonstrated either a significantly greater reduction in grey matter thickness and intensity contrast, or an increase in tissue damage (largest combined |Z| across Models 1-3). All three models pointed at the involvement of the parahippocampal gyrus, while Models 1 and 2 also showed the significant involvement of the functional connections of the primary olfactory cortex and of the left orbitofrontal cortex. For each region, the IDP’s spatial region of interest is shown at the top in blue, overlaid either on the FreeSurfer average inflated cortical surface, or the T1 template (left is shown on right). Bottom left for each IDP are the longitudinal percentage changes with age for the two groups (controls in blue, infected participants in orange), obtained by normalising ΔIDP using as baseline the values for the corresponding IDPs across the 785 participants’ scans. These are created using a 10-year sliding window average, with standard errors in grey. The somewhat counter-intuitive increase in thickness in the orbitofrontal cortex in older controls has been previously consistently reported in studies of ageing^54,55^. Bottom right are the scatter and box plots showing the difference in cortical thickness, intensity contrast, or diffusion indices between the two timepoints for the 384 controls (blue) and 401 infected participants (orange), allowing the visual comparison between the two groups in a binary way (hence under-estimating the effects estimated when modulating with age, see **Methods: Statistical Modelling — Main longitudinal model, deconfounding**). In red circles are the 15 hospitalised patients. OD, orientation dispersion; ISOVF, isotropic volume fraction. All y axes represent % change.

**Table 4.** Hypothesis-driven olfactory approach: top 10 out of 68 significant longitudinal group comparison results. The top 10 significant results, all surviving false discovery rate (FDR) correction, based on 297 imaging-derived phenotypes (IDPs), ranked based on their uncorrected P-values for our main analysis (Model 1), showing where the 401 SARS-CoV-2 infected participants and 384 controls differed over time. Associations with a total of 68 IDPs in total survived correction for multiple comparisons using FDR for Model 1 (full list of results in **Supplementary Table 1**). We report differences in longitudinal change (as a % of mean baseline value) between the two groups, standard error (SE) on these % changes for Model 1, as well as uncorrected and family-wise error (FWE) corrected P values. Results in italics survive correction for multiple comparisons using FDR for each corresponding Model. Results in bold also survive correction for multiple comparisons using FWE for each corresponding Model. <u>Note</u>: the Z-statistics reflect the statistical strength of the longitudinal group-difference modelling, and are not raw data effect sizes. All significant results involved either grey matter thickness, grey-white intensity contrast or proxy measures of tissue damage (mean diffusivity MD, isotropic volume fraction ISOVF, and orientation dispersion OD). L is left.

|  | Main Analysis (Model 1):<br>All SARS-CoV-2 cases (n=401)<br>vs controls (n=384) |  |  |  |  | Model 2:<br>Non-hospitalised cases (n=386)<br>vs controls (n=384) |  |  | Model 3:<br>Hospitalised cases (n=15)<br>vs controls (n=384) |  |  | Model 4:<br>Hospitalised (n=15) vs non-hospitalised cases (n=386) |  |  |
| --- | --- | --- | --- | --- | --- | --- | --- | --- | --- | --- | --- | --- | --- | --- |
| Imaging-Derived Phenotype (IDP) | % | SE | Z | Puncorr | Pfwe | Z | Puncorr | Pfwe | Z | Puncorr | Pfwe | Z | Puncorr | Pfwe |
| Temporal piriform cortex functional network - OD | **0.34** | **0.08** | **4.2** | **0.000023** | **0.0068** | **3.9** | **0.000081** | **0.0217** | 2.5 | 0.013985 | 0.9176 | 0.5 | 0.627678 | 1 |
| Olfactory tubercle functional network - ISOVF | **1.22** | **0.31** | **3.9** | **0.000102** | **0.028** | 3.4 | 0.000623 | 0.1319 | 2.8 | 0.004439 | 0.6576 | 1.4 | 0.155595 | 1 |
| Frontal piriform cortex functional network - MD | **0.39** | **0.1** | **3.8** | **0.000146** | **0.0386** | 3.4 | 0.000671 | 0.1417 | 2.4 | 0.017728 | 0.9532 | 1.3 | 0.202213 | 1 |
| Temporal piriform cortex functional network - MD | **0.38** | **0.1** | **3.8** | **0.00015** | **0.0396** | 3.3 | 0.000849 | 0.1717 | 2.5 | 0.011746 | 0.8913 | 1.4 | 0.172706 | 1 |
| Olfactory tubercle functional network - MD | **0.39** | **0.1** | **3.8** | **0.000171** | **0.0446** | 3.3 | 0.000946 | 0.188 | 2.5 | 0.011856 | 0.8931 | 1.4 | 0.173673 | 1 |
| Lateral orbitofrontal cortex L - thickness (DKT atlas) | **-0.76** | **0.2** | **-3.8** | **0.000172** | **0.0449** | -2.9 | 0.003178 | 0.48 | -3.1 | 0.001957 | 0.4541 | -1.9 | 0.061739 | 0.9999 |
| Temporal piriform cortex functional network – ISOVF | 1.12 | 0.3 | 3.7 | 0.000222 | 0.0564 | 3.3 | 0.001068 | 0.2088 | 2.6 | 0.009838 | 0.8562 | 1.3 | 0.18374 | 1 |
| Anterior olfactory nucleus functional network – MD | 0.42 | 0.11 | 3.7 | 0.000242 | 0.0621 | 3.5 | 0.000559 | 0.1213 | 1.9 | 0.060313 | 0.9993 | 0.8 | 0.432506 | 1 |
| Parahippocampal gyrus L - intensity contrast (Desikan) | -0.92 | 0.25 | -3.7 | 0.000276 | 0.0685 | -2.7 | 0.006201 | 0.704 | -3.5 | 0.000507 | 0.212 | -2.0 | 0.042678 | 0.998 |
| Anterior olfactory nucleus functional network - ISOVF | 1.27 | 0.35 | 3.6 | 0.000286 | 0.0701 | 3.3 | 0.000985 | 0.1957 | 2.4 | 0.016069 | 0.9399 | 1.1 | 0.268503 | 1 |

As secondary analyses, we found that significant longitudinal differences remained in the same set of significant brain regions surviving FDR or FWE correction when removing from the SARS-CoV-2 group those patients who had been hospitalised with COVID-19 (Model 2, 47 IDPs FDR-significant, 3 of which also FWE-significant, **Supplementary Table 1**). While fewer results were significant for the comparison between the 15 hospitalised patients and 384 controls (Model 3, 4 results FDR-significant, **Supplementary Table 1**), likely due to the large reduction in sample size for this model, this additional group comparison showed effects in the same regions of the parahippocampal gyrus, orbital cortex, and superior insula. Finally, we found no significant differences between the 15 hospitalised patients and 386 non-hospitalised SARS-COV-2 cases, likely due to the large reduction in sample size, but effect sizes and direction of these effects suggested stronger detrimental effects for the hospitalised cases in the orbitofrontal, insula, parahippocampal and frontal piriform cortex functionally-connected brain regions (all |Z|≥3, Model 4, **Supplementary Table 1**).

Across the 3 models comparing SARS-CoV-2 cases with controls (Models 1-3), the top 4 longitudinal differences were found in the functionally-connected regions of the temporal piriform cortex (diffusion index: orientation dispersion) and of the olfactory tubercle (diffusion index: isotropic volume fraction), as well as in the parahippocampal gyrus (intensity contrast) and lateral orbitofrontal cortex (thickness) (largest combined |Z| across Models 1-3; **Fig. 1**). For these results across Models 1-3, the percentage of SARS-CoV-2 infected participants who showed a greater longitudinal change than the median value in the controls was: 56% for the regions connected to the temporal piriform cortex, 62% for the regions connected to the olfactory tubercle, 57% left parahippocampal gyrus and 60% for the left orbitofrontal cortex.

While significant IDPs related to grey matter thickness were found, using our main case-vs-control analysis (Model 1), to be bilateral for both the anterior parahippocampal gyrus (perirhinal cortex) and entorhinal cortex, 10 of the 11 remaining significant IDP were left-lateralised (**Supplementary Table 1**). We thus directly investigated (left - right) differences in the SARS-CoV-2 group only for those significant IDPs, and found that the infected participants did not have significantly more reduced grey matter thickness on the left than on the right hemisphere (lowest P_uncorr_=0.30).

Of the top 10 IDPs showing a longitudinal effect between first and second scans, none correlated significantly with the time interval between their infection and their second scan, in the SARS-CoV-2 positive participants for whom we had a date of diagnosis (n=351; lowest P_uncorr_=0.08).

### Exploratory results

2,047 IDPs passed the initial tests of reproducibility (**Extended Data Fig. 3**) and data completeness. The main analysis (Model 1) revealed 65 significant longitudinal differences between the cases and controls passing FDR correction, including 5 that were FWE-significant (**Table 5**, **Supplementary Table 1** for the complete list of reproducible IDPs and results). **Extended Data Figs. 4** and **5** show the QQ plot relating to the FDR thresholding, and a summary figure of Z-statistics results for all 2,047 IDPs grouped into different IDP classes.

**Table 5.**
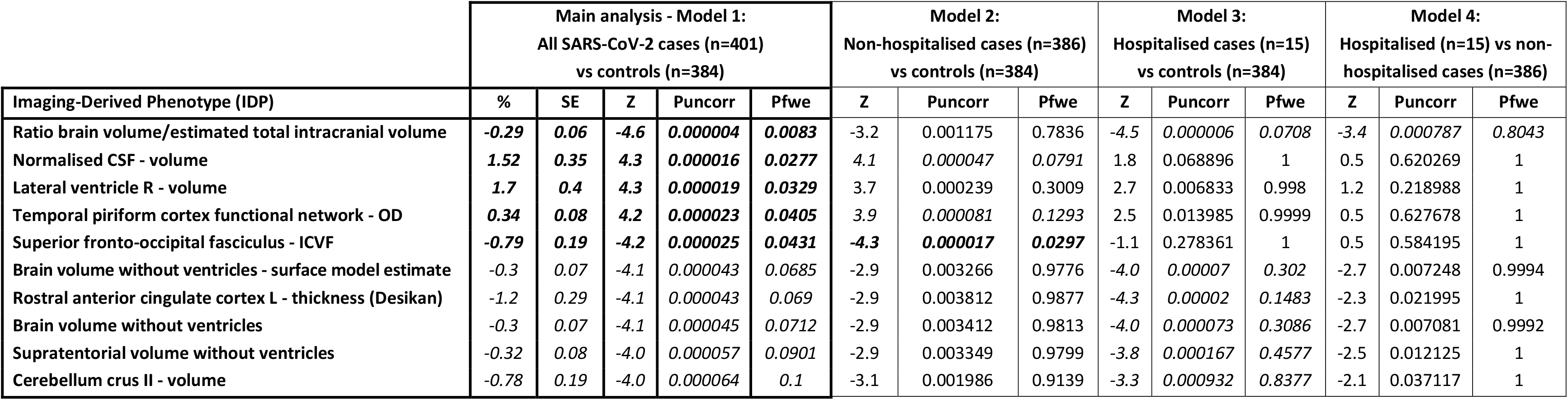
Exploratory approach: top 10 out of 65 significant longitudinal group comparison results. The top 10 significant results show where the 401 SARS-CoV-2 positive participants and 384 controls differed over time, ranked based on their uncorrected P-values for Model 1. Associations with a total of 65 imaging-derived phenotypes (IDPs) survived correction for multiple comparisons using false discovery rate (FDR) for Model 1 (full list of results in **Supplementary Table 1**). In italics, the findings surviving correction for multiple comparisons using FDR for each Model; in bold, those surviving using family-wise error (FWE). <u>Note</u>: the Z-statistics reflect the statistical strength of the longitudinal group-difference modelling, and are not raw data effect sizes. In addition to global measures relating to loss of brain volume (such as an increase of CSF volume), most of the top exploratory localised results implicate the primary connections of the olfactory system, as well as the rostral anterior cingulate cortex and the crus II of the cerebellum, both also olfactory-related regions. Intra-cellular volume fraction ICVF, orientation dispersion OD. L is left, R is right.

In particular, in this exploratory analysis covering the entire brain, 33 out of the 65 significant IDPs overlapped with the IDPs selected *a priori* for our hypothesis-driven approach of the involvement of the olfactory system. In addition, we found significant longitudinal effects in global measures of volume, such as the CSF volume normalised for head size and the ratio of the volume of the segmented brain to the estimated total intracranial volume generated by FreeSurfer, as well as in the volume of the left crus II of the cerebellum, the thickness of the left rostral anterior cingulate cortex and diffusion index in the superior fronto-occipital fasciculus (**Table 5**, **Supplementary Table 1**, see examples in **Extended Data Fig. 6**). For those significant IDPs, average percentage change differences between the two groups was moderate, ranging from ∼0.2 to ∼2% (except for two diffusion measures in the fimbria at >6%, due to the very small size of these regions-of-interest), with the largest differences seen in the volume of the parahippocampal gyrus and caudal anterior cingulate cortex (**Supplementary Table 1**). Scatter and box plots, as well as plots showing percentage longitudinal differences with age are available for the top 10 longitudinal IDPs as **Supplementary Longitudinal Plots**.

For the secondary analyses, when comparing the non-hospitalised cases to the controls (Model 2), the same general pattern emerged, albeit with a reduced number of significant results: one olfactory-related region, the functionally-connected areas to the temporal piriform cortex, showed significant longitudinal difference between the two groups in diffusion index, as well as one global volume measure (CSF normalised), and diffusion index in the superior fronto-occipital fasciculus (Model 2, 4 FDR-corrected, 1 FWE-corrected, **Supplementary Table 1**). Despite the considerably limited degrees of freedom in Models 3 and 4, many results survived multiple comparison correction, particularly for IDPs of cortical thickness, with an emphasis on the anterior cingulate cortex for Model 3 (66 FDR-corrected, 3 FWE-corrected), and a wide distribution across prefrontal, parietal and temporal lobes for Model 4 (29 FDR-corrected, **Fig. 2**).

**Fig. 2.**
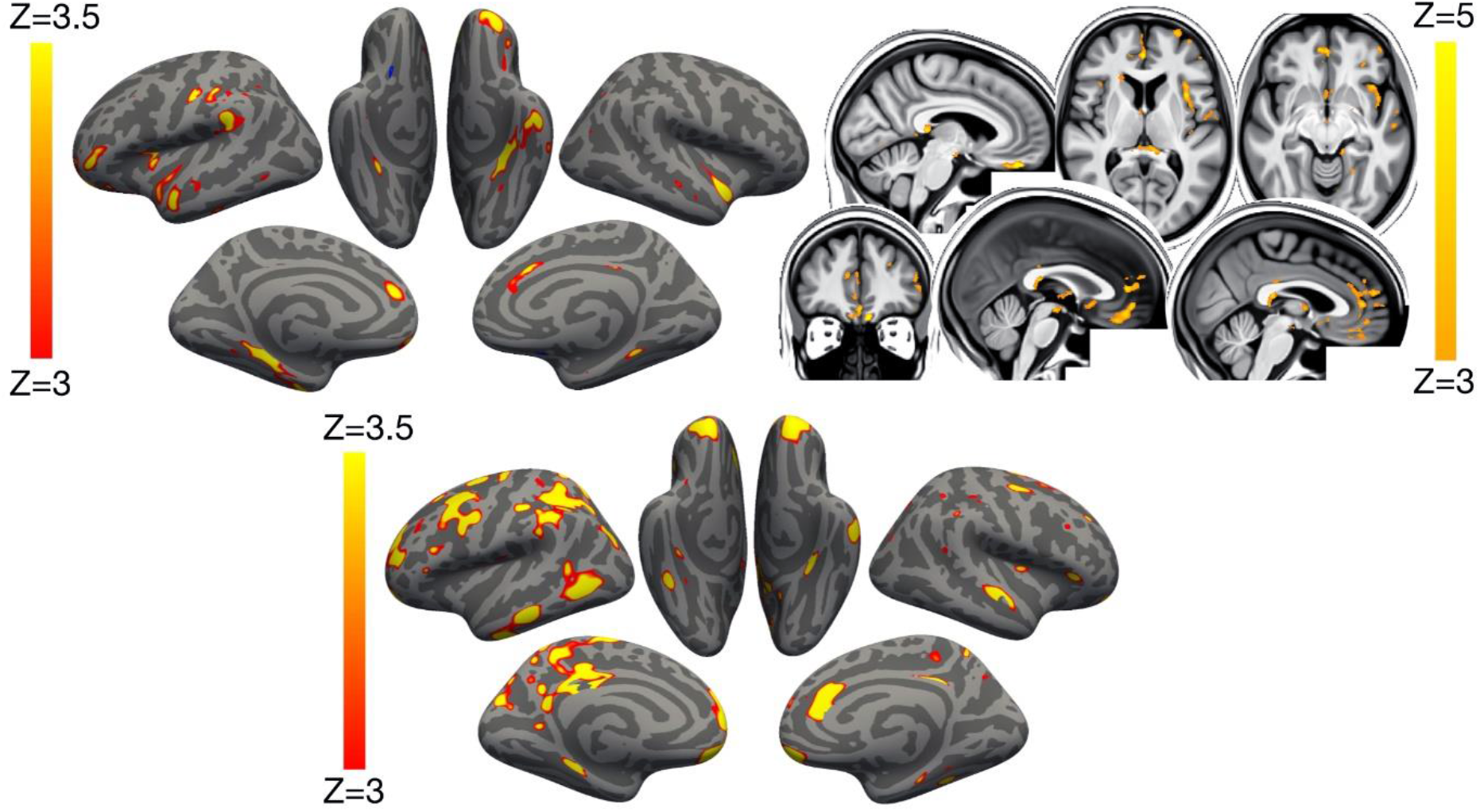
Vertex-wise and voxel-wise longitudinal group differences in grey matter thickness and mean diffusivity changes. Top row. Main analysis (Model 1): The thresholded map (|Z|>3) shows that the strongest, localised reduction of grey matter thickness in the 401 SARS-CoV-2 positive participants compared with the 384 controls are bilaterally in the parahippocampal gyrus, anterior cingulate cortex and temporal pole, as well as in the left orbitofrontal cortex, insula and supramarginal gyrus. Similarly, the strongest longitudinal differences in mean diffusivity (|Z|>3, left is shown on right) could be seen in the orbitofrontal cortex and anterior cingulate cortex, as well as in the left insula and amygdala. **Bottom row.** Secondary analysis (Model 4): The thresholded cortical thickness map (|Z|>3) demonstrated longitudinal differences between the 15 hospitalised and 386 non-hospitalised SARS-CoV-2 positive cases in the orbitofrontal frontal cortex and parahippocampal gyrus bilaterally, right anterior cingulate cortex, as well as marked widespread differences in fronto-parietal and temporal areas, especially in the left hemisphere. We show the voxel-wise or vertex-wise longitudinal effects for illustrative purposes, avoiding any thresholding based on significance (as this would be statistically circular - similar to our previous analyses reported in ^56^).

As many of the top exploratory and hypothesis-driven results included IDPs of cortical thickness and of mean diffusivity, we further conducted an exploratory visualisation of the vertex-wise thickness, and voxel-wise mean diffusivity longitudinal differences between the cases and controls over the entire cortical surface and brain volume, respectively (**Fig. 2**). Grey matter thickness showed bilateral longitudinal differences in the parahippocampal gyrus, anterior cingulate cortex and temporal pole, as well as in the left orbitofrontal cortex, insula and supramarginal gyrus.

When visually comparing hospitalised and non-hospitalised cases, these longitudinal differences showed a similar pattern, especially in the parahippocampal gyrus, orbitofrontal and anterior cingulate cortex, but also markedly extending, particularly in the left hemisphere, to many fronto-parietal and temporal regions. Mean diffusivity differences in longitudinal effects between cases and controls was seen mainly in the orbitofrontal cortex, anterior cingulate cortex, as well as in the left insula and amygdala.

While results seen in IDPs of grey matter thickness seemed to indicate that the left hemisphere is more strongly associated with SARS-CoV-2 infection, a direct (left - right) comparisons of all lateralised IDPs of thickness across the entire cortex showed no overall statistical difference between the two groups (lowest P_fwe_=0.43, and with no results surviving FDR correction).

### Cognitive results

Using the main model used to compare longitudinal imaging effects between SARS-CoV-2 positive participants and controls (Model 1), we explored differences between the two groups in 10 scores from 6 cognitive tasks. These 10 scores were selected using a data-driven approach based on out-of-sample participants who are the most likely to show cognitive impairment (**Supplementary Analysis 2**). After FDR correction, we found a significantly greater increase of the time taken to complete Trails A (numeric) and B (alphanumeric) of the Trail Making Test in the SARS-CoV-2 infected group (Trail A: 7.8%, P_uncorr_=0.0002, P_fwe_=0.005; Trail B: 12.2%, P_uncorr_=0.00007, P_fwe_=0.002; **Fig. 3**). These findings remained significant when excluding the 15 hospitalised cases (Model 2: Trail A: 6.5%, P_uncorr_=0.002, P_fwe_=0.03; Trail B: 12.5%, P_uncorr_=0.00009, P_fwe_=0.002).

**Fig. 3.**
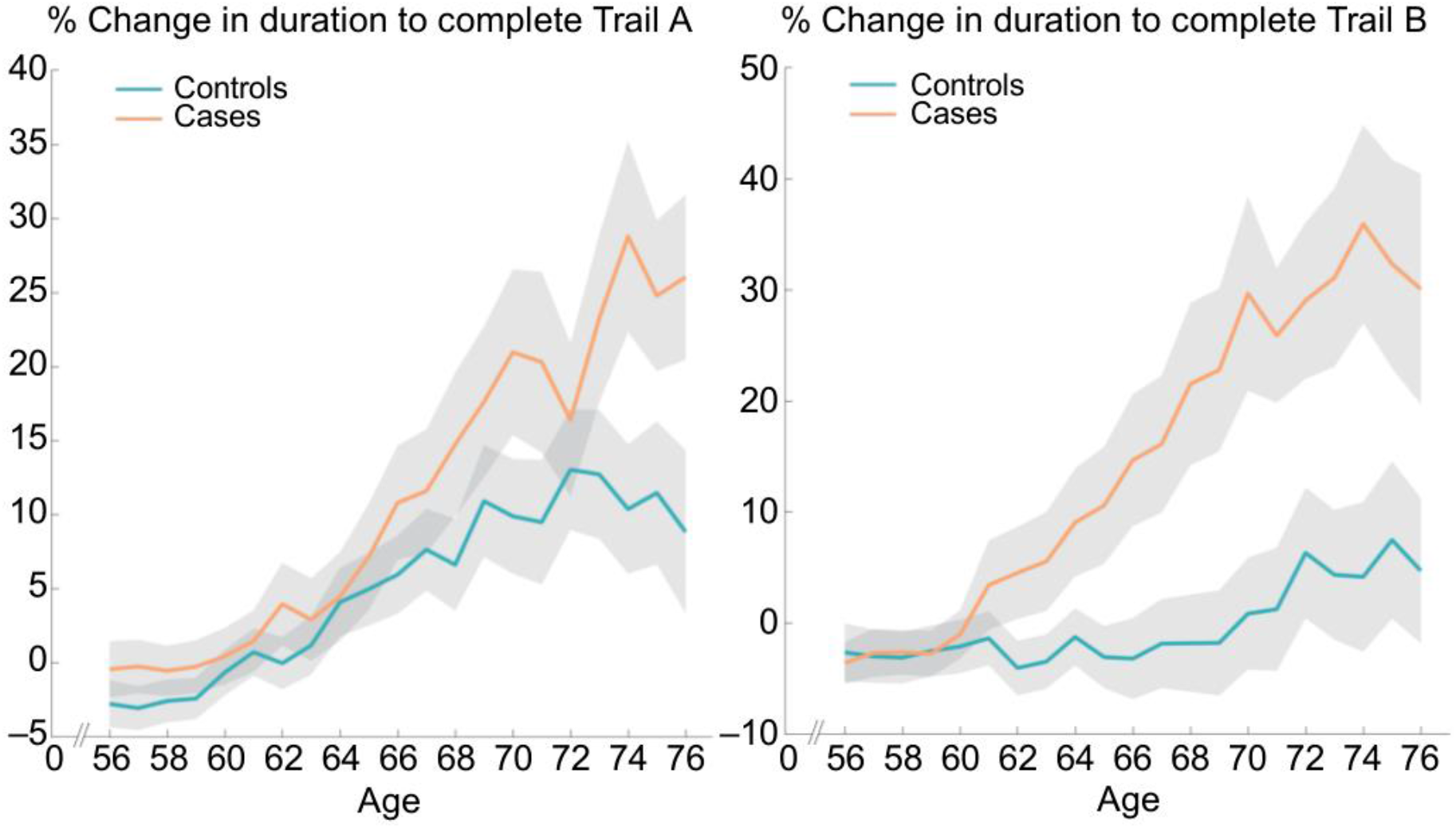
Percentage longitudinal change for SARS-CoV-2 positive participants and controls, in the duration to complete Trails A and B of the UK Biobank Trail Making Test. Absolute baseline (used to convert longitudinal change into percent change) estimated across the 785 participants. These curves were created using a 10-year sliding window across cases and controls (standard errors in grey).

In the SARS-CoV-2 group only, *post hoc* associations between the most significant cognitive score showing longitudinal effect using Model 1 (duration to complete Trail B, as reported above) and the top 10 results from each of the hypothesis-driven and exploratory approaches revealed a significant longitudinal association with the volume of the mainly cognitive lobule crus II of the cerebellum (r=-0.19, Pfwe=0.020).

### Additional baseline investigations

When looking at binary baseline differences between controls and future cases, none of the IDPs with significant longitudinal effects for either hypothesis-driven or exploratory approaches demonstrated significant differences at baseline between the two groups (lowest P_fwe_=0.59, nothing surviving FDR correction; **Supplementary Table 2**). When applying age-modulation in the two-group modelling of IDPs at baseline, a few of the IDPs demonstrated significant differences between control and future SARS-CoV-2 groups, mainly for diffusion indices in the olfactory functional networks, as well as in the subcortical grey matter. As some IDPs cover spatially extended regions of the brain, we visually explored whether these baseline differences had any spatial overlap with our longitudinal results, but found none (**Supplementary Fig. 2**). The full list of (binary and age-modulated) results from group comparisons between the two groups at baseline are available in **Supplementary Table 2** (and separately, at the second timepoint, in **Supplementary Table 3**). We also provide the scatter and boxplots, as well as the percentage differences with age at baseline for the top 10 significant longitudinal IDPs from the hypothesis-driven and exploratory approaches (**Supplementary Plots**).

In addition, none of the 10 pre-selected cognitive variables showed significant difference at baseline between SARS-CoV-2 and control groups (min P_uncorr_=0.08). With age-modulation, only one cognitive score, time to complete Pairs Matching round, showed a trend difference at baseline (P_uncorr_<0.05, P_fwe_=0.29, not passing FDR). This is a different cognitive score from the one showing longitudinal cognitive effects between the two groups, the UK Biobank Trail Making Test.

We also repeated the main analysis modelling for those top 10 IDPs found to show longitudinal differences between the SARS-CoV-2 and control groups, across both hypothesis-driven and exploratory approaches. For each of 6,301 non-imaging variables available (see **Methods: Additional analyses — Baseline group comparisons**), we included that variable as an additional confounder in the longitudinal analyses. On the basis of the regression Z-statistic values, the strength of the original associations was not reduced by more than 25% for any of the non-imaging variables.

We further carried out the same analyses, but using dimension reduction (principal component analysis) applied to these 6,301 non-imaging phenotypes (d=1 to d=700), and also just focusing on cognition, with 540 cognitive variables (d=10). We found no substantial reduction in our longitudinal results with any of these principal components. In particular, for cognition where two components were significantly different at baseline (PC1 and PC4, **Supplementary Analysis 1**), the strongest reduction in Z was found for crus II of the cerebellum when adding PC1 to the model, with a decrease in Z of only 5.7 % (from Z=4 to Z=3.77), while the Z values associated with all the other IDPs were reduced by less than 5%. Adding PC4 to our main model reduced Z by 0.4% at most.

### Additional, out-of-sample tests of longitudinal effects of pneumonia and influenza

To investigate whether pneumonia might have had an impact on our longitudinal findings, we assessed the age-modulated effects associated with pneumonia in an out-of-sample UK Biobank cohort that had been scanned twice. We identified 11 participants who contracted pneumonia not related to COVID-19 between the two scans, matched these to 261 controls, and applied our main analysis (Model 1) to these two groups. This longitudinal investigation showed some significant group differences in IDPs, but with no overlap with those IDPs we found for SARS-CoV-2 (all in the white matter, **Supplementary Analysis 3**). Overall, correlation between all IDPs’ (unthresholded) Z-statistics from pneumonia and SARS-CoV-2 longitudinal group comparisons was very low (r=0.057).

The sample size of cases who contracted influenza between the two scans in the out-of-sample UK Biobank cohort was unfortunately much smaller (n=5, including n=3 hospitalised cases), likely due to the low probability of influenza being recorded by a medical professional (GP or hospital). Nevertheless, for completeness, we also assessed longitudinally these two very small groups, compared with 127 matched controls. No result was significant for the 5 influenza cases, although a few IDPs showed significant longitudinal age-modulated effects, with just one IDP in the brainstem common to the SARS-CoV-2 findings (**Supplementary Analysis 4**). Correlation of Z-statistics between influenza and SARS-CoV-2 longitudinal group comparisons was again low (r=0.077).

## Discussion

To our knowledge, this is the first longitudinal imaging study of SARS-CoV-2 where participants were initially scanned *before* any had been infected. Our longitudinal analyses revealed a significant, deleterious impact associated with SARS-CoV-2. This impact could be seen mainly in the limbic and olfactory cortical system, for instance with a change in diffusion measures — that are proxies for tissue damage — in regions functionally connected with the piriform cortex, olfactory tubercle and anterior olfactory nucleus, as well as a more pronounced reduction of grey matter thickness and contrast in the SARS-CoV-2 infected participants in the left parahippocampal gyrus and lateral orbitofrontal cortex. While the greater atrophy for the SARS-CoV-2 positive participants was localised to a few, mainly limbic, regions, the increase in CSF volume and decrease of whole brain volume suggests an additional diffuse loss of grey matter superimposed onto the more regional effects observed in the olfactory-related areas. It is worth noting that these structural and microstructural longitudinal significant differences are modest in size, the strongest differences in changes observed between the SARS-CoV-2 positive and control groups corresponding to around 2% of mean baseline IDP value (**Supplementary Table 1**). This additional loss in the infected participants of 0.7% on average across the olfactory-related brain regions — and specifically ranging from 1.3% to 1.8% for the FreeSurfer volume of the parahippocampal/perirhinal and entorhinal cortex — can be helpfully compared with, for instance, the longitudinal loss per year of ∼0.2% (in middle age) to 0.3% (in older age) of hippocampal volume in community-dwelling individuals^29^. Our statistics also represent an *average* effect; not every infected participant will display brain longitudinal abnormalities. Comparing the few patients (n=15) who had been hospitalised with COVID-19 against non-hospitalised cases showed a more widespread pattern of greater reduction in grey matter thickness in fronto-parietal and temporal regions (**Fig. 2**). Finally, significantly greater cognitive decline, which persisted even after excluding the hospitalised patients, was seen in the SARS-CoV-2 positive group between the two timepoints, and this decline was associated with greater atrophy of crus II, a cognitive lobule of the cerebellum.

Much has been made of the benefit of using a longitudinal design to estimate, for example, trajectories of brain ageing and cognitive decline^30,31^. The longitudinal nature of the UK Biobank COVID-19 re-imaging study, with the baseline scan acquired *before* infection by SARS-CoV-2 and the second scan *after* infection, reveal differences over time *above and beyond* any potential baseline differences, thereby helping disentangle the (direct or indirect) contribution of the pathogenic process from pre-existing differences in the brain, or risk factors, of future COVID-19 patients. An illustrative example of the benefit of a longitudinal design is that, if looking solely at cross-sectional group comparisons at the second timepoint post infection (i.e., the analysis that would, by necessity, be carried out in *post hoc* studies), the strongest effect is seen in the volume of the thalamus. This effect disappears when taking into account the baseline scans however, since the thalamus of the participants who will later become infected appears to already differ from the controls years before infection. This highlights the difficulties in interpreting cross-sectional post-infection imaging differences as being *necessarily* the consequence of the infection itself. When looking at brain imaging baseline differences between the two groups across all IDPs, particularly in an age-modulated way, we did find a few further significant baseline differences beyond the volume of the thalamus (**Supplementary Table 2**). These were principally using diffusion imaging, but also using grey matter volume in the subcortical structures. Importantly, none of these baseline imaging differences spatially overlapped with the regions found to be different longitudinally (**Supplementary Fig. 2**). As this study is observational, as opposed to a randomised interventional study, one cannot make claims of disease causality with absolute certainty however, but interpretational ambiguities are greatly reduced compared with *post hoc* cross-sectional studies. The question remains as to whether the two groups are actually perfectly matched, as controls and cases could not be randomised *a priori*. Across the main risk factors, as well as thousands of lifestyle, health data and environment variables available in UK Biobank, we did not identify any significant differences when looking at each variable in isolation (only a few variables showed some trends at P<0.001 uncorrected, see **Supplementary Table 4**). This does not preclude the possibility of a sub-threshold pattern of baseline differences making one group more at risk of getting infected by SARS-CoV-2, and this risk perhaps interacting with the effects of the coronavirus. This motivated the use of principal component analyses, which revealed two significant components suggesting subtle lower cognitive abilities in the participants who got infected later on (**Supplementary Analysis 1**). Importantly, neither of these two cognitive components had any bearing on our longitudinal imaging results (reducing at most the strength of Z-statistics from Z=4 to Z=3.77 for the crus II of the cerebellum, when added in as an extra confound to the longitudinal analysis). Whether any of these imaging and cognitive differences at baseline played a subsequent role in those patients being more likely to get infected by the coronavirus, or to develop symptoms from infection, would need further investigation.

Our cohort-based, quantitative imaging study, unlike the majority of single case and case series studies published so far, does not focus on gross abnormalities that could be observed at the single subject-level with a naked eye, such as microhaemorrhages or (sub)acute ischaemic infarctions^9^. It does however rely on an anatomically *consistent* pattern of abnormalities caused by the disease process, a common spatial distribution of these pathological alterations across the infected participants, which could be uncovered by aligning all the images together in a common space, followed by applying a pipeline of modality-specific image processing algorithms. This automated, objective and quantitative processing of the images facilitates the detection of subtle changes that would not be visible at the individual level, but which point to a possible mechanism for the neurological effects of the coronavirus infection. Our hypothesis-driven analyses revealed a clear involvement of the olfactory cortex, which was also found in the exploratory analyses and the vertex-wise and voxel-wise maps of cortical thickness and mean diffusivity. While no differences were seen in the olfactory bulbs or piriform cortex *per se* (both located in a region above the sinuses prone to susceptibility distortions in the brain images, and both being difficult to segment in MRI data), we identified significant longitudinal differences in a network of regions functionally-connected to the piriform cortex, mainly constituted of the anterior cingulate cortex and orbitofrontal cortex, as well as the ventral striatum, amygdala, hippocampus and parahippocampal gyrus^28^. Some of the most consistent abnormalities across hypothesis-driven and exploratory analyses and all group comparisons were revealed in the left parahippocampal gyrus (**Table 4**, **Fig. 2**, **Supplementary Table 1**) — a limbic region of the brain that plays a crucial, integrative role for the relative temporal order of events in episodic memory^32–34^. Importantly, it is directly connected to the piriform cortex and entorhinal cortex, which are both part of the primary olfactory cortex^22,35^. Similarly, the orbitofrontal cortex, which we also found altered in the SARS-CoV-2 positive group, is often referred to as the secondary olfactory cortex, as it possesses direct connections to both entorhinal and piriform cortex^35^, as well as to the anterior olfactory nucleus^21,28^. In fact, in a recent functional connectivity study of the primary olfactory cortex, the orbitofrontal cortex was found to be connected to all four primary olfactory regions investigated (frontal and temporal piriform cortex, anterior olfactory nucleus and olfactory tubercle), possibly explaining why it is reliably activated even in basic and passive olfactory tasks^28^. Using the same olfactory connectivity maps, which overlap cortically in the orbitofrontal cortex, anterior cingulate cortex and insula, we found a more pronounced increase of diffusion metrics indicative of tissue damage in the SARS-CoV-2 group. The voxel-wise map of mean diffusivity pinpointed these longitudinal differences in the orbitofrontal and anterior cingulate cortex, as well as in the insula and the amygdala. The insula is not only directly connected to the primary olfactory cortex^21^, but is also considered to be the primary gustatory cortex. “Area G” (i.e., the dorsal part of the insula at the junction with the frontal and parietal operculum), in turn connects with the orbitofrontal cortex^36^. The vertex-wise and voxel-wise visualisation of both greater loss of grey matter and increase in mean diffusivity in the insula spatially correspond in particular to the area of consistent activation to all basic taste qualities^37^. Finally, the exploratory analysis revealed a more pronounced loss of grey matter in crus II, part of the cognitive, and olfactory-related lobule VII of the cerebellum^38^. These results are in line with previous post-infection PET findings showing, in more severe cases, FDG hypometabolism in the insula, orbitofrontal and anterior cingulate cortex, as well as lower grey matter volume in the insula and hippocampus^39,40^.

Early neurological signs in COVID-19 include hyposmia and hypogeusia, which appear to precede the onset of respiratory symptoms in the majority of affected patients^2,18,41^. In addition, a heavily-debated hypothesis has been that an entry point of SARS-CoV-2 to the central nervous system is via the olfactory mucosa, or the olfactory bulb^2,11,18^. (The coronavirus itself would not necessarily need to enter the central nervous system; anterograde degeneration from olfactory neurons might suffice to generate the pattern of abnormalities revealed in our longitudinal analyses.) The predominance observed in other studies of hyposmic and anosmic symptoms — whether caused directly by loss of olfactory neurons or by perturbation of supporting cells of the olfactory epithelium^15,20^ — could also, through repeated sensory deprivation, lead to loss of grey matter in these olfactory-related brain regions. Very focal reduction in grey matter in the orbitofrontal cortex and insula have been observed for instance in patients with severe olfactory dysfunction in a cross-sectional study of chronic rhinosinusitis^27^. A more extensive study of congenital and acquired (post-infectious, chronic inflammation due to rhinosinusitis, or idiopathic) olfactory loss also demonstrated an association between grey matter volume and olfactory function in the orbitofrontal cortex^17^. It also showed that duration of olfactory loss for those with acquired olfactory dysfunction, ranging from 0 to over 10 years, was related to more pronounced loss of grey matter in the gyrus rectus and orbitofrontal cortex^17^. On the other hand, it has been reported in a longitudinal study that patients with idiopathic olfactory loss had higher grey matter volume after undergoing olfactory training in various brain regions including the orbitofrontal cortex and gyrus rectus^42^. This raises the interesting possibility that the pattern of longitudinal abnormalities observed here in the limbic, olfactory brain regions of SARS-CoV-2 positive participants, if they are indeed related to olfactory dysfunction, might be attenuated over time if the infected participants go on to recover their sense of smell and taste. There is for instance some very preliminary evidence, in a few previously hospitalised COVID-19 patients, that brain hypometabolism becomes less pronounced when followed-up 6 months later, even if it does not entirely resolve^39,43^. In our much milder cohort, structural (as opposed to functional) changes might take longer and require larger numbers to be detected. When we tested whether time between infection and second brain scan had any relationship — positive, indicative of recovery, or negative, indicative of an ongoing degenerative process — with the grey matter loss or increase in diffusivity in the significant IDPs, we found no significant effect. This result is also possibly owing to the relatively small range in duration of infection at the time of this study, between 1 and 13 months for those 351 infected participants for whom we had a diagnosis date, and particularly with less than 20% of these participants having been infected for over 6 months (**Supplementary Fig. 3**). Another source of variability is that each individual in our cohort was infected between the months of March 2020 and April 2021, periods that saw various dominant strains of SARS-CoV-2. Of those 351 participants for whom we have a proxy date of infection, but no formal way of assessing the strain responsible for the infection, a small minority of the participants were likely infected with the original strain, and a majority with the variants of concern present in the UK from October 2020 onwards (predominantly Alpha, but also Beta and Gamma), while presumably very few participants, if any, were infected with the Delta variant, which only appeared in the UK in April 2021. Since the second scans have been acquired over a relatively short period in these positive participants (February-May 2021), SARS-CoV-2 strains and time between infection and second scan are also highly collinear. Additional follow-up of this cohort, not only increasing the number of cases infected for 6 months or longer, but also including cases infected by the Delta variant, would be particularly valuable in determining the longer-term effects of infection on these limbic structures, as well as possible differential effects between the various strains.

For various possible explanations for our longitudinal brain results, please see **Supplementary Discussion**.

Many of our results were found using imaging biomarkers of grey matter thickness or volume, which can be sensitive markers of a neurodegenerative process compared with other imaging modalities^44^, and are robust measurements that makes them ideal in a longitudinal setting^45^. In fact, the longitudinal differences between the SARS-CoV-2 positive and control groups, while significantly localised in a limbic olfactory and gustatory network, seemed also — at a lower level — to be generalised, as illustrated in the significant shift in the distribution of Z values over the entire cortical surface (**Supplementary Fig. 4**). This means that there is an overall stronger decrease of grey matter thickness across the entire cortex in the infected participants, but that this effect is particularly dominant in the olfactory system. A marked atrophy of fronto-parietal and temporal regions can also be seen when contrasting hospitalised and non-hospitalised cases, suggesting increased damage in the more moderate and severe cases, with an additional significant shift in Z values (**Supplementary Fig. 4**). The pattern of loss of grey matter in the hospitalised patients compared with the milder cases is in line with PET-FDG reports showing fronto-parietal and temporal decrease in glucose in hospitalised COVID-19 patients^16,43^.

The overlapping olfactory- and memory-related functions of the regions shown to alter significantly over time in SARS-CoV-2, including the parahippocampal gyrus/perirhinal cortex, entorhinal cortex and hippocampus in particular (**Supplementary Table 1**), raise the possibility that longer-term consequences of SARS-CoV-2 infection might in time contribute to Alzheimer’s disease or other forms of dementia^2^. This has led to the creation of an international consortium including the Alzheimer’s Association and representatives from more than 30 countries to investigate these questions^2^. In our sample of infected participants with mainly mild symptoms, we found no signs of memory impairment. However, these SARS-CoV-2 positive participants showed a worsening of executive function, taking a significantly greater time to complete trail A and particularly trail B of the Trail Making Test (**Fig. 3**). These findings remained significant after excluding the few hospitalised cases. While the UK Biobank version of the Trail Making Test is carried out online and unsupervised, there is good to very good agreement with the standard paper-and-pencil Trail Making Test on its measurements for completion of the two trails^46^, two measures known to be sensitive to detect impairment of executive function and attention, for instance in affective disorders and in schizophrenia^47,48^, and to discriminate mild cognitive impairment and dementia from healthy ageing^49^. In turn, the duration to complete the alphanumeric trail B was associated *post hoc* with the longitudinal changes in the cognitive part of the cerebellum, namely crus II, which is also specifically activated by olfactory tasks^38,50^. In line with this result, this particular part of the cerebellum has been recently shown to play a key role in the association with (and prediction of future) cognitive impairment in patients with stroke (subarachnoid haemorrhage)^51^. On the other hand, the parahippocampal gyrus and other memory-related regions did not show in our study any alteration on a functional level, i.e., any *post hoc* association with the selected cognitive tests. It remains to be determined whether the loss of grey matter and increased tissue damage seen in these specific limbic regions may in turn increase the risk for these participants of developing memory problems doi.org/10.1093/braincomms/fcab295, and perhaps dementia in the longer term^2,4,52^.

Limitations of this study include the lack of stratification of severity of the cases, beyond the information of whether they had been hospitalised (information on O2 saturation levels and details of treatment or hospital procedures is currently available on only a few participants); lack of clinical correlates as they are not currently available as part of the UK Biobank COVID-19-related links to health records (of particular relevance, potential hyposmic and hypogeusic symptoms and blood-based markers of inflammation); lack of identification of the specific SARS-CoV-2 strain having infected each participant; small number of participants from Asian, Black or other ethnic background other than White; and some of the cases and controls’ SARS-CoV-2 infection status being identified through antibody lateral flow test kits that have varied diagnostic accuracy^53^. However, it is worth noting that any potential misclassification of controls as positive cases (due to false positives in testing) and positive cases as controls (due to the absence of confirmed negative status and/or false negative tests) could only bias our results toward the null hypothesis of no difference between cases and controls. For those cases, no distinction is possible at present to determine whether a positive test is due to infection or vaccination, so potential cases identified only through lateral flow test in vaccinated participants were not included in this study. Information on the vaccination status (except for those identified through lateral flow test), and how both vaccination dates might interact with the date of infection, is also currently unavailable. While the two groups were not significantly different across major demographic and risk factor variables, we identified a subtle pattern of lower cognitive abilities in the participants who went on to be infected, but this could not explain away our longitudinal findings. The future positive cases also showed lower subcortical volume, and higher diffusion abnormalities at baseline compared with the controls, in brain regions not overlapping with our longitudinal results. One issue inherent to the recruitment strategy of UK Biobank, based on participants volunteering after being contacted at home for a possible re-imaging session, is the high number of mild cases. This can be seen however as a strength of this study: the majority of the brain imaging publications so far have focussed on moderate to severe cases of COVID-19^9^, hence there is a fundamental need for more information on the cerebral effects of the disease in its milder form. The UK Biobank COVID-19 re-imaging study is ongoing, and further information will eventually be made available. On the statistical approach, we have chosen a model form given strong priors of highly increased detrimental effects of SARS-CoV-2 and greater vulnerability of the brain with age. Using this objective model and rigorous statistical inference, we found significant and interpretable results. We have not tested all possible models for all possible IDPs; instead, we have focussed on one possible model drawn from independent, existing literature and found that it is “useful”, i.e., statistically significant. The model may not be optimal for every feature considered; in other words, this model might not be the most sensitive possible model for every IDP. However, the main expected outcome in such cases would be that we would fail to find significant results, and not that there would be any inflation of false positives. Finally, on the imaging side, our exploratory approach revealed significant longitudinal differences in the volume of the whole brainstem, but the UK Biobank scanning protocol and processing does not allow us to clarify which specific nuclei (e.g., potentially those that are key autonomic and respiratory control centres) might be involved, with the exception of the substantia nigra.

This is the first longitudinal imaging study comparing brain scans acquired from individuals before and after SARS-CoV-2 infection to those scans from a well-matched control group. It also is one of the largest COVID-19 brain imaging studies, with 785 participants including 401 individuals infected by SARS-CoV-2. Its unique design makes it possible to more confidently tease apart the pathogenic contribution associated, directly or indirectly, with the infection from pre-existing risk factors. By using automated, objective and quantitative methods, we uncovered a consistent spatial pattern of longitudinal abnormalities in limbic brain regions forming a mainly olfactory network. Whether these abnormal changes are the hallmark of the spread of the pathogenic effects, or of the virus itself in the brain, and whether these may prefigure a future vulnerability of the limbic system in particular, including memory, for these participants, remains to be investigated.

## Supporting information

Supplementary_Information_Guide

## Data Availability

Data can be accessed through the UK Biobank.

## Methods

### Ethics

Human subjects: UK Biobank has approval from the North West Multi-centre Research Ethics Committee (MREC) to obtain and disseminate data and samples from the participants (http://www.ukbiobank.ac.uk/ethics/), and these ethical regulations cover the work in this study. Written informed consent was obtained from all participants.

### Study Design

As part of the UK Biobank imaging study^57^, thousands of subjects had received brain scans before the start of the COVID-19 pandemic. Multimodal brain imaging data, collected at four sites with identical imaging hardware, scanner software and protocols, and passing quality controls, was obtained from 42,729 participants over the age of 45 years, and made available to researchers worldwide.

Before the COVID-19 pandemic, longitudinal (first- and second-timepoint scanning) had already begun in the UK Biobank imaging study, with about 3,000 participants returning for a second scan prior to scanning being paused in 2020 as a result of the pandemic. More recently, starting in February 2021, hundreds of UK Biobank participants who had already taken part in UK Biobank imaging before the pandemic were invited back for a second scan. This COVID-19 re-imaging study was set up to investigate the effects of SARS-CoV-2 infection by comparing imaging scans taken from participants before vs after infection.

The full list of inclusion criteria for the participants in this re-imaging study is as follows:

- had already attended an imaging assessment at one of the three imaging sites (the fourth opened just before the pandemic began),
- still lived within the catchment area of the clinic they attended for their first imaging assessment,
- had no incidental findings identified from their scans taken at the first imaging visit,
- had not withdrawn or died
- had a valid email and postal address,
- had high-quality scans from the first imaging visit,
- lived within 60 km of the clinic (extended to 75 km in Feb 2021), due to travel restrictions during the lockdown period.

(See for more details the online documentation: https://biobank.ndph.ox.ac.uk/showcase/ showcase/docs/casecontrol_covidimaging.pdf)

Amongst those, some participants were identified as having been infected with SARS-CoV-2 based on: (i) results of diagnostic antigen tests identified through linkage to health-related records, or (ii) their primary care (GP) data or hospital records, or (iii) results of two antibody tests.

The diagnostic antigen tests results data for England, Scotland, and Wales are made available on an ongoing basis by UK Biobank, and these data are provided by Public Health England (PHE), Public Health Scotland (PHS), and Secure Anonymised Information Linkage (SAIL, the databank from Wales), respectively. The data contain information on the date when the specimen was taken, origin (binary code for whether the patient was an inpatient when the specimen was taken), and result (binary code or positive and negative for SARS-CoV-2) of the tests along with encoded participant IDs (see biobank.ctsu.ox.ac.uk/crystal/ukb/docs/c19link_phe_sgss.pdf for further information on how regular updates of SARS-CoV-2 test results in England are made in UK Biobank).

For the primary care (GP) data, UK Biobank used this following set of codes: 1. TPP: Y213a, Y228d, XaLTE (if the event date is after January 1st, 2020), Y22b8, Y23f7, Y20d1, Y24ad, Y246f, Y269d, Y23f0, Y2a3b, Y2a15, Y212f, Y26a1, Y26b2, Y23e9, Y211c, Y23ec, Y2a3d; 2. EMIS: EMISNQCO303, 720293000, 720294006, 840535000, 840536004, 870361009, 870362002, 871552002, 871553007, 871555000, 871556004, 871557008, 871558003, 871559006, 871560001, 871562009, 1240581000000104, 1300721000000109, 1321541000000108, 1321551000000106, 1321661000000108, 1324881000000100. For the hospital records, the code used to identify positive SARS-CoV-2 cases was ICD10: U07.1. The dates of the records for both GP and hospital data were extracted along with the encoded participant IDs. In particular, the hospital records contain information on admission and discharge including episode start and end dates, primary and secondary causes for admission, critical care if applicable, and types of operations or procedures performed. We first identified hospitalised infected patients who had the ICD code U07.1 as a primary or secondary cause, and extracted information (e.g., admission/discharge date) relating to the episodes. OPCS-4 codes E85.1, E85.6, E85.2, and X52.9 were used to find out whether the patients were provided respiratory support during the episodes. No other information, for instance symptoms such as hyposmia or hypogeusia of particular relevance, were made available in these medical records.

Participants were also invited to take a home-based lateral flow (Fortress Fast COVID-19 Home test, Fortress Diagnostics and ABC-19TM Rapid Test, Abingdon Health) to detect the presence of SARS-CoV-2 antibodies. A second kit was sent to all participants who recorded an initial positive result and who had indicated they had not yet been vaccinated, in order to reduce the number of false positives.

Participants were classified as SARS-CoV-2 positive cases if they had a positive test record in any of the three data sources described above. Date of diagnosis (**Table 2**) was determined based on the information available in (i) and (ii). For participants with multiple positive test records, we took the earliest date as the date of diagnosis.

Controls were then selected by identifying, from the remaining previously imaged UK Biobank participants, those who had a negative antibody test result, as determined from the home-based lateral flow kits, and/or who had no record of confirmed or suspected COVID-19 from primary care, hospital records or diagnostic antigen test data. Controls were selected to match 1:1 to positive SARS-CoV-2 cases according to five criteria:

- sex
- ethnicity (white/non-white, as numbers were too low to allow for a finer distinction)
- date of birth (+/-6 months)
- location of first imaging assessment clinic
- date of first imaging assessment (+/-6 months).

Permission to use the UK Biobank Resource was obtained via Material Transfer Agreement (www.ukbiobank.ac.uk/media/p3zffurf/biobank-mta.pdf).

### Image Processing

For this work, we primarily used the IDPs generated by our team on behalf of UK Biobank, and made available to all researchers by UK Biobank^57,58^. The IDPs are summary measures, each describing a different aspect of brain structure or function, depending on what underlying imaging modality is used^57,58^.

The protocol includes three structural MRI scans (T1, T2 fluid attenuation inversion recovery (FLAIR) and susceptibility-weighted MRI), as well as diffusion MRI and resting and task functional MRI. T1 scans make it possible to derive global measures of brain and cerebrospinal fluid (CSF) volumes, as well as localised measures of grey matter volume and cortical thickness and area. The T2 FLAIR scan identifies differences that might be indicative of inflammation or tissue damage. Susceptibility-weighted MRI is sensitive to iron and myelin content. Diffusion MRI measurements give insight into the tissue microstructure integrity. Resting-state functional MRI is performed on an individual who is not engaged in any particular activity or task, and can provide indices related to the functional connectivity between brain regions^59^. Functional connectivity is intrinsically noisy when each region-pair connection is considered individually, so we focused here our analysis on 6 dimensionally-reduced functional connectivity networks^56^. We also did not consider *a priori* task-fMRI activation IDPs, as these have previously been found to have very low reproducibility and heritability^60^.

We used 1,524 existing UK Biobank IDPs, including: regional grey matter, brain and CSF volume, local cortical surface area, volume and thickness, cortical grey-white contrast, white matter hyperintensity volume, white matter microstructural measures such as fractional anisotropy and mean diffusivity, resting-state amplitude and dimensionally-reduced connectivity measures. In addition, we also generated 1,106 new IDPs, as described below.

We computed additional IDPs obtained using Quantitative Susceptibility Mapping (QSM), which has been recently added into our UK Biobank processing pipeline (doi.org/10.1101/ 2021.06.28.450248). Magnitude and phase data from the susceptibility-weighted MRI acquisitions were processed to provide quantitative measures reflecting clinically relevant tissue susceptibility properties. Median T2* was calculated within 17 subcortical structures (with their regions-of-interest (ROIs) estimated from the T1) as IDPs; 14 of these are the same subcortical regions already estimated by the core UK Biobank pipeline, and here we added 3 more subcortical ROIs: left and right substantia nigra^61^ and regions of white matter hyperintensities (lesions)^62^. Second, susceptibility-weighted MRI phase data were processed for QSM following a pipeline recently developed for UK Biobank^63^ (doi.org/10.1101/ 2021.05.19.21257316). QSM (CSF-referenced) IDPs were calculated in the same 17 subcortical structures as the T2* IDPs.

Additional IDPs were created via refined sub-segmentations of the hippocampus, amygdala and thalamus as implemented in FreeSurfer^64–67^. We extracted these ROI masks from the FreeSurfer processing and applied them to the T2* and diffusion images (diffusion tensor model: MD and FA; NODDI model: OD, ISOVF, ICVF) to generate additional subcortical IDPs.

Finally, we generated new IDPs tailored to the olfactory and gustatory systems, as described below.

### Hypothesis-driven approach

Based on prior expectations from animal models and *post mortem* findings, we chose to focus *a priori* our primary analyses on a subset of 332 regions-of-interest (297 of which passed the reproducibility thresholding; see Reproducibility section below) from the available 2,630 IDPs^21,22,36^; these correspond anatomically to the telencephalic primary and secondary connections of the olfactory and gustatory cortex. Briefly, these include the piriform cortex, parahippocampal gyrus, entorhinal cortex, amygdala, insula, frontal/parietal operculum, medial and lateral orbitofrontal cortex, hippocampus and basal ganglia. As no labelling of the piriform cortex exists in any of the atlases used in the UK Biobank imaging processing, we refined a previously published ROI of the piriform cortex (frontal and temporal), anterior olfactory nucleus and olfactory tubercle, by limiting it to the cortical ribbon of our UK Biobank T1-weighted standard space (https://github.com/zelanolab/primaryolfactorycortexparcellation^28^). We further used maps from the same study’s resting-state fMRI analysis of the functional connectivity of each of the four parts of this ROI (piriform frontal, piriform temporal, anterior olfactory nucleus and olfactory tubercle) to the rest of the brain, to generate four additional extended ROIs of the functionally-connected cortical and subcortical regions to these primary olfactory areas^28^. For this, we thresholded their connectivity t-value maps to keep only significant voxels (P_fwe_<0.05, with threshold-free cluster enhancement), and used the maps as weighted (and, separately, binarised) masks, to further extract grey matter volume (GM), T2* and diffusion values; this was done by: (i) regressing each of these maps into the GM, T2* or diffusion images in their respective native spaces, and separately, (ii) by binarising the maps and extracting mean and 95^th^ percentile values.

Additionally, masks for the left and right olfactory bulbs were generated by manually drawing a binary mask for the right olfactory bulb on an averaged template-space T2 FLAIR volume generated from 713 UK Biobank subjects, and mirroring this to obtain the mask for the left (having confirmed by visual inspection that symmetry in this region allowed for this to be effective). Both masks were then modulated by the T2 intensities in their respective ROIs, to account for partial volume effects, generating the final “label” maps with values ranging between 0-1. For the hypothalamus, we combined and refined ROIs from two previously published and publicly available atlases of a probabilistic hypothalamus map (https://neurovault.org/collections/3145/^61^) and hypothalamic subregions^68^. Both the probabilistic hypothalamus map and the binarised map obtained from fusing the 26 hypothalamic subregions were transformed to our standard space where the probabilistic map was then masked by the binarised map. We then extracted volume, and T2 mean and 95th percentile intensity measurements in subjects’ native spaces, using the olfactory bulb and hypothalamus maps (unthresholded and thresholded at 0.3, to reduce concerns about arbitrariness of threshold selection when re-binarising these very thin ROI masks after interpolation, a step which is unavoidable when transforming masks from one space to another). For the hypothalamus, we additionally extracted these metrics from T2* and diffusion images. All of the above preprocessing steps were defined and completed before any analyses of longitudinal change and case-control modelling.

The full list of 297 pre-determined and reproducible IDPs is available in **Supplementary Table 1.**

### Exploratory Approach

The full set of 2,630 IDPs described above were used for a more exploratory, inclusive analysis of SARS-CoV-2 infection effects on brain structure and function (see full list of reproducible IDPs in **Supplementary Table 1**).

### Statistical Modelling

The following modelling was applied in the same way to both the hypothesis-driven analyses of a subset of IDPs, and the all-IDPs exploratory analyses.

#### Outlier identification of the IDPs

All IDPs from all subjects were pooled for initial processing (at this stage blinded to the SARS-CoV-2 status of participants): 42,729 Scan 1 datasets (all pre-pandemic), 2,943 pre-pandemic Scan 2 datasets, and 890 Scan 2 datasets acquired after the beginning of the COVID-19 pandemic. Outlier values (individual IDPs from individual scanning sessions) were removed on the basis of being more extreme than 8 times the median absolute deviation from the median for a given IDP. Missing data for individual subjects and specific IDPs can therefore occur because of this step, or because the IDP was missing in the original data (e.g., because a given modality was not usable from a given participant). The fraction of total non-missing data, averaged across IDPs, is 0.93; all full results tables include the number of usable measurements for each IDP and for each statistical test. Importantly, there was no imbalance in amount of missing/outlier data between cases and controls: the number of cases with usable data, normalised by the total number of subjects with usable data, has the following percentiles across IDPs: percentiles [0, 1, 50, 99, 100] = 0.50, 0.50, 0.52, 0.52, 0.60, i.e., the median percentile is 0.52. From this analysis, the only 3 IDPs having this fraction greater than 0.53 were thalamic nuclei diffusion IDPs, which do not appear in any of our main results. These are also the only 3 IDPs with more than 24% missing/outlier data.

The IDPs from the 890 subjects imaged during the pandemic (SARS-CoV-2 positive cases and controls), from both timepoints, were then retained. Subjects were kept if at least the T1-weighted structural image was usable from both timepoints, resulting in IDPs at both timepoints (IDP1 and IDP2) from 785 subjects. The data were then pooled into a single dataset comprising 785 × 2 = 1,570 imaging sessions, and cross-sectional deconfounding, treating all scans equivalently, was carried out for head size, age, scanner table position, and image motion in the diffusion MRI data. This deconfounding is part of the data preprocessing, and is done at the level of individual scan sessions; hence, this needs to be carried out before combining all scans and subjects together in the main modelling. These imaging confound variables first had outlier removal applied as described above, though using a higher threshold of 15 times the median absolute deviation, because some important confounds have extremely non-Gaussian underlying distributions (e.g., MRI scanner table position), and we found that a threshold of 8 was too aggressive for these variables, for values that are perfectly acceptable when considered with the domain knowledge of these variables^58,69^.

#### Reproducibility of the IDPs

We then evaluated the scan-rescan reproducibility of IDPs, in order to discard IDPs that were not reasonably reproducible between scans. For each IDP, we correlated the IDP1 with IDP2 values, separately for cases and controls, resulting in two reproducibility measures (Pearson correlation r) for each IDP. The vectors of r values (one value per IDP) derived from cases and from controls were extremely highly correlated (r=0.98), showing that potential effects associated with infection are subtle compared with between-subject variability and IDP noise; hence, we averaged these cases and controls’ r values to give a single reproducibility measure for each IDP. From the initial set of 2,630 IDPs, the least reproducible IDPs (r<0.5) were discarded, leaving 2,048 IDPs. Finally, IDPs with high levels of missing data (usable values from fewer than 50 subjects) were discarded, leaving in total 2,047 IDPs.

#### Main longitudinal model, deconfounding

Despite initial case-control subject pairing (resulting in case and control groups being well matched), missing/outlier data potentially disrupted this exact paired matching, and thus we also included in the modelling confound variables derived from those factors originally used as pairing criteria: difference between the subjects’ ages at each of their two scans, the difference of the squares of the ages (to account for quadratic dependencies of IDPs on age), genetic sex, and ethnicity (white vs non-white).

Longitudinal IDP change (ΔIDP) was estimated by regressing IDP2 on IDP1^70^, as well as including in the regression the confound variables listed above.

The case-vs-control difference in this longitudinal IDP effect was modelled with a group difference regressor comprised of the case-vs-control binary variable modulated by a function of age at Scan 2 (Age2, a close proxy for age at infection for the SARS-CoV-2 group, with less than a year’s error). We chose to focus on an “objective” age model given the strong prior knowledge of a highly increased detrimental effect, at older ages, of SARS-CoV-2 infection and a greater vulnerability of the brain with age. The age dependence has been found to be exponential in studies of the effects of COVID-19 on hospitalisation and fatality rate^71,72^. We used here the exact age dependence found by a data-driven, meta-regression of 28 studies, with no free or subjectively-chosen parameters, to modulate the binary case-vs-control variable, based on age at Scan 2^72^.

The main case-vs-control group difference regressor of interest is therefore:

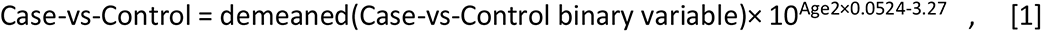

where the age-dependence constants are taken from the meta-regression analysis^72^ (see **Supplementary Analysis 5**). To ensure that the fitting of this term is not influenced by an effect that is common to controls and cases, we added a matching confound variable of 10^Age2×0.0524-3.27^, i.e., the same ageing term without the group-difference multiplier.

Our main model of interest therefore simply combines IDP1 and IDP2, the above group-difference model, and the confounds matrix:

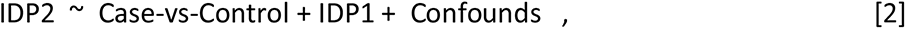

where the confounds matrix comprises the terms described above: Age2-Age1, Age2^2^-Age1^2^, ethnicity, sex, and 10^Age2×0.0524–3.27^.

By using a simple, single case-vs-control regressor for the main effect of interest, we optimise power for finding effects that follow this form, at the risk of sub-optimal power (sensitivity to finding true effects) if the effect does not follow this form.

Many forms for the case-vs-control model might be used. Possible models include: a binary regressor; single-regressors with age-modulated differences (such as the one primarily used here); more flexible models with multiple-regressors. Without testing a huge number of possible different models, one cannot make claims of absolute optimality. Nevertheless, our primary aim is not to prove model optimality, but to identify the effects of disease. To that aim, we have found statistically significant results with the simple model used here. Importantly, the exact choice of exponential model also held little bearing on our findings. Even opting for a binary case-vs-control regressor — i.e., without any age modulation — yielded similar, if a little weaker, primary results, consistent with our expectation of increased effects at higher ages (**Supplementary Table 5** and **Supplementary Analysis 5** for more details and discussion of non-modulated modelling results). **Supplementary Analysis 6** provides further model-fitting validity and robustness evaluations, including diagnostic residual scatter-plots and residual QQ plots, showing no obvious evidence of structured problems in model residuals or of model misspecifications.

The group-difference regressor is scaled to have average peak-peak height 1, so that the regression parameter from fitting Case-vs-Control can easily be converted into a percentage change measure, when normalised by the mean baseline value for a given IDP. For the main longitudinal modelling, this represents the average group difference in the longitudinal IDP change, and for the separate modelling of baseline IDPs only, this percentage reflects the average group difference in the baseline values. In addition to reporting % effects and associated standard errors, we also report the statistical significance as Z-statistics (Gaussianised regression model T-statistics), and P-values. Here, Z is more useful than T, because different IDPs have different patterns of missing data, and hence Z is more usefully comparable across IDPs. The regression inference automatically takes care of the degrees-of-freedom, including accounting for missing data and confound variables. For each IDP, any missing data is ignored (that subject is left out for that analysis). As part of the estimation of the longitudinal IDP changes, ΔIDP outliers (for each IDP, and each subject) were removed (set as missing), if they were more than 8 times the median absolute deviation from the median.

#### Multiple comparison correction

We used permutation testing to estimate family-wise-error P-values (Pfwe), i.e., correcting for the multiple comparisons across IDPs while accounting for the dependences among IDPs. We randomly permuted the residualised case-vs-control regressor relative to the residualised IDP2s, with 10,000 random permutations. At each permutation we computed the association Z value for each IDP, and recorded the maximum absolute value across all IDPs. By taking the absolute value, we corrected for the two-tailed nature of the test, i.e., we did not pre-assume the direction of any effect. After building up the null distribution of the maximum |Z| across IDPs, we then tested the original |Z| values against this distribution, to obtain family-wise error corrected P-values (P_fwe_), fully correcting for multiple comparisons across all IDPs. We also computed for each test the false discovery rate (FDR) at 5%, generating a threshold that can be applied to uncorrected P-values to determine their FDR significance.

We thus computed both FDR- and FWE-corrected inferences as two distinct measures of strength of evidence for a given effect. In this study, we primarily rely on FDR correction, which provides good power while controlling for multiple testing in a principled manner, but we wish to also indicate when a result additionally attains FWE significance. We therefore always specify the findings obtained using both correction methods in the Results section and the main and Supplementary Tables.

#### Group comparisons

In the rest of the manuscript, we refer to the main age-modulated-group comparison analysis (comparing IDPs at second timepoint controlling for IDPs at baseline) between SARS-CoV-2 positive cases and controls, as described above, as Model 1.

As secondary follow-up analyses, we also applied the same hypothesis-driven and exploratory approaches as described above, to compare non-hospitalised SARS-CoV-2 positive cases against controls (Model 2), and hospitalised patients against controls (Model 3). Separately, we also carried out the same analysis between hospitalised and non-hospitalised cases, adding as covariates three risk factors showing significant differences between these two SARS-CoV-2 groups (Model 4). For these secondary models (2-4), we again used age-modulated group-difference regressors as described above for Model 1. Power to detect effects in the two latter models, considering the hospitalised patients as a separate group, is of course considerably reduced, given the small number of hospitalised cases in this cohort.

For all 4 models, testing was carried out twice: first using the *a priori* focussed subset of IDPs identified for the hypothesis-driven analyses, and then using the full set of IDPs for the exploratory analyses. In both cases IDPs were identified as having significant group differences, corrected for multiple comparisons.

We thus carried out 8 imaging group comparison longitudinal analyses:

- the primary analysis comparing all cases vs all controls (Model 1), first in the set of olfactory-related IDPs *a priori* drawn, then in the exploratory set of IDPs
- secondary, ancillary analyses, using both hypothesis-driven and exploratory sets of IDPs:
- all non-hospitalised cases vs all controls (Model 2),
- all hospitalised cases vs all controls (Model 3),
- all hospitalised cases vs all non-hospitalised cases (Model 4).

### Cognitive analysis

While cognitive testing offers limited measurements of cognitive function in UK Biobank, we included in our ancillary cognitive analysis 10 variables sensitive to cognitive impairment. For this, we drew these variables using a data-driven approach based on identifying out-of-sample current and future dementia cases in UK Biobank, and comparing them to matched controls (**Supplementary Analysis 2**). The top most significant variables from this out-of-sample analysis were:

- three variables from the UK Biobank Trail Making Test: both durations to complete trails A and B, as well as the total number of errors made traversing trail B,
- one variable from the Symbol Digit Test: the number of symbol digit matches made correctly,
- one measure of reaction time: mean time to correctly identify matches at the card game “Snap”,
- one measure of reasoning: the “fluid intelligence” score,
- one measure of numeric memory: the maximum number of digits remembered correctly,
- three variables of the Pairs Matching test: numbers of correct and incorrect matches, and time to complete the test.

Based on these 10 variables from 6 different cognitive tests, we carried out two analyses: (i) the same group comparison between SARS-CoV-2 cases and controls of the longitudinal effect as described above, but substituting ΔIDP for ΔCOG, (ii) a *post hoc* regression analysis, in the SARS-CoV-2 group only, of the ΔCOG showing the most significant difference between cases and controls against the top 10 most significant ΔIDP for the hypothesis-driven approach and the top 10 for the exploratory approach. All results were evaluated for FWE and FDR significance, correcting for multiple comparisons across all cognitive, or IDP variables where applicable.

### Additional analyses

#### Baseline group comparisons

##### Risk factors

We compared the SARS-CoV-2 positive and control groups at baseline across common risk factors for infection and severity of disease: age, sex, blood pressure (systolic and diastolic), weight (including BMI, and waist-hip ratio), diabetes, smoking, alcohol consumption and socio-economic status (via the Townsend deprivation index). For this, we used the ‘Last Observation Carried Forward’ (LOCF) imputation method, for which we considered all the values available closest to the Scan 1 visit (for the majority of the values, these were available from the same visit, on the same day that Scan 1 was acquired); we also tested that there was no difference between SARS-CoV-2 and control groups in the distribution of the visits used to collect the LOCF values.

##### All other non-imaging phenotypes

We also examined whether the SARS-CoV-2 and control groups differed at baseline across all non-imaging phenotypes (lifestyle, environmental, health-related, dietary), across all UK Biobank visits. We assessed the 6,301 pre-Scan2 non-imaging phenotypes having at least 3% of values as being distinct from the majority value, and results were corrected for multiple comparisons using FDR and FWE (i.e., where relevant we refer to both in Results).

##### IDPs

To complement our longitudinal analyses, we carried out a baseline-only (and, separately, second timepoint only) cross-sectional group comparison between SARS-CoV-2 cases and controls, across all 2,047 IDPs, correcting for multiple comparisons across all IDPs using the same permutation-testing procedure as described above.

In particular, this approach is of interest to test whether brain regions showing significant longitudinal changes demonstrate initial differences, pre-existing *before* the infection, between the two groups.

##### Cognition

We finally assessed whether the two groups differed at baseline in their cognition, based on the results from the 10 variables from 6 different cognitive tests preselected above, correcting for multiple comparisons across cognitive variables.

#### Lateralised effects

As a *post hoc* analysis, we explored whether the longitudinal effects observed in grey matter thickness was lateralised, by subtracting right ΔIDP from the corresponding left ΔIDP, for: (i) all ΔIDPs of grey matter thickness showing significance in the main case-control analyses (across the hypothesis-driven and exploratory approaches), within the SARS-CoV-2 group only (to avoid circularity); (ii) all ΔIDPs of grey matter thickness across the entire cortex (151 pairs of left-right matched IDPs), and testing for associations between the left-right difference and the case-vs-control age modulated regressor. Results were corrected for multiple comparisons using FDR and FWE.

#### Effect of time of SARS-CoV-2 infection

For 351 SARS-CoV-2 positive participants who had a date available for infection (hence, in effect excluding those identified through antibody lateral flow tests), we further looked *post hoc* at the possible effect of time interval between infection and second brain scan (acquired post-infection) on the significant IDP from our hypothesis-driven approach, to evaluate whether a longer interval might mean either a reduced loss of grey matter through potential progressive recovery of sensory inputs (olfaction), or greater loss as a function of a longer, ongoing degenerative process.

#### Impact of non-imaging factors

We ran an additional analysis to test whether any non-imaging variables measured before SARS-CoV-2 infection might explain *post hoc* the longitudinal effects observed in our significant IDPs. We considered non-imaging variables with at least 50% non-missing data in the participants (n=6,301). We included individually each of these variables as additional confound for a repeat of the original Model 1 regression tests for those IDPs found to show significant longitudinal differences between the two groups, for both hypothesis-driven and exploratory approaches. If the strength of the original association was reduced by more than 25%, based on the regression Z-statistics, we considered a non-imaging variable to potentially explain the IDP-infection association. See **Supplementary Analysis 7** for further details.

## Data and code availability statement

All source data is available (upon data access application) from UK Biobank. Please see https://www.fmrib.ox.ac.uk/ukbiobank/covid/ for analysis code from this study, as well as at: doi.org/10.5281/zenodo.5903258.

## Acknowledgements

This work was primarily supported by a Wellcome Trust Collaborative Award 215573/Z/19/Z. K.L.M. was supported by a Wellcome Trust Senior Research Fellowship 202788/Z/16/Z. The Wellcome Centre for Integrative Neuroimaging (WIN FMRIB) is supported by centre funding from the Wellcome Trust (203139/Z/16/Z). S.L. was supported by the Rina M. Bidin Foundation Fellowship in Research of Brain Treatment and the Pacific Parkinson’s Research Institute. P.K. was supported by the UK Research and Innovation (MR/S034978/1). P.M.M. acknowledges generous personal and research support from the Edmond J. Safra Foundation and Lily Safra, an NIHR Senior Investigator Award, the UK Dementia Research Institute and the NIHR Biomedical Research Centre at Imperial College London. This research has been conducted in part using the UK Biobank Resource under Application Number 8107. We are grateful to UK Biobank for making the data available, and to all UK Biobank study participants, who generously donated their time to make this resource possible. We would like to thank Profs. Bruce Fischl and Doug Greve on guidance with the FreeSurfer analyses. Analysis was carried out at the Oxford Biomedical Research Computing (BMRC) facility. BMRC is a joint development between the Wellcome Centre for Human Genetics and the Big Data Institute, supported by Health Data Research UK and the NIHR Oxford Biomedical Research Centre.

## Author contributions

G.D., S.L., F. A.-A., C.A., C.W., P. McC., F.L., J.L.R.A., L.G., E.D., S.J., K.L.M. and S.M.S. created, extracted and organised the imaging and clinical data. S.M.S. carried out the imaging analyses. B.T., A.M.W., and T.E.N co-supervised the statistical analyses. R.C., P.M.M, N.A., K.L.M. and S.M.S. contributed to the creation of the UK Biobank COVID-19 re-imaging project. G.D., P.K., K.L.M. and S.M.S conceived the brain imaging study. G.D. interpreted the results. G.D. and S.M.S. wrote the paper. All co-authors revised the paper.

## Extended Data

### Extended Data Figures

**Extended Data Fig. 1:**
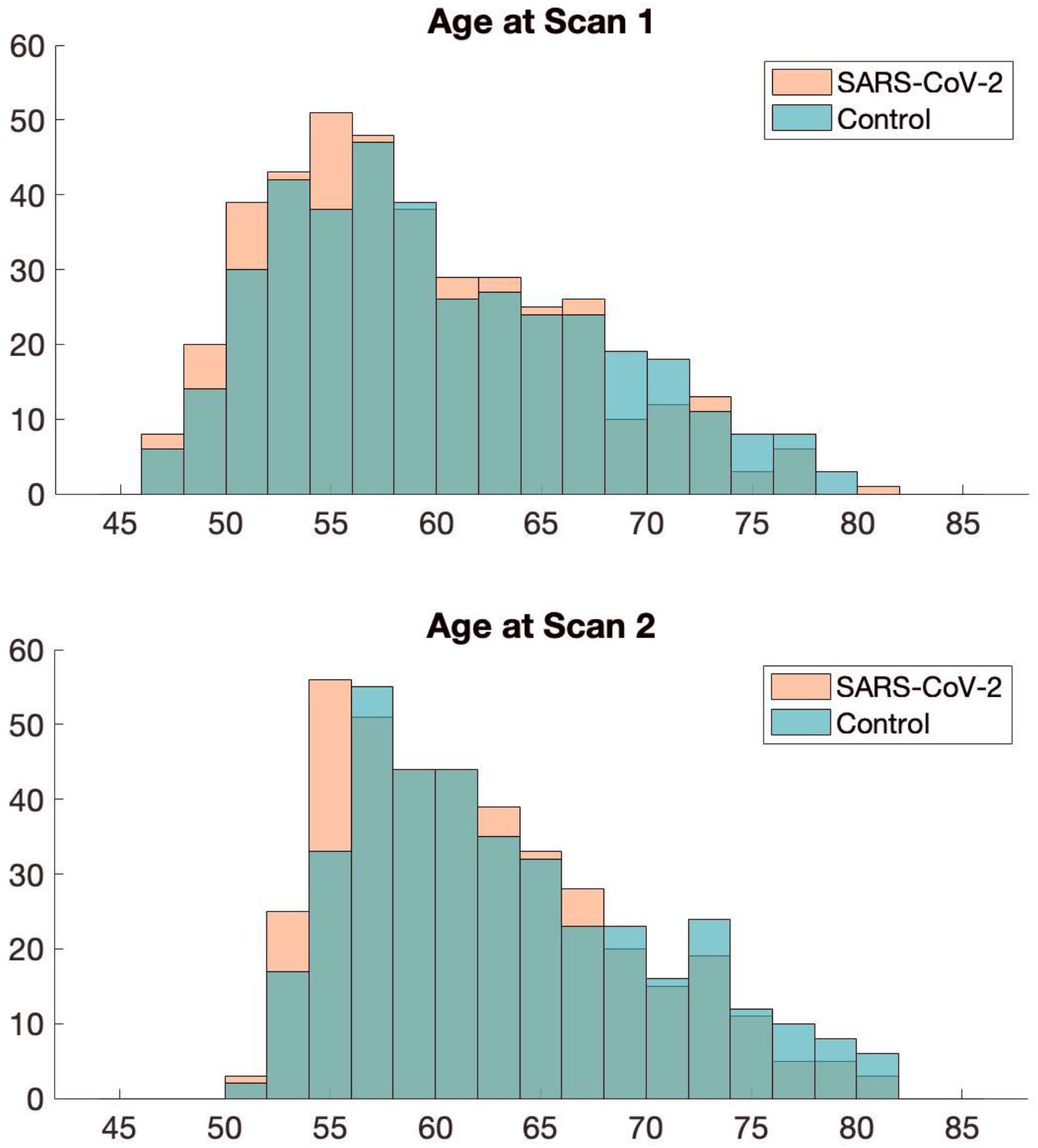
Age distributions for SARS-CoV-2 positive participants and controls at each timepoint do not differ significantly. Two-sample Kolmogorov-Smirnov was used to compute the P values for age comparisons, since age for each group was not normally distributed (Lilliefors P = 1e-03 for each group, and both age at Scan 1 or Scan 2). This showed no significant difference in age distribution between SARS-CoV-2 participants and controls at Scan 1: P = 0.15 or at Scan 2: P = 0.08.

**Extended Data Fig. 2:**
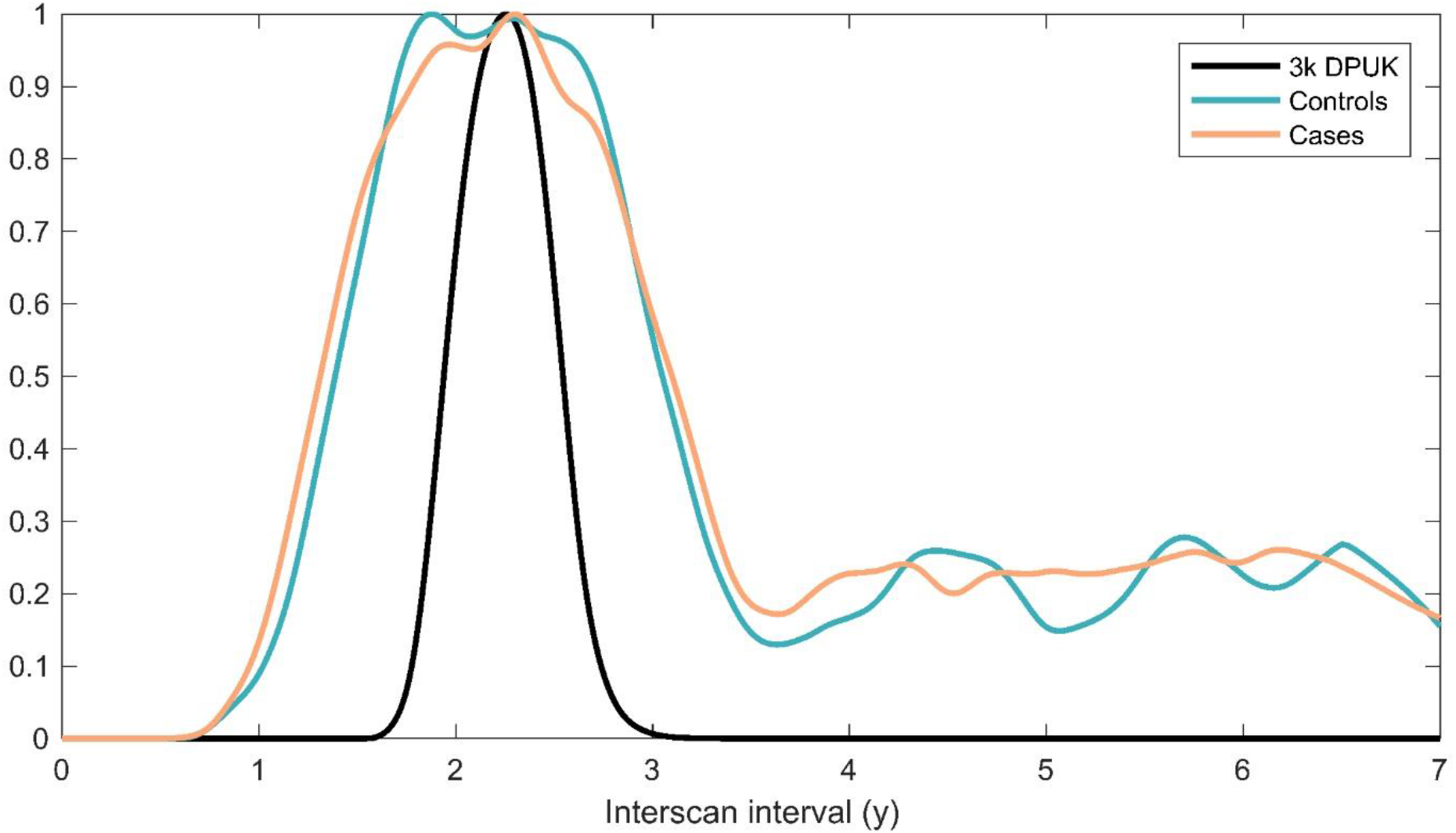
Histograms showing the well-matched distributions of Scan 1 - Scan 2 intervals for case and control groups. The below IDP reproducibility **Extended Data Fig. 3** shows, for comparison against the cases and controls, reproducibility from around 3,000 (2,943) UK Biobank participants who had returned for a second scan prior to the pandemic; hence we also show here the interscan intervals for this “3k” group, with tighter control over this interval (we have normalised each of those 3 groups to have a peak of 1, to make the relative comparison easier).

**Extended Data Fig. 3:**
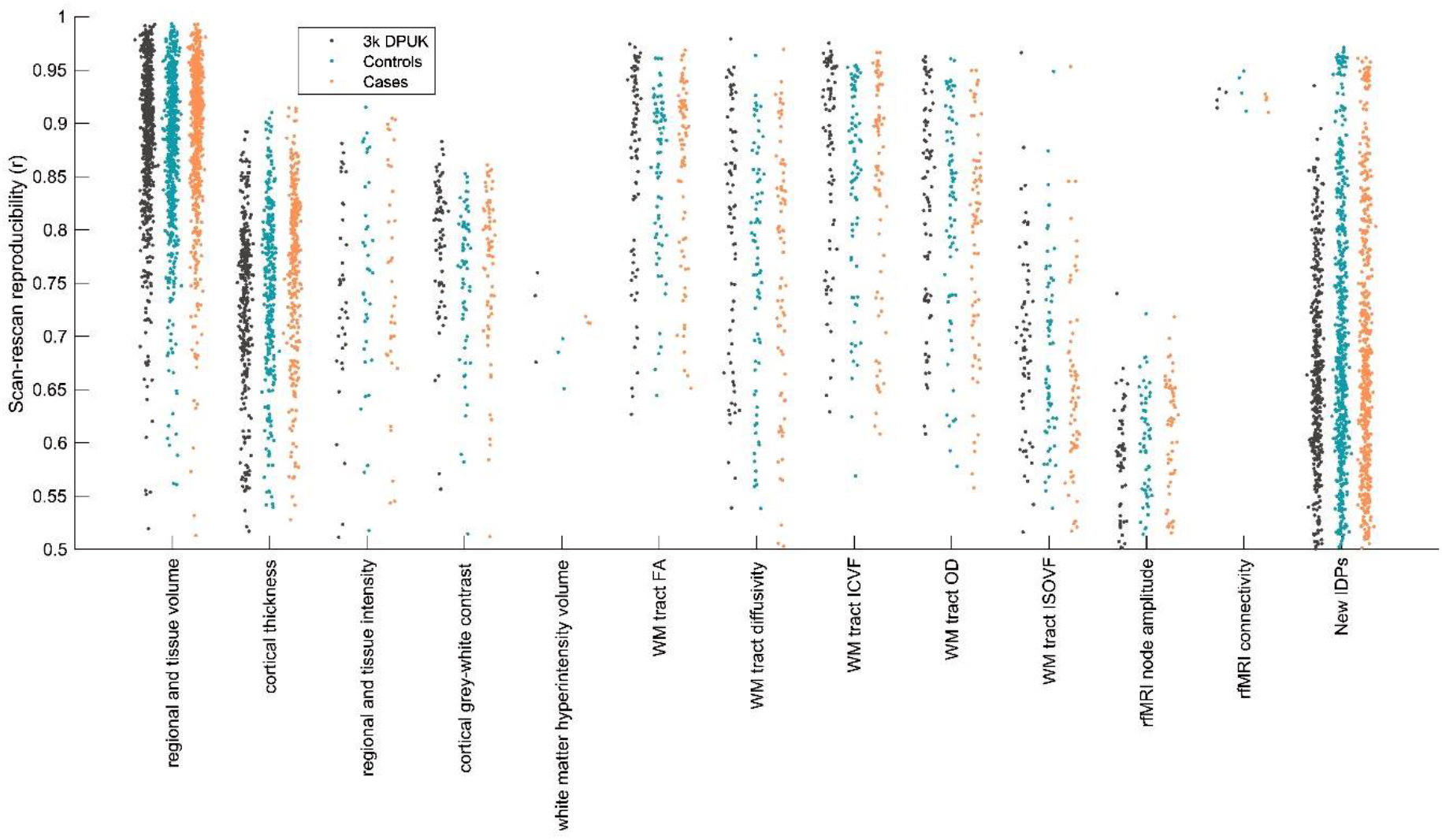
Scan-rescan reproducibility for all 2,047 IDPs used in the main modelling. Each dot represents a single IDP, arranged into different classes of IDPs. For each IDP, the vector of values for each subject (i.e., 785×1 vector) from the first scan was correlated with the equivalent vector of IDP values from the second scan. The y axis shows the resulting correlation coefficient. These calculations are made separately for the pre-pandemic scan- rescan datasets (“3k DPUK”), and for cases and controls, demonstrating highly similar distributions within each IDP class for all 3 subject groups.

**Extended Data Fig. 4:**
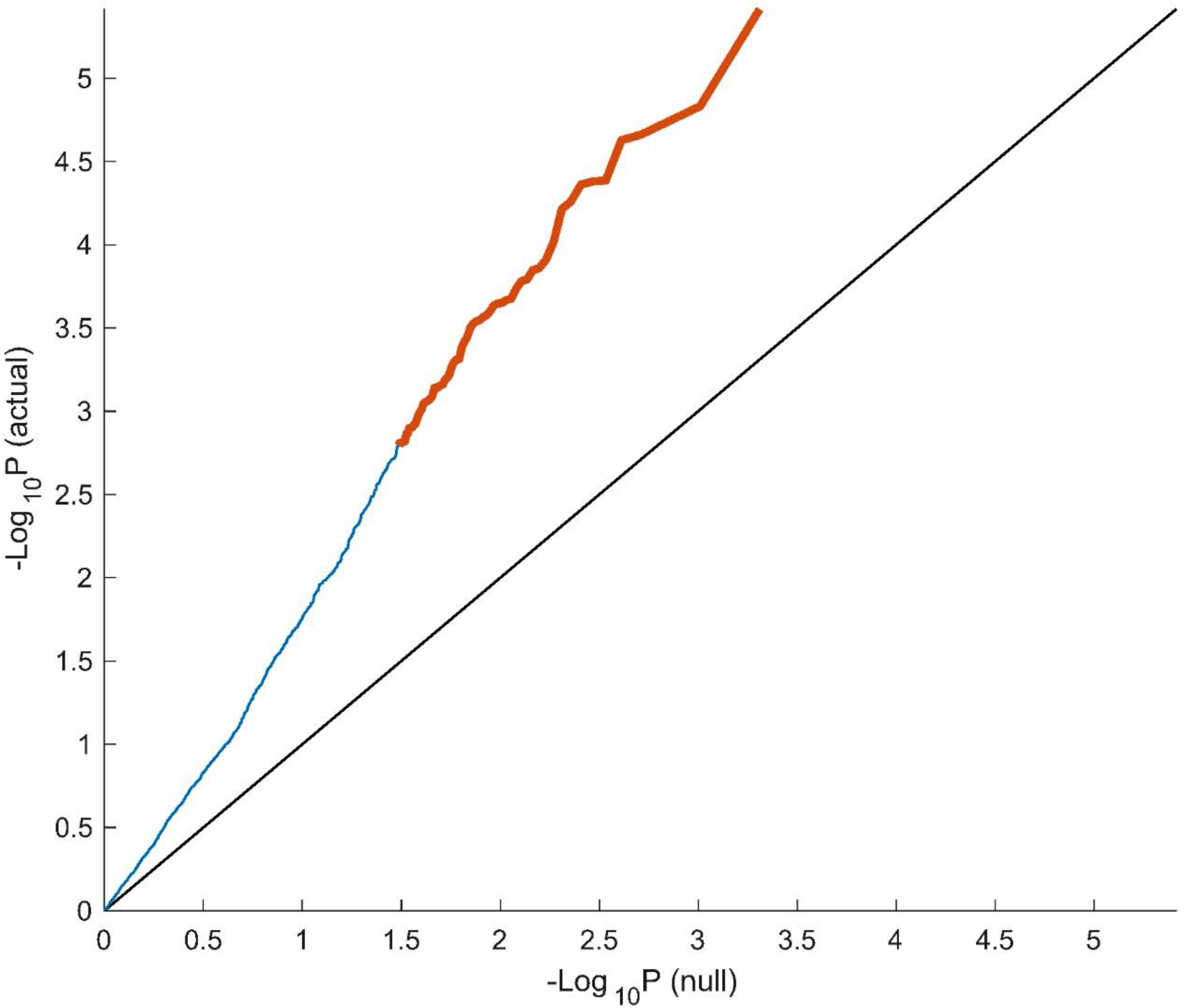
QQ plot for -Log_10_(P_uncorrected_) against the theoretical null distribution. The black line at y=x shows the expected plot if no effects were present in the data. Orange points reflect ΔIDPs where the case-control effect passes FDR significance, and blue reflects those that do not.

**Extended Data Fig. 5:**
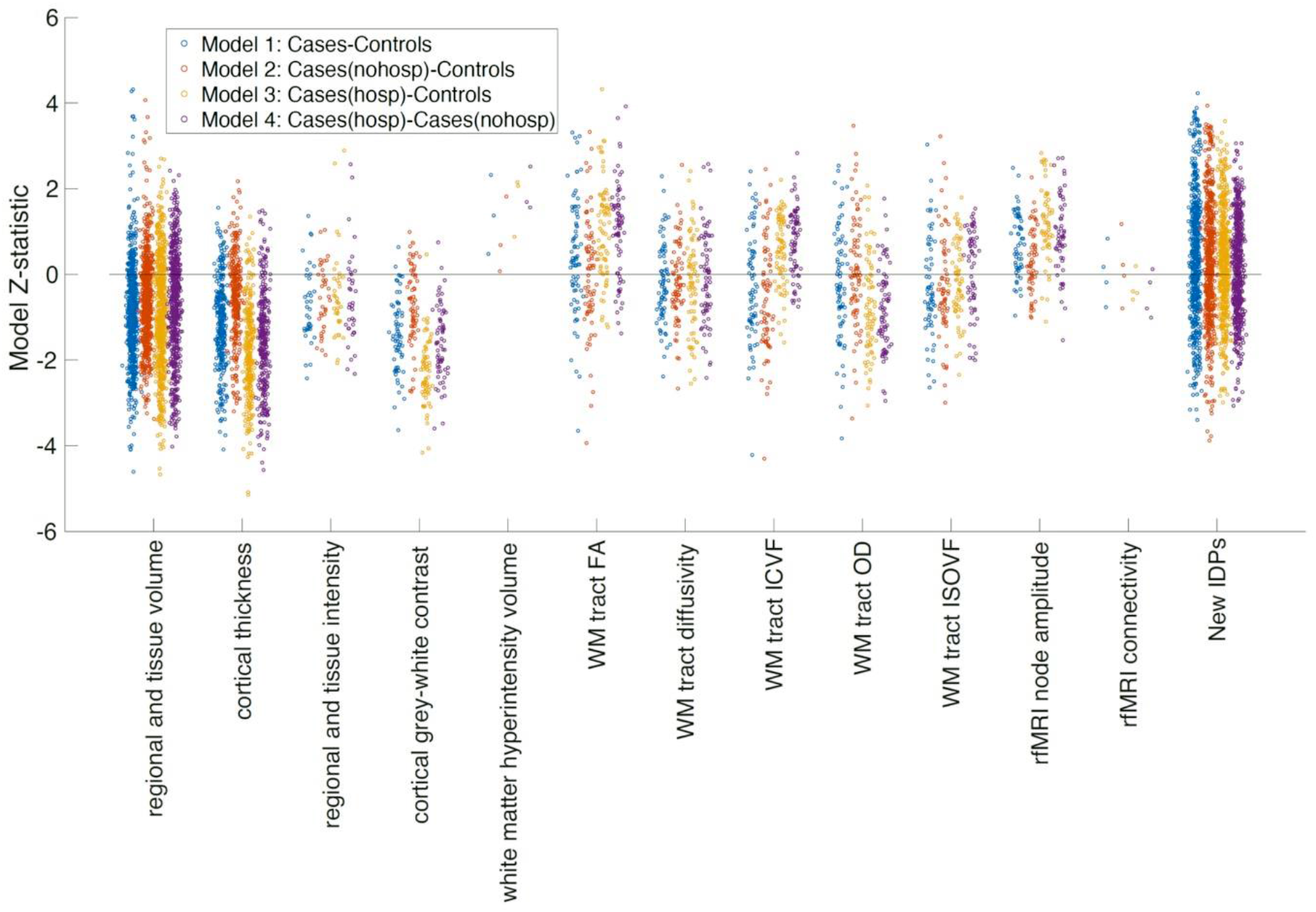
Model Z-statistics (one point per IDP, arranged in IDP classes) for the 4 main models. Note that these are model Z-statistics, not raw effect size. Some IDP classes (e.g., cortical thickness and grey-white intensity contrast) show consistent group-difference effect directions across most IDPs (i.e., different brain regions), and all 4 models.

**Extended Data Fig. 6:**
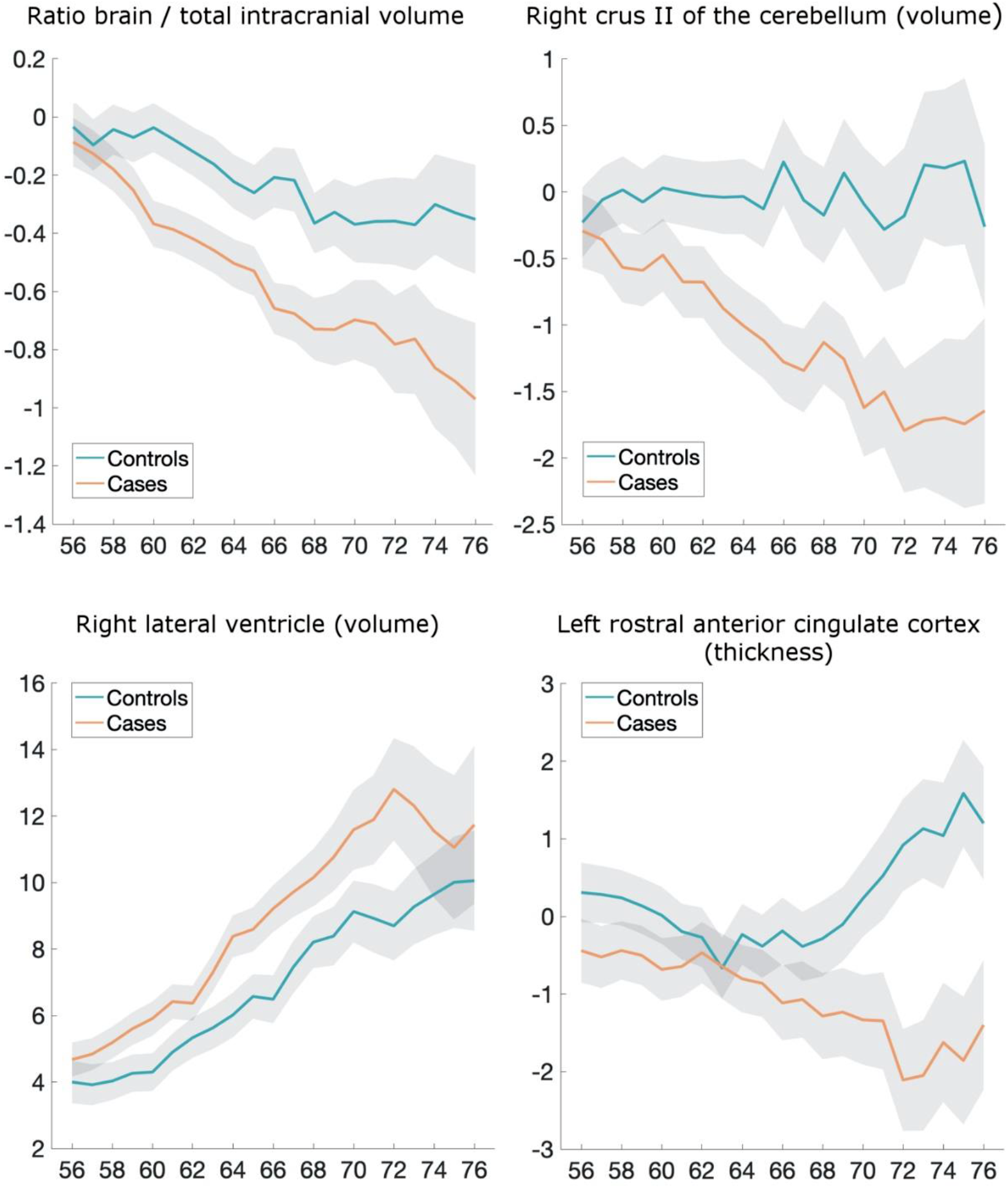
Examples of some of the most significant longitudinal group comparison results - exploratory approach. Four amongst the top IDPs consistently showing longitudinal differences between SARS-CoV-2 cases and controls. All demonstrate either a greater reduction in local or global brain thickness and volume, or an increase in CSF volume. For each four IDP are the percentage changes with age for the two groups, obtained by normalising ΔIDP using as baseline the values for the corresponding IDPs across the 785 scans (created using a 10-year sliding window across cases and controls, with standard errors in grey). The somewhat counterintuitive increase in thickness in the rostral anterior cingulate cortex in older controls has been previously consistently reported in studies of ageing, together with that of the orbitofrontal cortex^54,55^.

## References

1 Paterson, R. W. et al. The emerging spectrum of COVID-19 neurology: clinical, radiological and laboratory findings. Brain 143, 3104–3120, doi:10.1093/brain/awaa240 (2020).

2 de Erausquin, G. A. et al. The chronic neuropsychiatric sequelae of COVID-19: The need for a prospective study of viral impact on brain functioning. Alzheimers Dement, doi:10.1002/alz.12255 (2021).

3 Yang, A. C. et al. Dysregulation of brain and choroid plexus cell types in severe COVID-19. Nature, doi:10.1038/s41586-021-03710-0 (2021).

4 Deleidi, M. & Isacson, O. Viral and inflammatory triggers of neurodegenerative diseases. Sci Transl Med 4, 121ps123, doi:10.1126/scitranslmed.3003492 (2012).

5 Butowt, R., Meunier, N., Bryche, B. & von Bartheld, C. S. The olfactory nerve is not a likely route to brain infection in COVID-19: a critical review of data from humans and animal models. Acta Neuropathol 141, 809–822, doi:10.1007/s00401-021-02314-2 (2021).

6 Taquet, M., Geddes, J. R., Husain, M., Luciano, S. & Harrison, P. J. 6-month neurological and psychiatric outcomes in 236 379 survivors of COVID-19: a retrospective cohort study using electronic health records. Lancet Psychiatry 8, 416–427, doi:10.1016/S2215-0366(21)00084-5 (2021).

7 Taquet, M., Luciano, S., Geddes, J. R. & Harrison, P. J. Bidirectional associations between COVID-19 and psychiatric disorder: retrospective cohort studies of 62 354 COVID-19 cases in the USA. Lancet Psychiatry 8, 130–140, doi:10.1016/S2215-0366(20)30462-4 (2021).

8 Helms, J. et al. Neurologic Features in Severe SARS-CoV-2 Infection. N Engl J Med 382, 2268–2270, doi:10.1056/NEJMc2008597 (2020).

9 Manca, R., De Marco, M., Ince, P. G. & Venneri, A. Heterogeneity in Regional Damage Detected by Neuroimaging and Neuropathological Studies in Older Adults With COVID-19: A Cognitive-Neuroscience Systematic Review to Inform the Long-Term Impact of the Virus on Neurocognitive Trajectories. Front Aging Neurosci 13, 646908, doi:10.3389/fnagi.2021.646908 (2021).

10 Mukerji, S. S. & Solomon, I. H. What can we learn from brain autopsies in COVID-19? Neuroscience letters 742, 135528, doi:10.1016/j.neulet.2020.135528 (2021).

11 Meinhardt, J. et al. Olfactory transmucosal SARS-CoV-2 invasion as a port of central nervous system entry in individuals with COVID-19. Nat Neurosci 24, 168–175, doi:10.1038/s41593-020-00758-5 (2021).

12 Puelles, V. G. et al. Multiorgan and Renal Tropism of SARS-CoV-2. N Engl J Med 383, 590–592, doi:10.1056/NEJMc2011400 (2020).

13 Matschke, J. et al. Neuropathology of patients with COVID-19 in Germany: a post-mortem case series. Lancet Neurol 19, 919–929, doi:10.1016/S1474-4422(20)30308-2 (2020).

14 Lechien, J. R. et al. Olfactory and gustatory dysfunctions as a clinical presentation of mild-to-moderate forms of the coronavirus disease (COVID-19): a multicenter European study. Eur Arch Otorhinolaryngol 277, 2251–2261, doi:10.1007/s00405-020-05965-1 (2020).

15 Cooper, K. W. et al. COVID-19 and the Chemical Senses: Supporting Players Take Center Stage. Neuron 107, 219–233, doi:10.1016/j.neuron.2020.06.032 (2020).

16 Hosp, J. A. et al. Cognitive impairment and altered cerebral glucose metabolism in the subacute stage of COVID-19. Brain 144, 1263–1276, doi:10.1093/brain/awab009 (2021).

17 Postma, E. M., Smeets, P. A. M., Boek, W. M. & Boesveldt, S. Investigating morphological changes in the brain in relation to etiology and duration of olfactory dysfunction with voxel-based morphometry. Sci Rep 11, 12704, doi:10.1038/s41598-021-92224-w (2021).

18 Butowt, R. & Bilinska, K. SARS-CoV-2: Olfaction, Brain Infection, and the Urgent Need for Clinical Samples Allowing Earlier Virus Detection. ACS Chem Neurosci 11, 1200–1203, doi:10.1021/acschemneuro.0c00172 (2020).

19 Netland, J., Meyerholz, D. K., Moore, S., Cassell, M. & Perlman, S. Severe acute respiratory syndrome coronavirus infection causes neuronal death in the absence of encephalitis in mice transgenic for human ACE2. J Virol 82, 7264–7275, doi:10.1128/JVI.00737-08 (2008).

20 Brann, D. H. et al. Non-neuronal expression of SARS-CoV-2 entry genes in the olfactory system suggests mechanisms underlying COVID-19-associated anosmia. Sci Adv 6, doi:10.1126/sciadv.abc5801 (2020).

21 Carmichael, S. T., Clugnet, M. C. & Price, J. L. Central olfactory connections in the macaque monkey. The Journal of comparative neurology 346, 403–434, doi:10.1002/cne.903460306 (1994).

22 Palouzier-Paulignan, B. et al. Olfaction under metabolic influences. Chem Senses 37, 769–797, doi:10.1093/chemse/bjs059 (2012).

23 Guedj, E. et al. (18)F-FDG brain PET hypometabolism in post-SARS-CoV-2 infection: substrate for persistent/delayed disorders? Eur J Nucl Med Mol Imaging 48, 592–595, doi:10.1007/s00259-020-04973-x (2021).

24 Raman, B. et al. Medium-term effects of SARS-CoV-2 infection on multiple vital organs, exercise capacity, cognition, quality of life and mental health, post-hospital discharge. EClinicalMedicine 31, 100683, doi:10.1016/j.eclinm.2020.100683 (2021).

25 Griffanti, L. et al. Adapting the UK Biobank Brain Imaging Protocol and Analysis Pipeline for the C-MORE Multi-Organ Study of COVID-19 Survivors. Front Neurol 12, 753284, doi:10.3389/fneur.2021.753284 (2021).

26 Reichert, J. L. & Schopf, V. Olfactory Loss and Regain: Lessons for Neuroplasticity. Neuroscientist 24, 22–35, doi:10.1177/1073858417703910 (2018).

27 Han, P. et al. Olfactory brain gray matter volume reduction in patients with chronic rhinosinusitis. Int Forum Allergy Rhinol 7, 551–556, doi:10.1002/alr.21922 (2017).

28 Zhou, G., Lane, G., Cooper, S. L., Kahnt, T. & Zelano, C. Characterizing functional pathways of the human olfactory system. Elife 8, doi:10.7554/eLife.47177 (2019).

29 Fraser, M. A. et al. Longitudinal trajectories of hippocampal volume in middle to older age community dwelling individuals. Neurobiol Aging 97, 97–105, doi:10.1016/j.neurobiolaging.2020.10.011 (2021).

30 Ronnlund, M., Nyberg, L., Backman, L. & Nilsson, L. G. Stability, growth, and decline in adult life span development of declarative memory: cross-sectional and longitudinal data from a population-based study. Psychol Aging 20, 3–18, doi:10.1037/0882-7974.20.1.3 (2005).

31 Vidal-Pineiro, D. et al. Individual variations in ‘brain age’ relate to early-life factors more than to longitudinal brain change. Elife 10, doi:10.7554/eLife.69995 (2021).

32 Diana, R. A., Yonelinas, A. P. & Ranganath, C. Imaging recollection and familiarity in the medial temporal lobe: a three-component model. Trends in cognitive sciences 11, 379–386, doi:10.1016/j.tics.2007.08.001 (2007).

33 Staresina, B. P., Duncan, K. D. & Davachi, L. Perirhinal and parahippocampal cortices differentially contribute to later recollection of object- and scene-related event details. J Neurosci 31, 8739–8747, doi:10.1523/JNEUROSCI.4978-10.2011 (2011).

34 Naya, Y. & Suzuki, W. A. Integrating what and when across the primate medial temporal lobe. Science (New York, N.Y 333, 773–776, doi:10.1126/science.1206773 (2011).

35 Doty, R. L. Olfaction: Smell of Change in the Air. Cerebrum 2017 (2017).

36 Avery, J. A. et al. Taste Quality Representation in the Human Brain. J Neurosci 40, 1042–1052, doi:10.1523/JNEUROSCI.1751-19.2019 (2020).

37 Chikazoe, J., Lee, D. H., Kriegeskorte, N. & Anderson, A. K. Distinct representations of basic taste qualities in human gustatory cortex. Nat Commun 10, 1048, doi:10.1038/s41467-019-08857-z (2019).

38 Ferdon, S. & Murphy, C. The cerebellum and olfaction in the aging brain: a functional magnetic resonance imaging study. NeuroImage 20, 12–21, doi:10.1016/s1053-8119(03)00276-3 (2003).

39 Kas, A. et al. The cerebral network of COVID-19-related encephalopathy: a longitudinal voxel-based 18F-FDG-PET study. Eur J Nucl Med Mol Imaging 48, 2543–2557, doi:10.1007/s00259-020-05178-y (2021).

40 Qin, Y. et al. Long-term microstructure and cerebral blood flow changes in patients recovered from COVID-19 without neurological manifestations. J Clin Invest 131, doi:10.1172/JCI147329 (2021).

41 Tsai, S. T., Lu, M. K., San, S. & Tsai, C. H. The Neurologic Manifestations of Coronavirus Disease 2019 Pandemic: A Systemic Review. Front Neurol 11, 498, doi:10.3389/fneur.2020.00498 (2020).

42 Han, P., Musch, M., Abolmaali, N. & Hummel, T. Improved Odor Identification Ability and Increased Regional Gray Matter Volume After Olfactory Training in Patients With Idiopathic Olfactory Loss. Iperception 12, 20416695211005811, doi:10.1177/20416695211005811 (2021).

43 Blazhenets, G. et al. Slow but Evident Recovery from Neocortical Dysfunction and Cognitive Impairment in a Series of Chronic COVID-19 Patients. J Nucl Med 62, 910–915, doi:10.2967/jnumed.121.262128 (2021).

44 Douaud, G. et al. DTI measures in crossing-fibre areas: increased diffusion anisotropy reveals early white matter alteration in MCI and mild Alzheimer’s disease. NeuroImage 55, 880–890, doi:10.1016/j.neuroimage.2010.12.008 (2011).

45 Douaud, G. et al. Preventing Alzheimer’s disease-related gray matter atrophy by B-vitamin treatment. Proceedings of the National Academy of Sciences of the United States of America 110, 9523–9528, doi:10.1073/pnas.1301816110 (2013).

46 Fawns-Ritchie, C. & Deary, I. J. Reliability and validity of the UK Biobank cognitive tests. PLoS One 15, e0231627, doi:10.1371/journal.pone.0231627 (2020).

47 Mahlberg, R., Adli, M., Bschor, T. & Kienast, T. Age effects on trail making test during acute depressive and manic episode. Int J Neurosci 118, 1347–1356, doi:10.1080/00207450601059452 (2008).

48 Mahurin, R. K. et al. Trail making test errors and executive function in schizophrenia and depression. Clin Neuropsychol 20, 271–288, doi:10.1080/13854040590947498 (2006).

49 Ashendorf, L. et al. Trail Making Test errors in normal aging, mild cognitive impairment, and dementia. Arch Clin Neuropsychol 23, 129–137, doi:10.1016/j.acn.2007.11.005 (2008).

50 Sobel, N. et al. Odorant-induced and sniff-induced activation in the cerebellum of the human. J Neurosci 18, 8990–9001 (1998).

51 Rowland, M. J. et al. Early brain injury and cognitive impairment after aneurysmal subarachnoid haemorrhage. Sci Rep 11, 23245, doi:10.1038/s41598-021-02539-x (2021).

52 Heneka, M. T., Kummer, M. P. & Latz, E. Innate immune activation in neurodegenerative disease. Nat Rev Immunol 14, 463–477, doi:10.1038/nri3705 (2014).

53 Deeks, J. J. et al. Antibody tests for identification of current and past infection with SARS-CoV-2. Cochrane Database Syst Rev 6, CD013652, doi:10.1002/14651858.CD013652 (2020).

54 Salat, D. H. et al. Thinning of the cerebral cortex in aging. Cerebral cortex 14, 721–730, doi:10.1093/cercor/bhh032 (2004).

55 Zhao, L. et al. Age-Related Differences in Brain Morphology and the Modifiers in Middle-Aged and Older Adults. Cereb Cortex 29, 4169–4193, doi:10.1093/cercor/bhy300 (2019).

56 Elliott, L. T. et al. Genome-wide association studies of brain imaging phenotypes in UK Biobank. Nature 562, 210–216, doi:10.1038/s41586-018-0571-7 (2018).

## References

57 Miller, K. L. et al. Multimodal population brain imaging in the UK Biobank prospective epidemiological study. Nature neuroscience 19, 1523–1536, doi:10.1038/nn.4393 (2016).

58 Alfaro-Almagro, F. et al. Image processing and Quality Control for the first 10,000 brain imaging datasets from UK Biobank. NeuroImage 166, 400–424, doi:10.1016/j.neuroimage.2017.10.034 (2018).

59 Littlejohns, T. J. et al. The UK Biobank imaging enhancement of 100,000 participants: rationale, data collection, management and future directions. Nat Commun 11, 2624, doi:10.1038/s41467-020-15948-9 (2020).

60 Smith, S. M. et al. An expanded set of genome-wide association studies of brain imaging phenotypes in UK Biobank. Nat Neurosci 24, 737–745, doi:10.1038/s41593-021-00826-4 (2021).

61 Pauli, W. M., Nili, A. N. & Tyszka, J. M. A high-resolution probabilistic in vivo atlas of human subcortical brain nuclei. Sci Data 5, 180063, doi:10.1038/sdata.2018.63 (2018).

62 Griffanti, L. et al. BIANCA (Brain Intensity AbNormality Classification Algorithm): A new tool for automated segmentation of white matter hyperintensities. NeuroImage 141, 191–205, doi:10.1016/j.neuroimage.2016.07.018 (2016).

63 Wang, C. et al. Methods for quantitative susceptibility and R2* mapping in whole post-mortem brains at 7T applied to amyotrophic lateral sclerosis. NeuroImage 222, 117216, doi:10.1016/j.neuroimage.2020.117216 (2020).

64 Iglesias, J. E. et al. A probabilistic atlas of the human thalamic nuclei combining ex vivo MRI and histology. NeuroImage 183, 314–326, doi:10.1016/j.neuroimage.2018.08.012 (2018).

65 Iglesias, J. E. et al. Bayesian longitudinal segmentation of hippocampal substructures in brain MRI using subject-specific atlases. NeuroImage 141, 542–555, doi:10.1016/j.neuroimage.2016.07.020 (2016).

66 Iglesias, J. E. et al. Bayesian segmentation of brainstem structures in MRI. NeuroImage 113, 184–195, doi:10.1016/j.neuroimage.2015.02.065 (2015).

67 Saygin, Z. M. et al. High-resolution magnetic resonance imaging reveals nuclei of the human amygdala: manual segmentation to automatic atlas. NeuroImage 155, 370–382, doi:10.1016/j.neuroimage.2017.04.046 (2017).

68 Neudorfer, C. et al. A high-resolution in vivo magnetic resonance imaging atlas of the human hypothalamic region. Sci Data 7, 305, doi:10.1038/s41597-020-00644-6 (2020).

69 Alfaro-Almagro, F. et al. Confound modelling in UK Biobank brain imaging. NeuroImage 224, 117002, doi:10.1016/j.neuroimage.2020.117002 (2021).

70 Vickers, A. J. The use of percentage change from baseline as an outcome in a controlled trial is statistically inefficient: a simulation study. BMC Med Res Methodol 1, 6, doi:10.1186/1471-2288-1-6 (2001).

71 Papst, I. et al. Age-dependence of healthcare interventions for COVID-19 in Ontario, Canada. BMC Public Health 21, 706, doi:10.1186/s12889-021-10611-4 (2021).

72 Levin, A. T. et al. Assessing the age specificity of infection fatality rates for COVID-19: systematic review, meta-analysis, and public policy implications. Eur J Epidemiol 35, 1123–1138, doi:10.1007/s10654-020-00698-1 (2020).

