## Supplementary_Information_Guide for "SARS-CoV-2 is associated with changes in brain structure in UK Biobank"

### Supplementary Discussion & References

**Various possible explanations for our longitudinal brain findings — Pages 1-2.**

### Supplementary Figures

**Supplementary Fig. 1 — Page 3. Histogram of the date of diagnosis.**

**Supplementary Fig. 2 — Page 4. Voxel-wise, cross-sectional baseline group differences between future infected participants and controls in grey matter volume and mean diffusivity (age-modulated).**

**Supplementary Fig. 3 — Page 5. Histogram of the time between diagnosis and the second scan.**

**Supplementary Fig. 4 — Page 6. Histograms of group comparison Z statistics of longitudinal change in cortical thickness.**

### Supplementary Tables

**Supplementary Tables 1-5 — Page 7.**

### Supplementary Plots

**Supplementary Longitudinal Plots — Pages 8-26.**

**Supplementary Baseline Plots — Pages 27-45.**

### Supplementary Analyses

**Supplementary Analysis 1 — Pages 46-48. Are nIDPs group-different at baseline, when clustering nIDPs using PCA?**

**Supplementary Analysis 2 — Pages 49-51. Cognitive tests sensitive to cognitive impairment in people at risk for dementia**

**Supplementary Analysis 3 — Pages 52-53. Does pneumonia show pre-post IDP changes?**

**Supplementary Analysis 4 — Pages 54-55. Does influenza show pre-post IDP changes?**

**Supplementary Analysis 5 — Page 56. The form of the case-control group-difference regressor**

**Supplementary Analysis 6 — Pages 57-58. Further model-fitting validity/robustness evaluations**

**Supplementary Analysis 7 — Pages 59-60. Do nIDPs explain the main (group-difference, longitudinal effect) results?**

**Supplementary Analyses References — Page 61.**
