## Supplementary for "SARS-CoV-2 is associated with changes in brain structure in UK Biobank"

### Supplementary Discussion

An alternative explanation for the brain differences revealed in this longitudinal study might be that the coronavirus infection leads to neuroinflammation, which can initiate chronic neuronal dysfunction[1,2]. In particular, activation of the peripheral innate immune system can induce the production of inflammatory cytokines in the brain, leading not only to severe impairment in memory, cognition and emotional behaviour, but also to microglial abnormalities in the hippocampus, which is particularly vulnerable to neuroinflammation[3-5]. In line with these findings, in the case of an averred neurotropic virus such as influenza, which spreads to central nervous system via infection of the olfactory neurons in the olfactory epithelium[6], various long-term inflammatory-induced functional and structural alterations of the hippocampus, accompanied by impairment in spatial memory, have been recently observed[7]. Even in the possible absence of neurotropism, mild SARS-CoV-2 infection appears to lead to impaired hippocampal neurogenesis, decreased oligodendrocytes and myelin loss[8]. It is also possible that the choroid plexus, which appears to relay peripheral inflammation due to SARS-CoV-2 into the brain[2], plays a role in the specific targeting of olfactory regions, as they might regulate the dynamics of the subventricular zone responsible for the neurogenesis occurring in the olfactory bulbs[9]. (Maladaptive) immune regulation through the microglia might also modulate these neurogenic niches[10]. Finally, other factors related to being infected by SARS-CoV-2, such as added anxiety, stress or isolation, might play a role in our findings. However, we did not find any non-imaging phenotype (measured pre-infection) that, when controlled for, greatly reduced the association of infection with the longitudinal IDP effects. It is also worth noting that most of the cases involved in this study were either asymptomatic or mild — and indeed, most of our significant olfactory-related results stand when excluding the infected participants with more moderate or severe COVID-19 (hospitalised cases) — and most of the controls would have had also been exposed to higher levels of anxiety, stress and isolation during the pandemic in the UK. In addition, brain regions typically involved in these mental health factors, while overlapping with our limbic results, do not affect these regions consistently: higher and lower volumes have both been observed in these limbic structures for anxiety and stress, including higher (as opposed to lower in our study) volume of the parahippocampal gyrus in stress[11-14], while social isolation impacts a different network of brain regions (the so-called “default mode” network), which does not overlap with the pattern associated with SARS-CoV-2 infection in our results[15]. While the number of influenza cases arising between two scans is probably too low at present in UK Biobank to draw any firm conclusion, further longitudinal investigations we carried out out-of-sample demonstrated that the pattern of abnormalities associated with pneumonia (not related to COVID-19) do not overlap with that of SARS-CoV-2 (**Supplementary Analysis 3**). Whether our findings, which seem to delineate a clear limbic network of the primary and secondary olfactory and possibly gustatory cortex, reflect a corollary effect of the infection, an indirect brain-related pathophysiological process of SARS-CoV-2 through either anterograde degeneration or neuroinflammatory process, or a direct effect of the spread of the virus itself, remains to be elucidated.

### Supplementary Discussion References


1 Deleidi, M. & Isacson, O. Viral and inflammatory triggers of neurodegenerative diseases. *Sci Transl Med* **4**, 121ps123, doi:10.1126/scitranslmed.3003492 (2012).

2 Yang, A. C. *et al.* Dysregulation of brain and choroid plexus cell types in severe COVID-19. *Nature*, doi:10.1038/s41586-021-03710-0 (2021).

3 Camara, M. L. *et al.* Effects of centrally administered etanercept on behavior, microglia, and astrocytes in mice following a peripheral immune challenge. *Neuropsychopharmacology* **40**, 502-512, doi:10.1038/npp.2014.199 (2015).

4 Klein, R. S., Garber, C. & Howard, N. Infectious immunity in the central nervous system and brain function. *Nat Immunol* **18**, 132-141, doi:10.1038/ni.3656 (2017).

5 Elmore, M. R. *et al.* Respiratory viral infection in neonatal piglets causes marked microglia activation in the hippocampus and deficits in spatial learning. *J Neurosci* **34**, 2120-2129, doi:10.1523/JNEUROSCI.2180-13.2014 (2014).

6 Ludlow, M. *et al.* Neurotropic virus infections as the cause of immediate and delayed neuropathology. *Acta Neuropathol* **131**, 159-184, doi:10.1007/s00401-015-1511-3 (2016).

7 Hosseini, S. *et al.* Long-Term Neuroinflammation Induced by Influenza A Virus Infection and the Impact on Hippocampal Neuron Morphology and Function. *J Neurosci* **38**, 3060-3080, doi:10.1523/JNEUROSCI.1740-17.2018 (2018).

8 Fernández-Castañeda, A. *et al*. Mild respiratory SARS-CoV-2 infection can cause multi-lineage cellular dysregulation and myelin loss in the brain. bioRxiv, doi.org/10.1101/2022.01.07. 475453 (2022).

9 Falcao, A. M. *et al.* The path from the choroid plexus to the subventricular zone: go with the flow! *Front Cell Neurosci* **6**, 34, doi:10.3389/fncel.2012.00034 (2012).

10 Chintamen, S., Imessadouene, F. & Kernie, S. G. Immune Regulation of Adult Neurogenic Niches in Health and Disease. *Front Cell Neurosci* **14**, 571071, doi:10.3389/fncel.2020.571071 (2020).

11 Wang, X., Cheng, B., Luo, Q., Qiu, L. & Wang, S. Gray Matter Structural Alterations in Social Anxiety Disorder: A Voxel-Based Meta-Analysis. *Front Psychiatry* **9**, 449, doi:10.3389/fpsyt.2018.00449 (2018).

12 Wang, X. *et al.* Distinct grey matter volume alterations in adult patients with panic disorder and social anxiety disorder: A systematic review and voxel-based morphometry meta-analysis. *J Affect Disord* **281**, 805-823, doi:10.1016/j.jad.2020.11.057 (2021).

13 Bromis, K., Calem, M., Reinders, A., Williams, S. C. R. & Kempton, M. J. Meta-Analysis of 89 Structural MRI Studies in Posttraumatic Stress Disorder and Comparison With Major Depressive Disorder. *Am J Psychiatry* **175**, 989-998, doi:10.1176/appi.ajp.2018.17111199 (2018).

14 Kuhn, S. & Gallinat, J. Gray matter correlates of posttraumatic stress disorder: a quantitative meta-analysis. *Biological psychiatry* **73**, 70-74, doi:10.1016/j.biopsych.2012.06.029 (2013).

15 Spreng, R. N. *et al.* The default network of the human brain is associated with perceived social isolation. *Nat Commun* **11**, 6393, doi:10.1038/s41467-020-20039-w (2020).

### Supplementary Figures

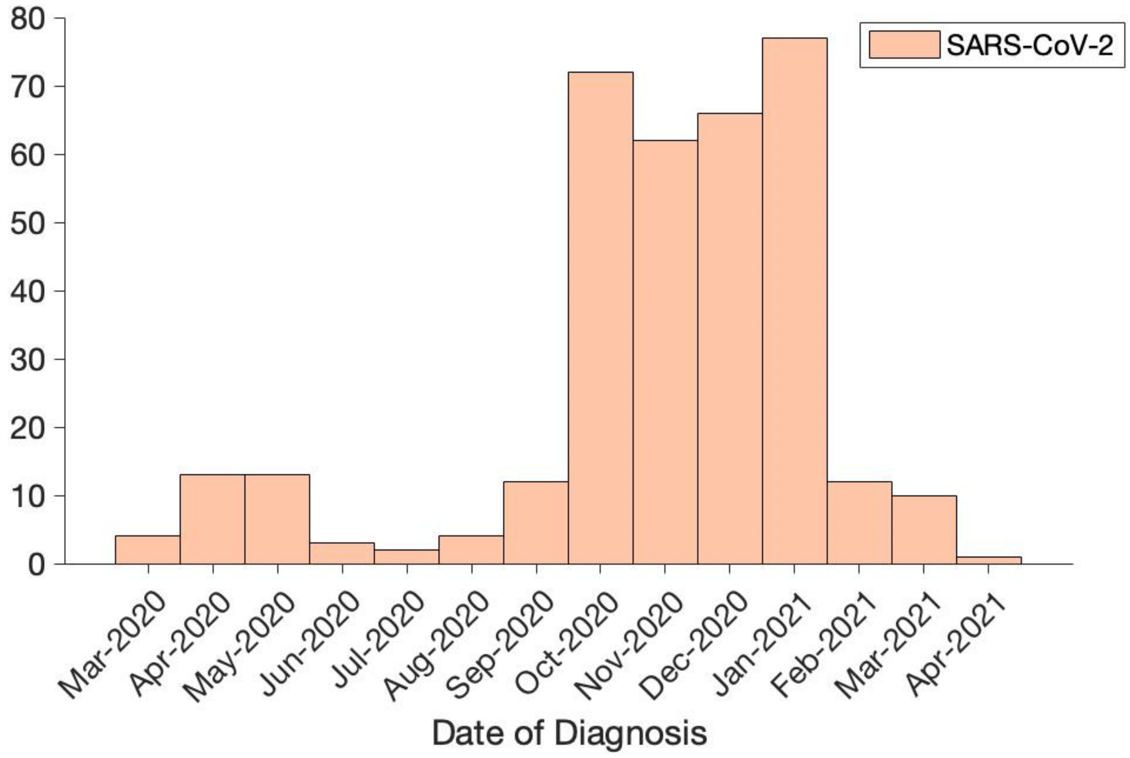


**Supplementary Fig. 1. Histogram of the date of diagnosis.** Diagnosis date was available for 351 cases, covering periods when the original strain, as well as the Alpha, Beta and Gamma variants were in circulation.

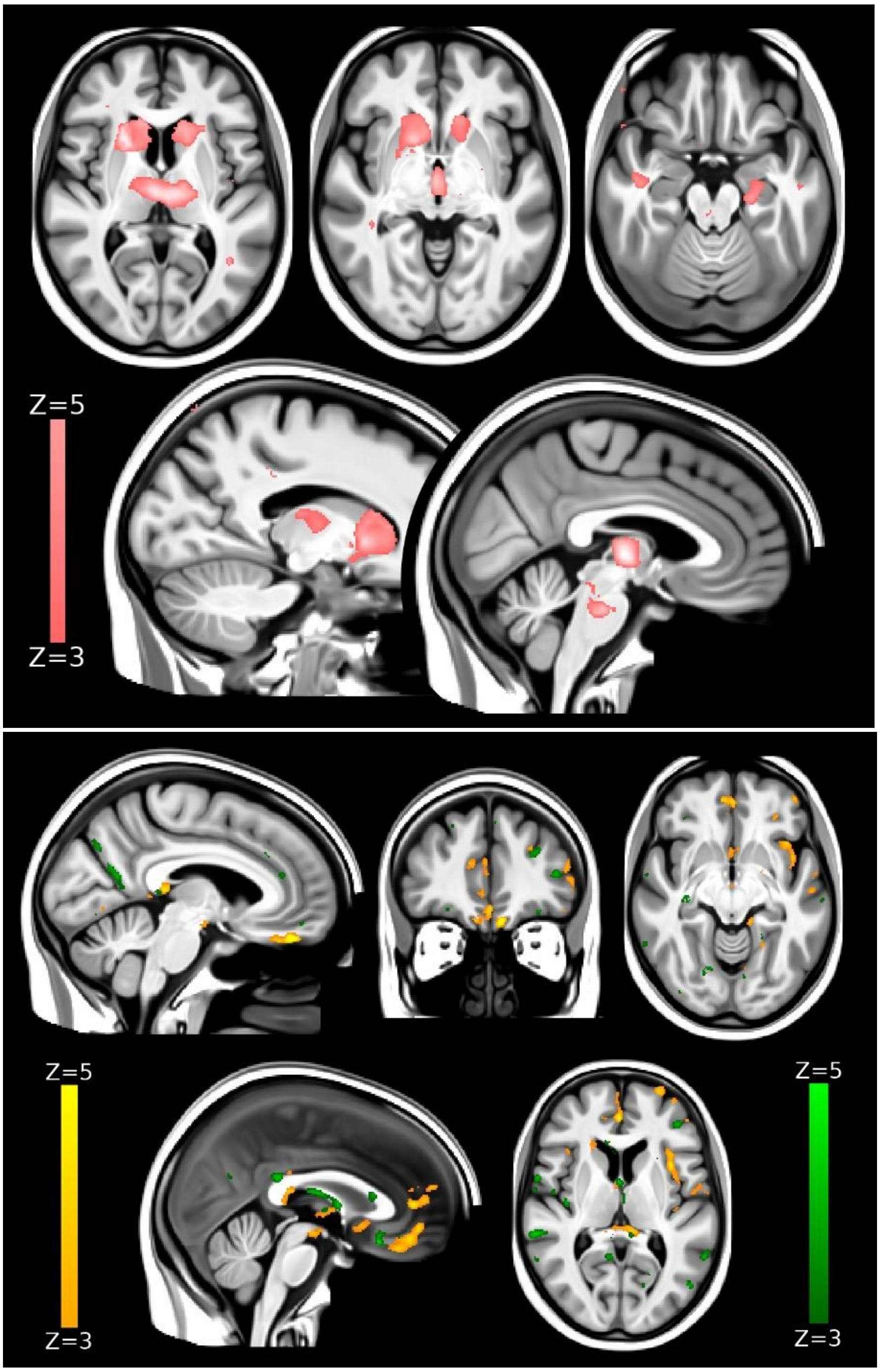


**Supplementary Fig. 2. Voxel-wise, cross-sectional baseline group differences between future infected participants and controls in grey matter volume and mean diffusivity (age-modulated). Top row.** The thresholded map (|Z|>3) shows that the localised lower grey matter volume at baseline in the future 401 SARS-CoV-2 positive participants compared with the 384 controls are bilaterally in the subcortical structures, specifically the caudate nucleus, putamen, ventral striatum, thalamus and hippocampus, and in the brainstem. None of these regions spatially overlap with the main longitudinal results. **Bottom row.** The thresholded map (|Z|>3) of mean diffusivity shows very few, scattered clusters of baseline differences (green), not overlapping with those longitudinal differences between the two groups (orange). We show the voxel-wise cross-sectional effects for illustrative purposes, avoiding any thresholding based on significance (as this would be statistically circular).

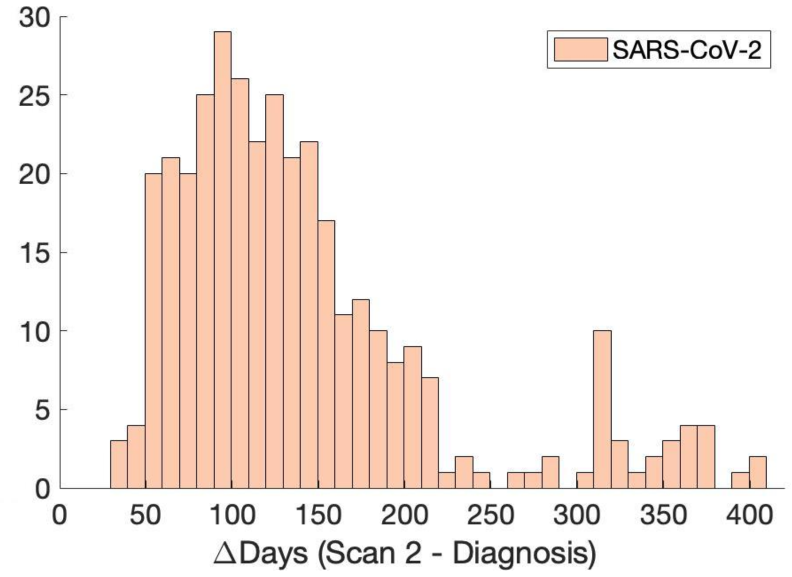


**Supplementary Fig. 3. Histogram of the time between diagnosis and the second scan.** Diagnosis date was available for 351 cases, more than 80% of whom had less than 6 months between infection and their second scan.

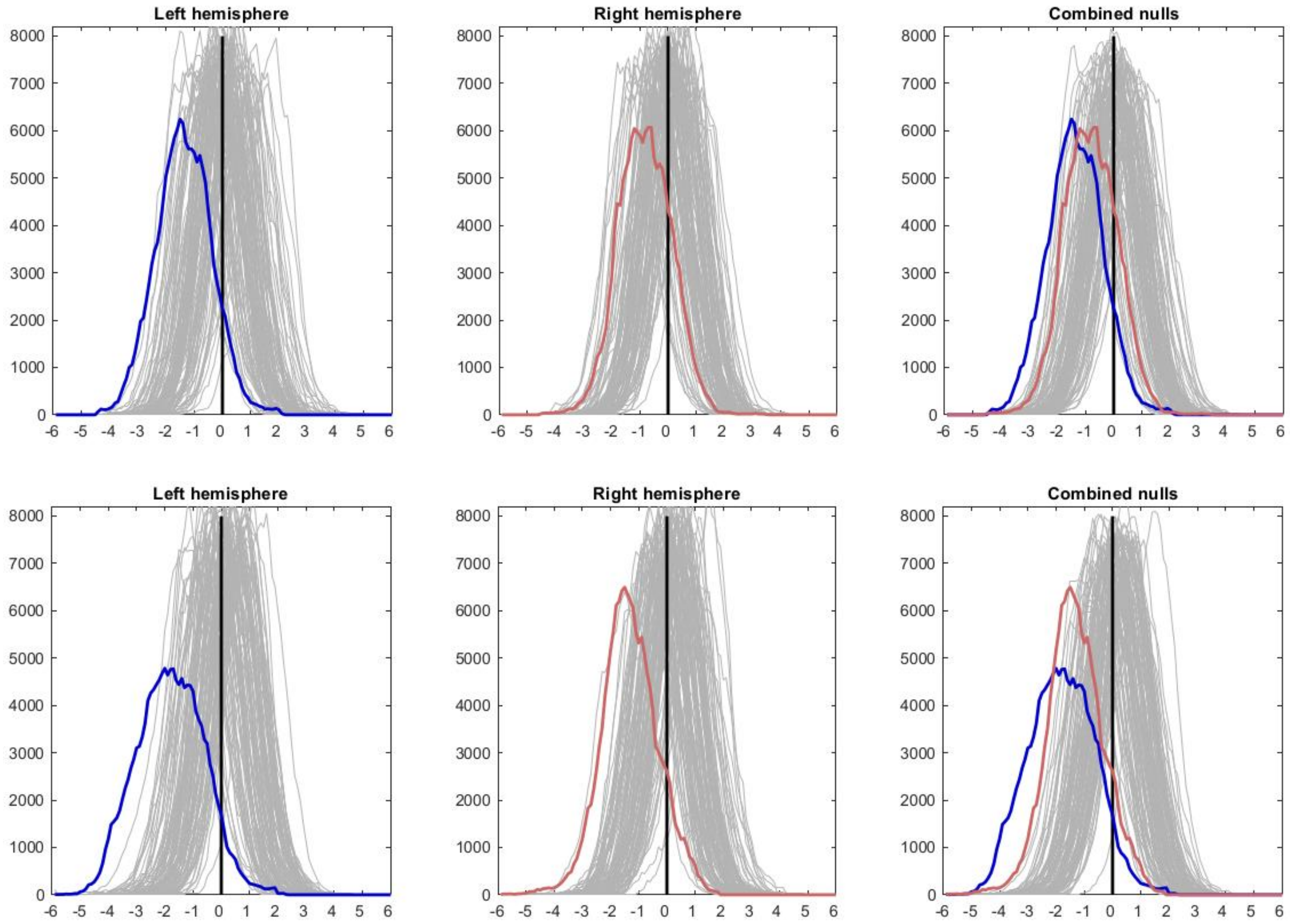


**Supplementary Fig. 4. Histograms of group comparison Z statistics of longitudinal change in cortical thickness. Top row.** Model 1: All SARS-CoV-2 participants vs controls. Left, histogram of Z-statistics (blue) across all cortical vertices of the left hemisphere (with grey lines showing 100 null histograms created through random permutations of the group variable). Middle: right hemisphere Z-statistics (orange) and matched nulls. Right: the same left and right hemisphere Z-statistics histograms overlaid (with a pooled null histogram in grey). **Bottom row.** Model 4: All hospitalised vs non-hospitalised SARS-CoV-2 participants. Same representation of Z-statistics histograms as in the top row.

### Supplementary Tables

**Supplementary Table 1. Full list of reproducible imaging-derived phenotypes (IDPs) used in the hypothesis-driven and exploratory approaches, and corresponding statistics for the longitudinal analyses (Models 1-4).**

Please see separate xls spreadsheet for the full table.

**Supplementary Table 2: Full list of reproducible imaging-derived phenotypes (IDPs) used in the hypothesis-driven and exploratory approaches, and corresponding statistics for the cross-sectional, baseline analysis comparing SARS-CoV-2 and control groups (binary and age-modulated).**

Please see separate xls spreadsheet for the full table.

**Supplementary Table 3: Full list of reproducible imaging-derived phenotypes (IDPs) used in the hypothesis-driven and exploratory approaches, and corresponding statistics for the cross-sectional, second timepoint analysis comparing SARS-CoV-2 and control groups (binary and age-modulated).**

Please see separate xls spreadsheet for the full table.

**Supplementary Table 4. Full list of non-imaging phenotypes (nIDPs) used for the cross-sectional, baseline comparison between SARS-CoV-2 and control groups, and corresponding statistics (binary).**

Please see separate xls spreadsheet for the full table.

**Supplementary Table 5: Full list of reproducible imaging-derived phenotypes (IDPs) used in the hypothesis-driven and exploratory approaches, and corresponding statistics for the longitudinal analyses using a binary regressor for group comparisons.**

Please see separate xls spreadsheet for the full table.

### Supplementary Longitudinal Plots

Longitudinal percentage changes with age plots, as well as scatter and box plots for the top 10 results found using the hypothesis-driven approach, and for the top 10 results found using the exploratory approach.

#### Longitudinal hypothesis-driven results

Temporal piriform cortex functional network – OD

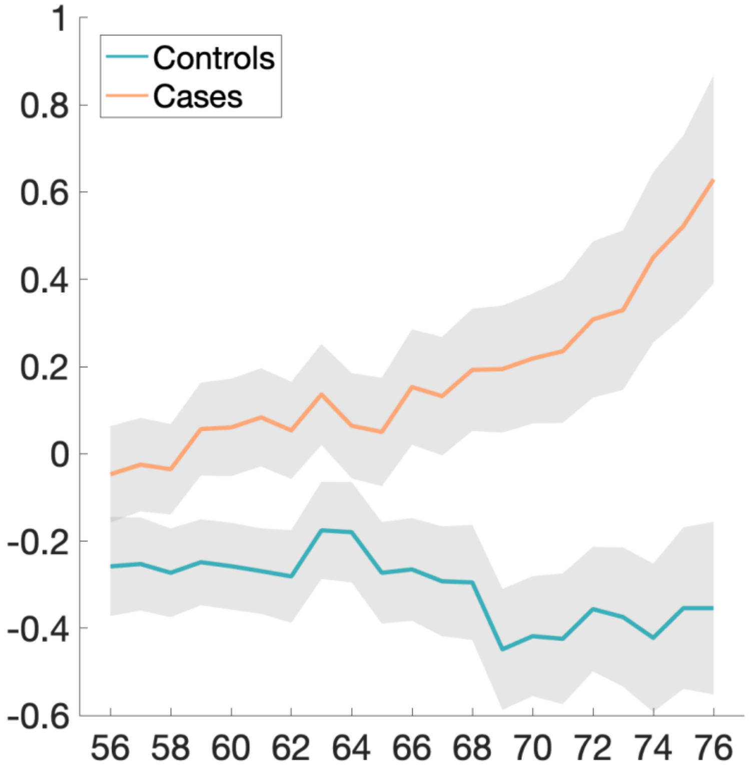


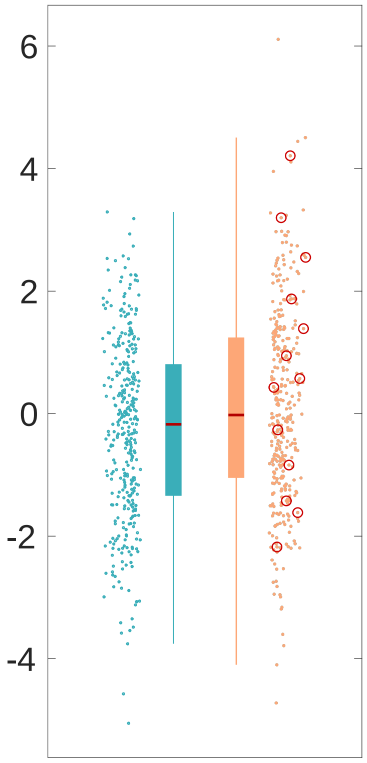

Olfactory tubercle functional network – ISOVF

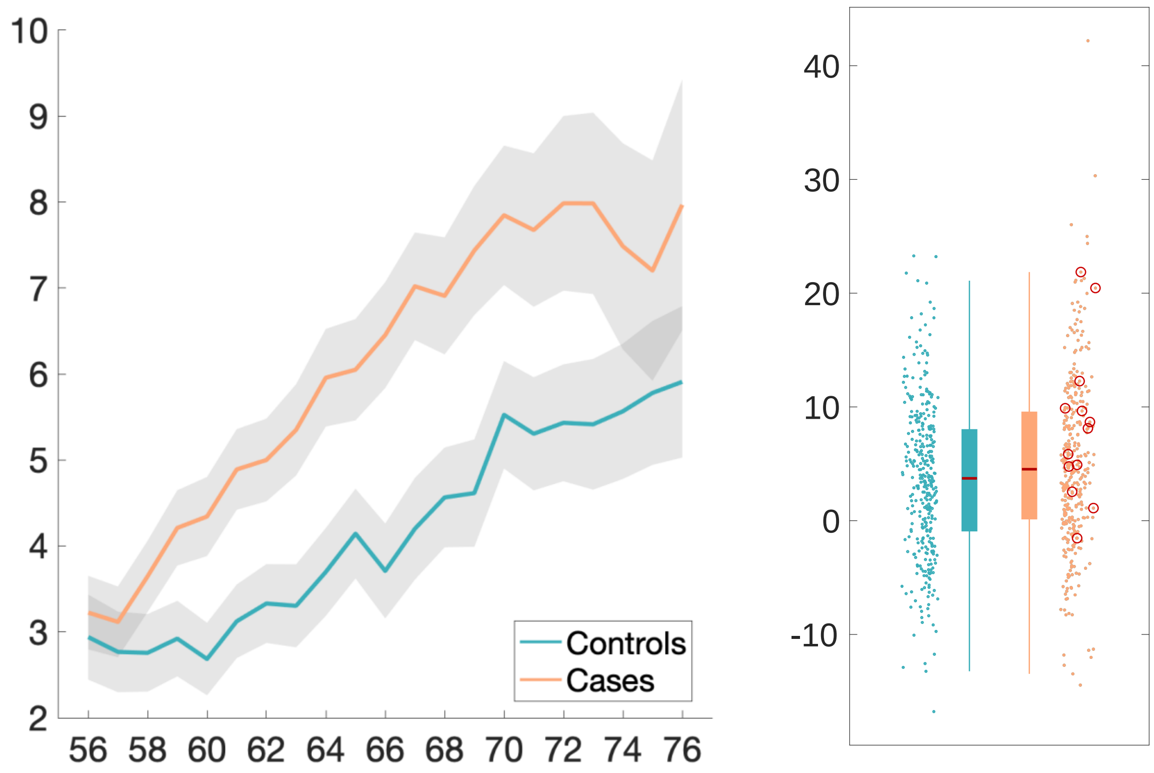

Frontal piriform cortex functional network – MD (mean)

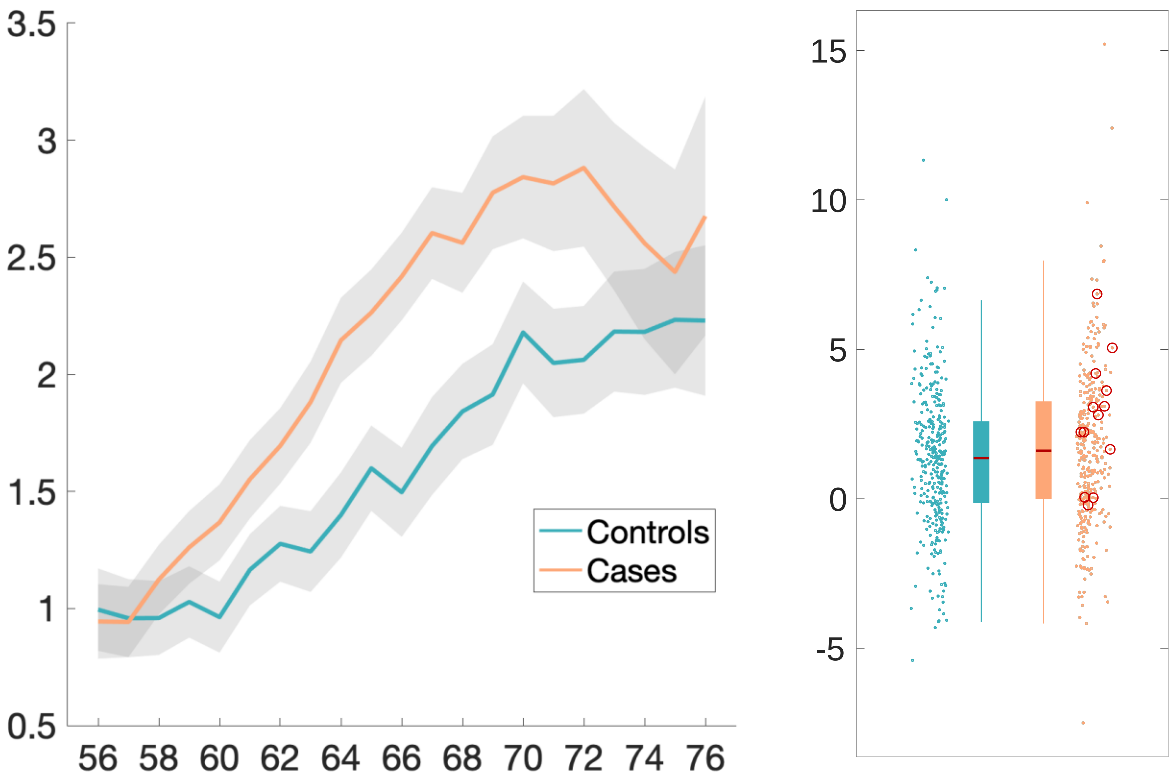

Temporal piriform cortex functional network – MD

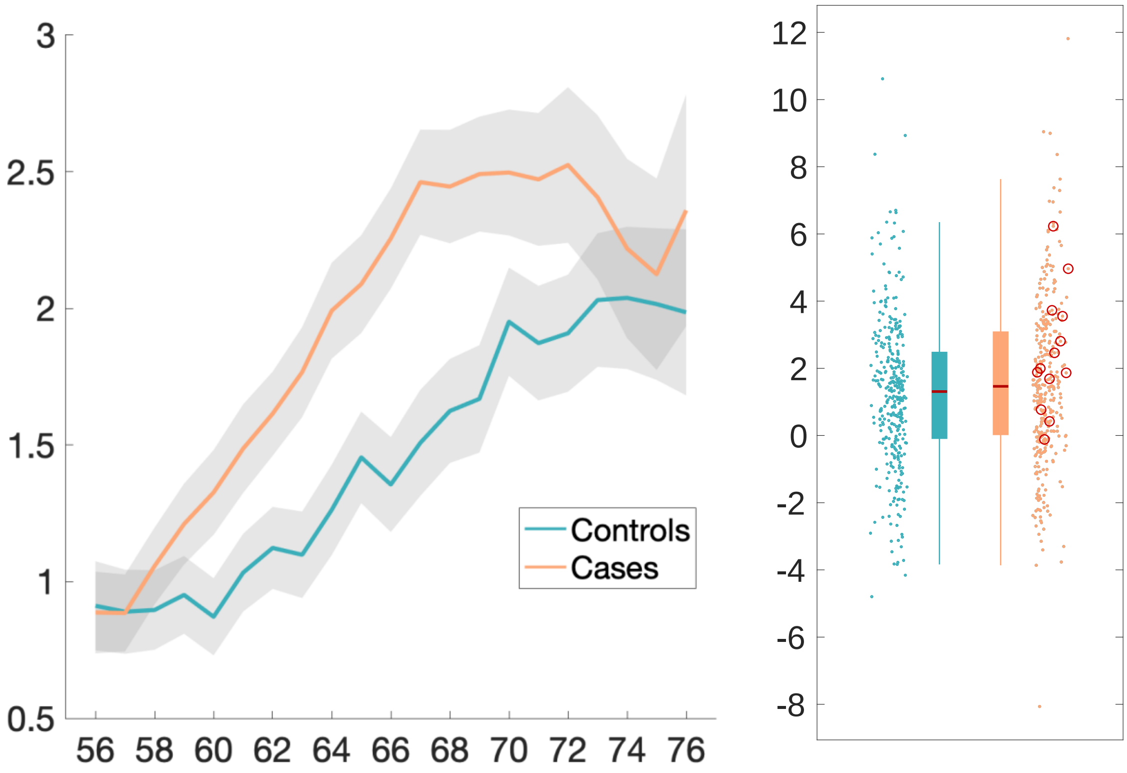

Olfactory tubercle functional network – MD

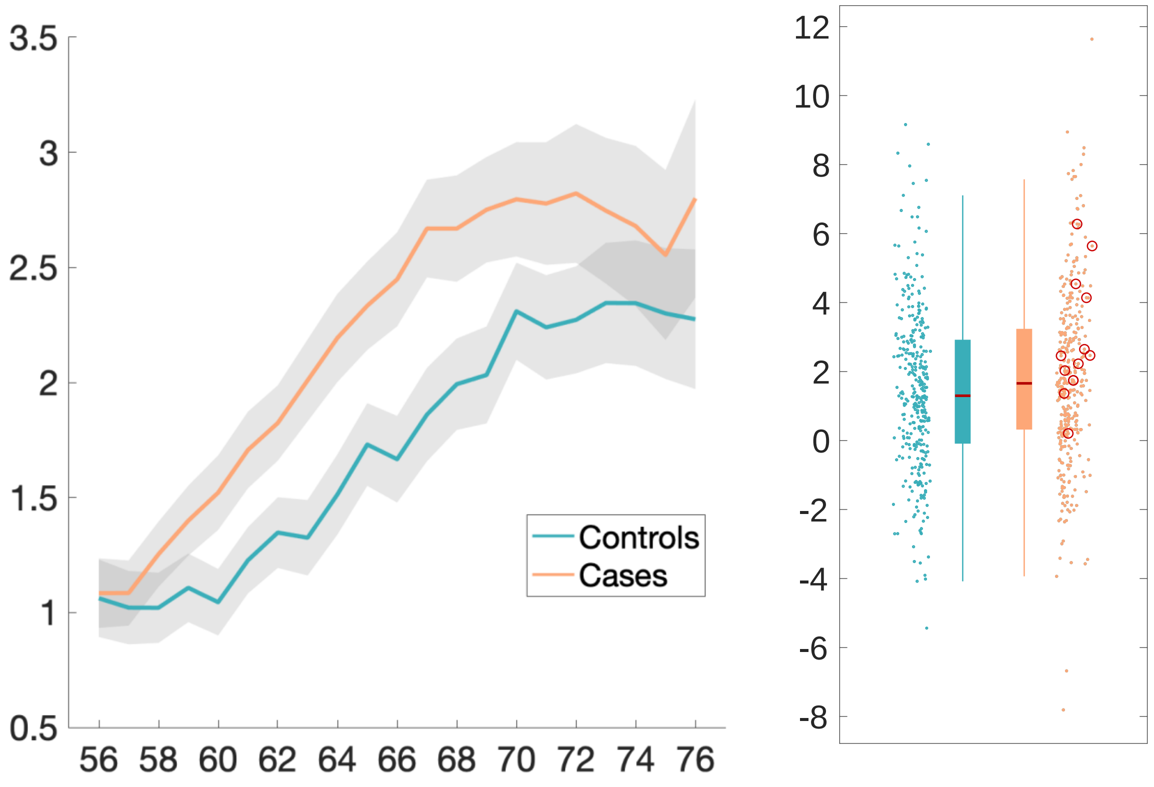

Lateral orbitofrontal cortex L – thickness (DKT atlas)

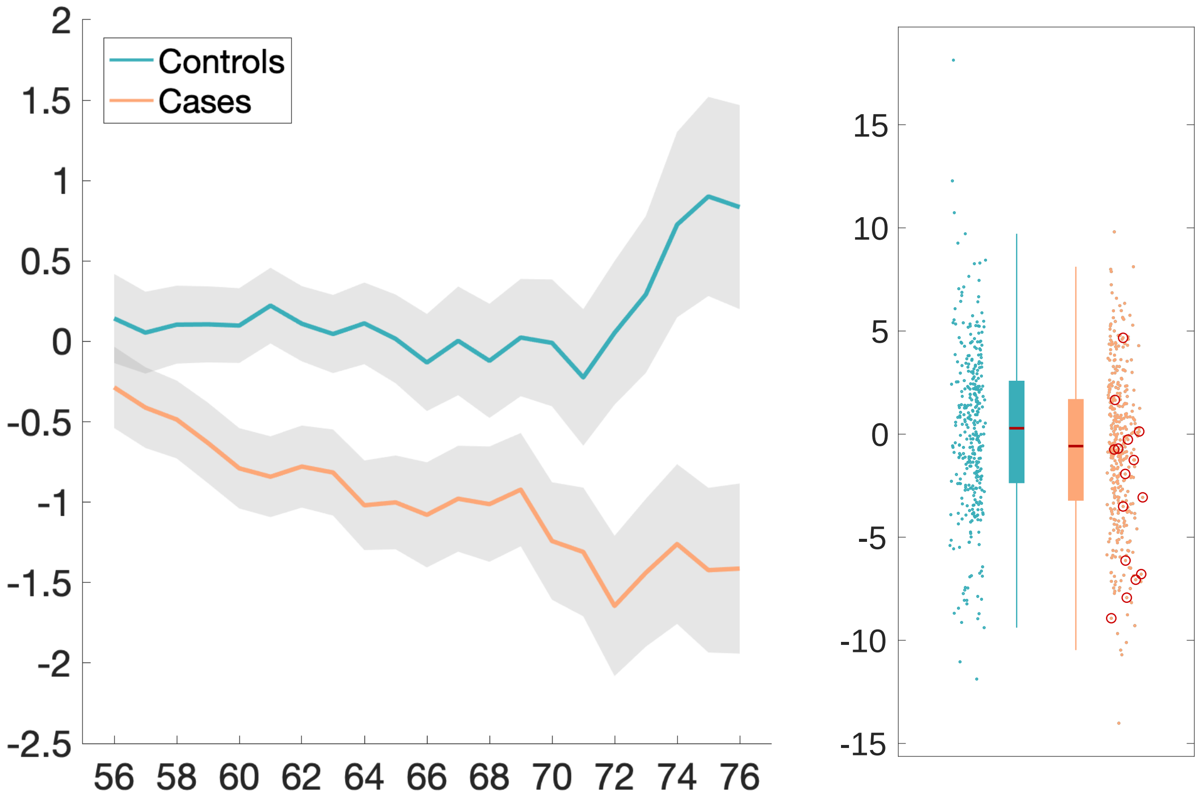

Temporal piriform cortex functional network – ISOVF

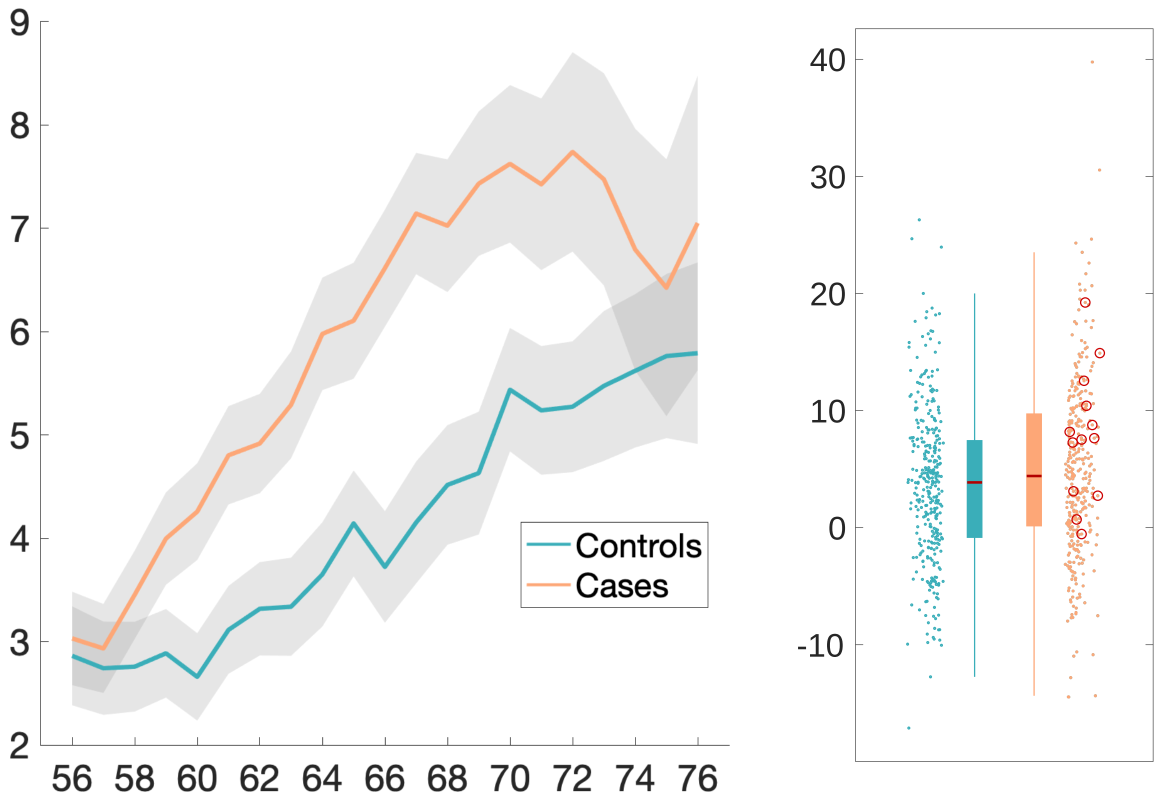

Anterior olfactory nucleus functional network – MD

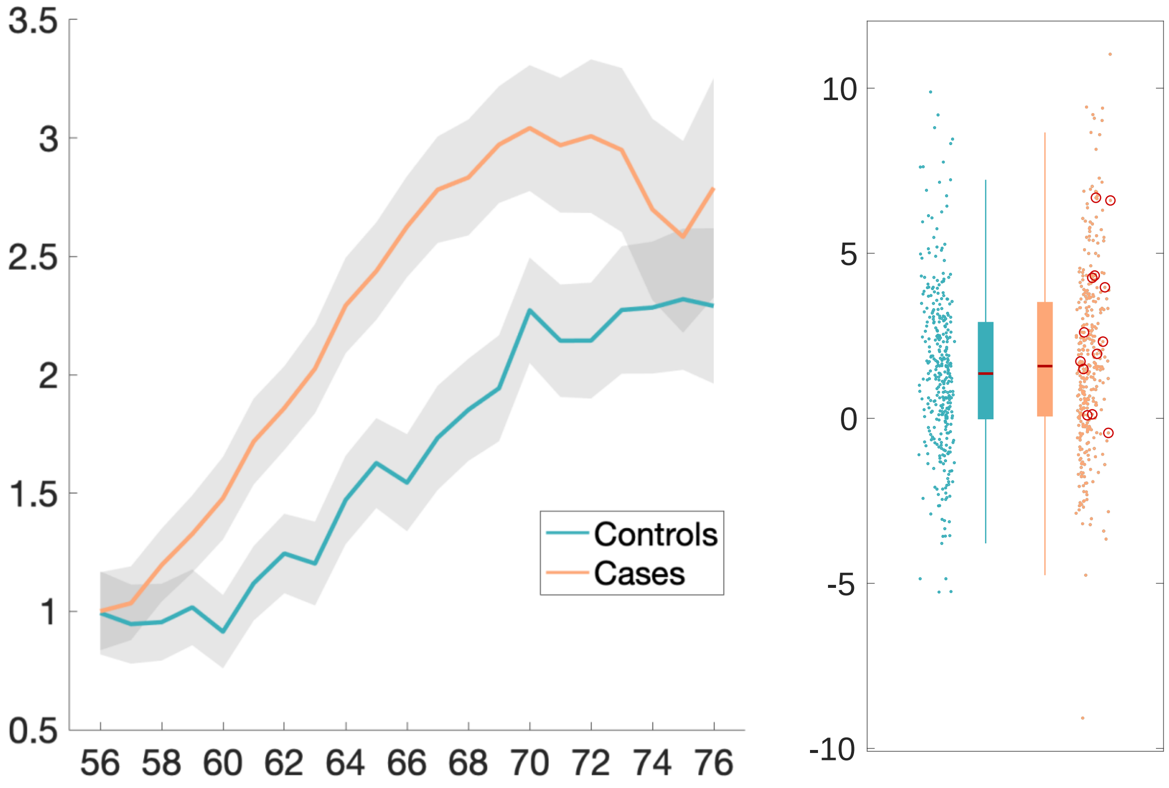

Parahippocampal gyrus L – intensity contrast (Desikan atlas)

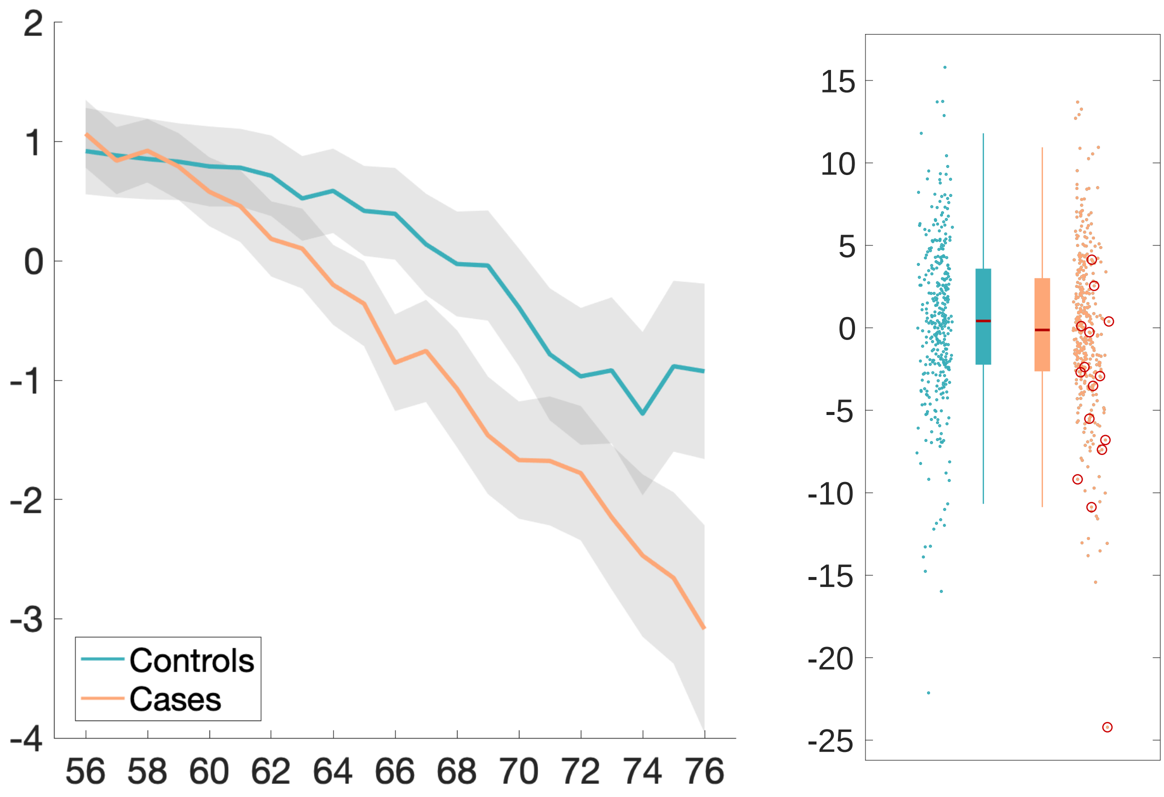

Anterior olfactory nucleus functional network – ISOVF

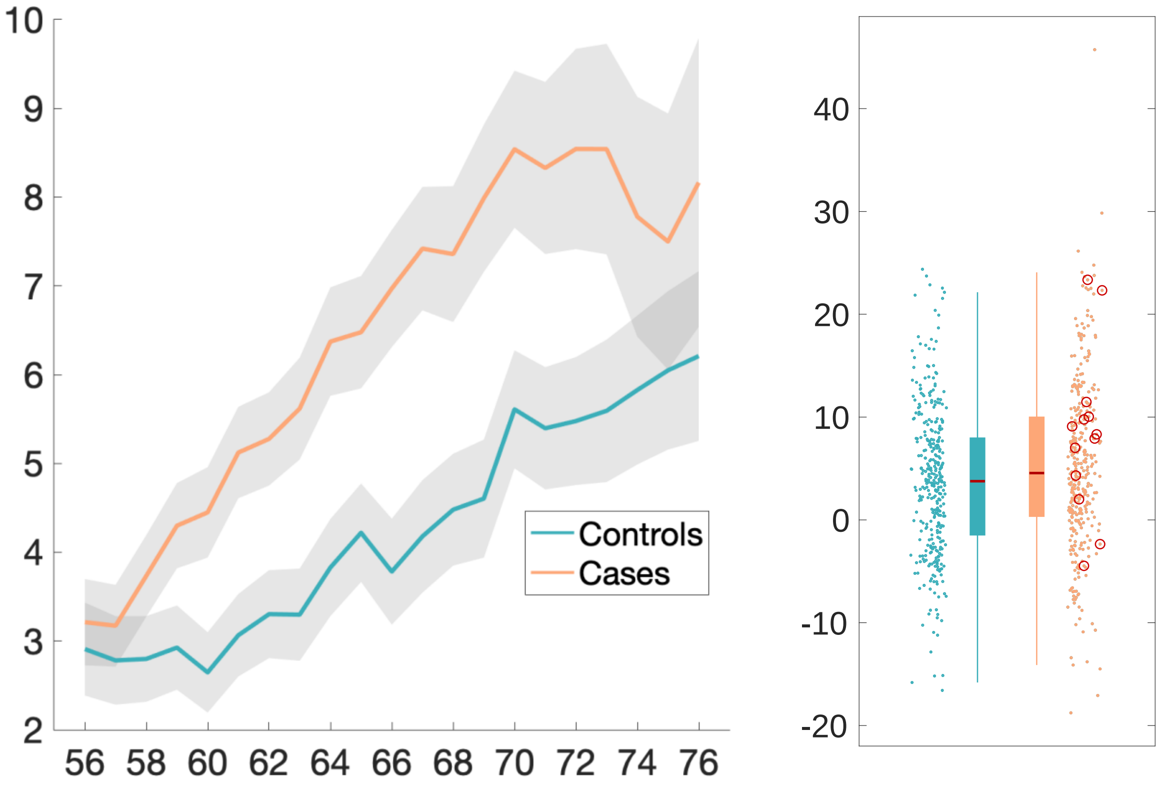

**Longitudinal exploratory results**

Ratio brain volume/estimated total intracranial volume

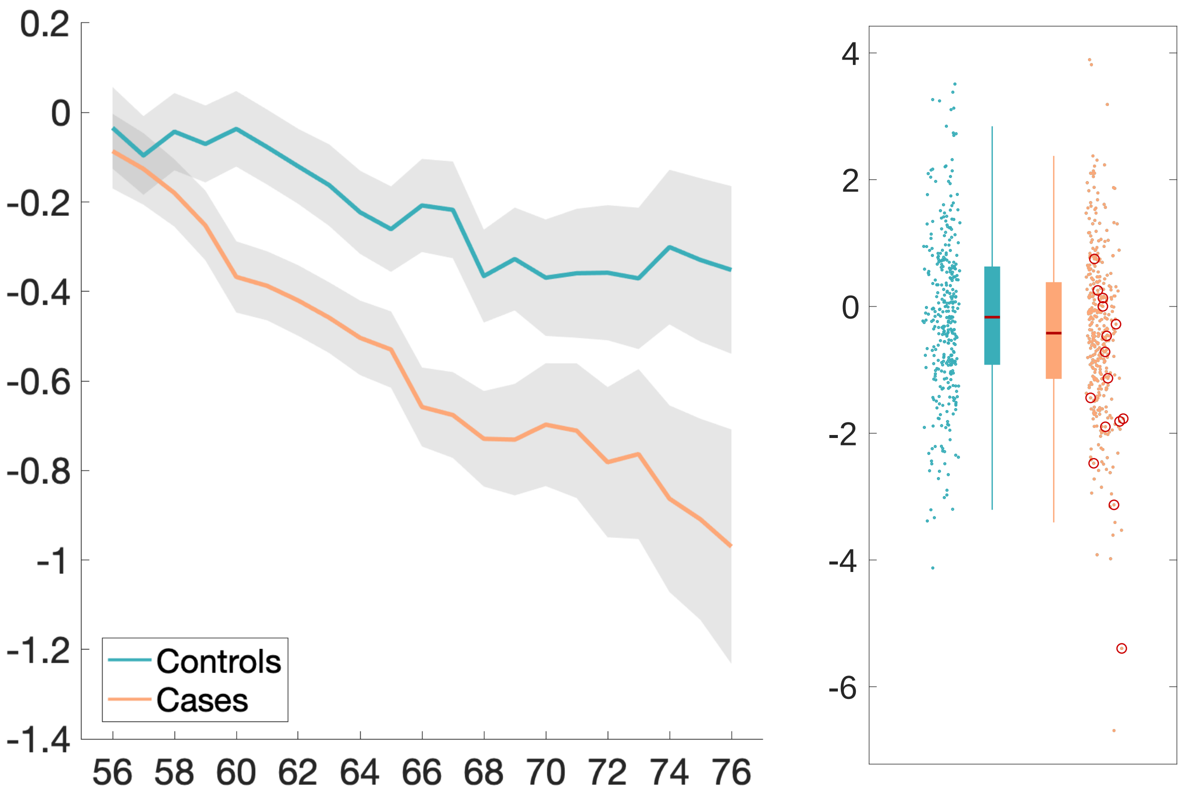

Normalised CSF – volume

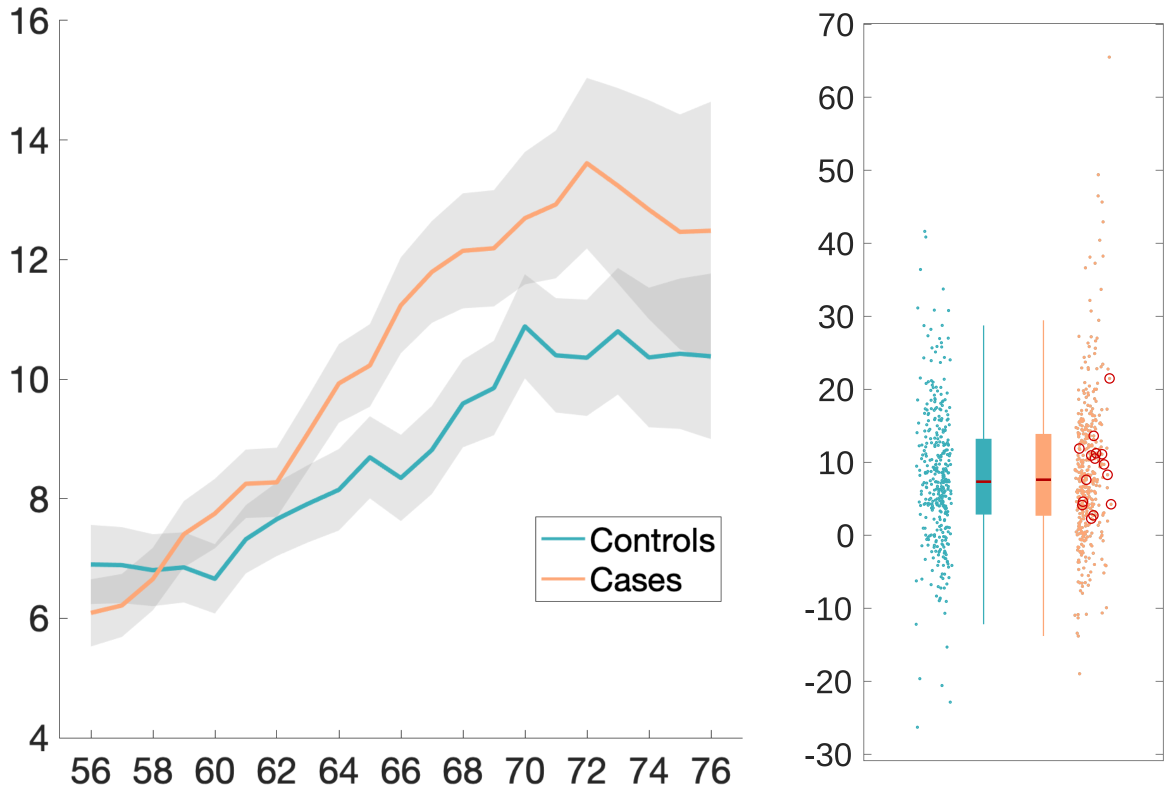

Lateral ventricle R – volume

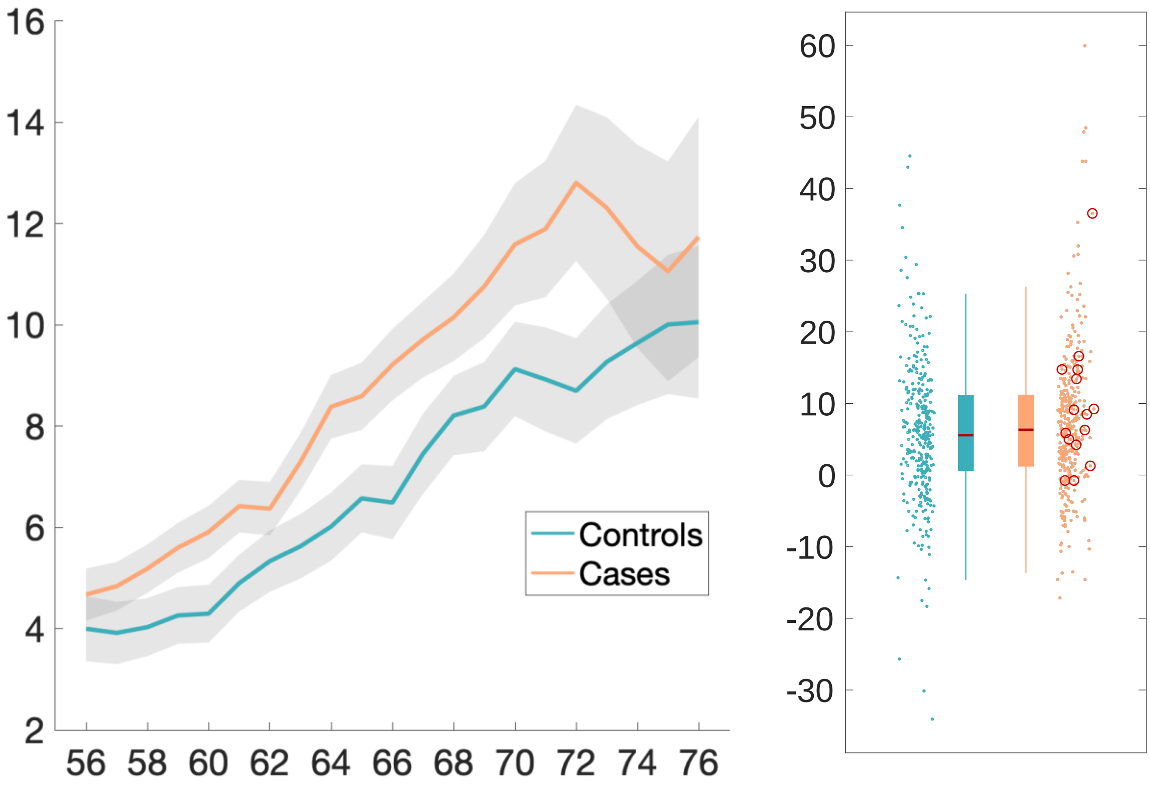

Superior fronto-occipital fasciculus R – ICVF

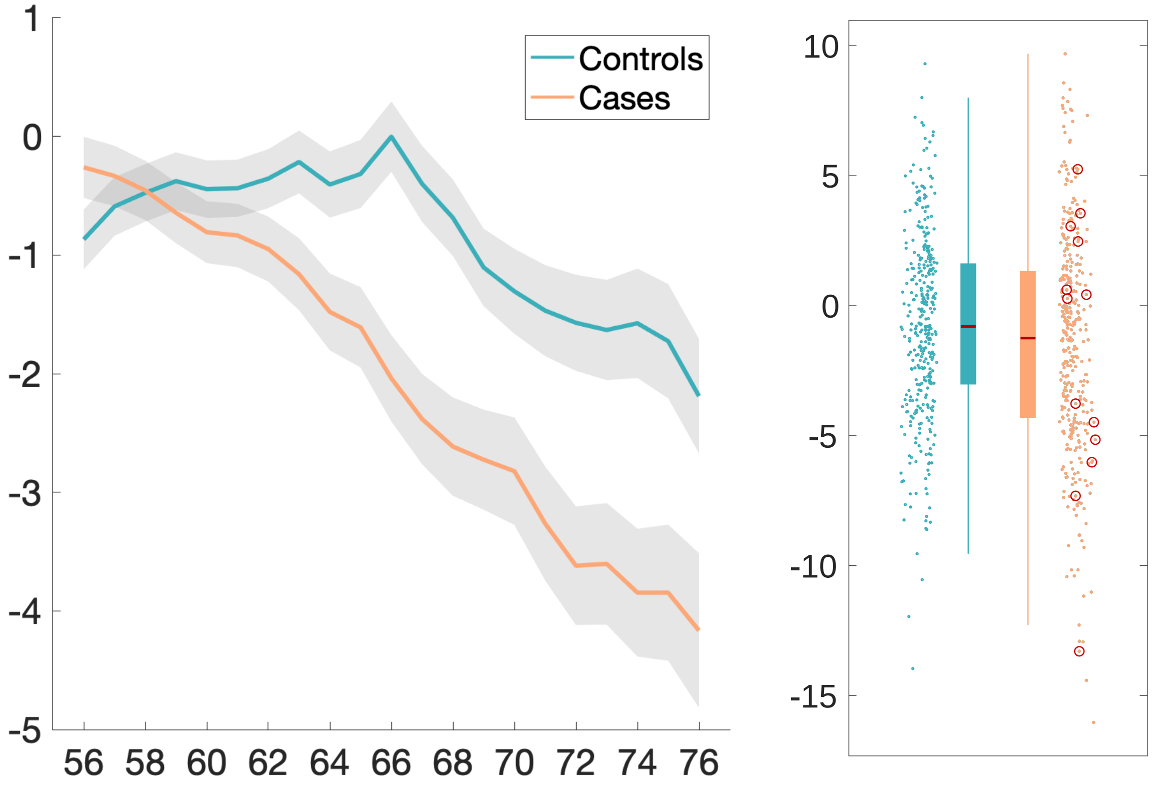

Brain volume without ventricles (surface model estimate)

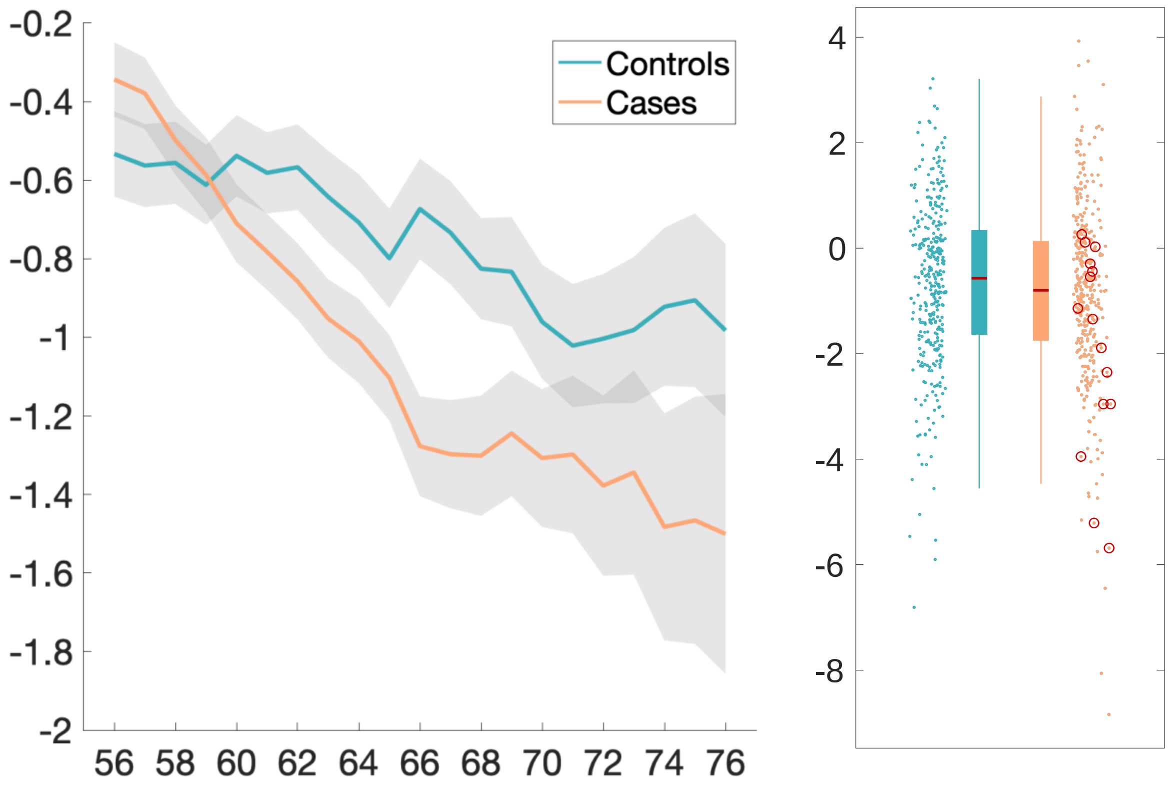

Rostral anterior cingulate cortex L – thickness (Desikan atlas)

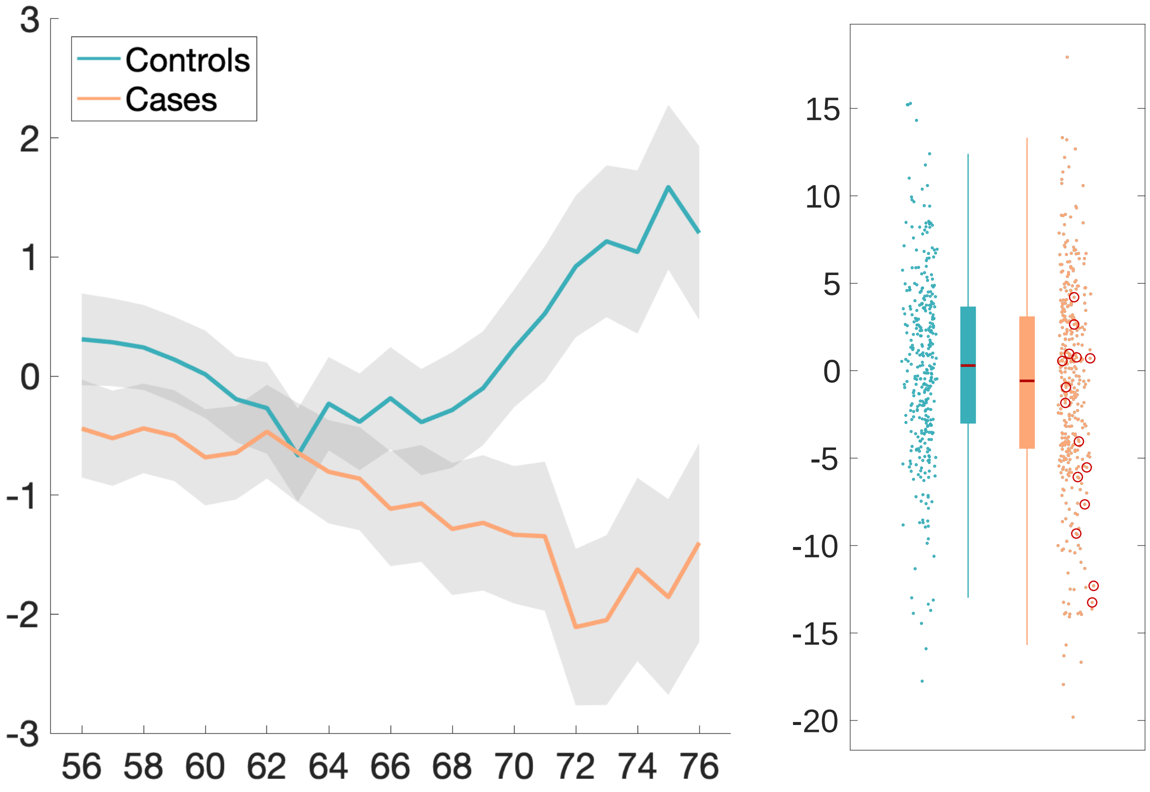

Brain volume without ventricles

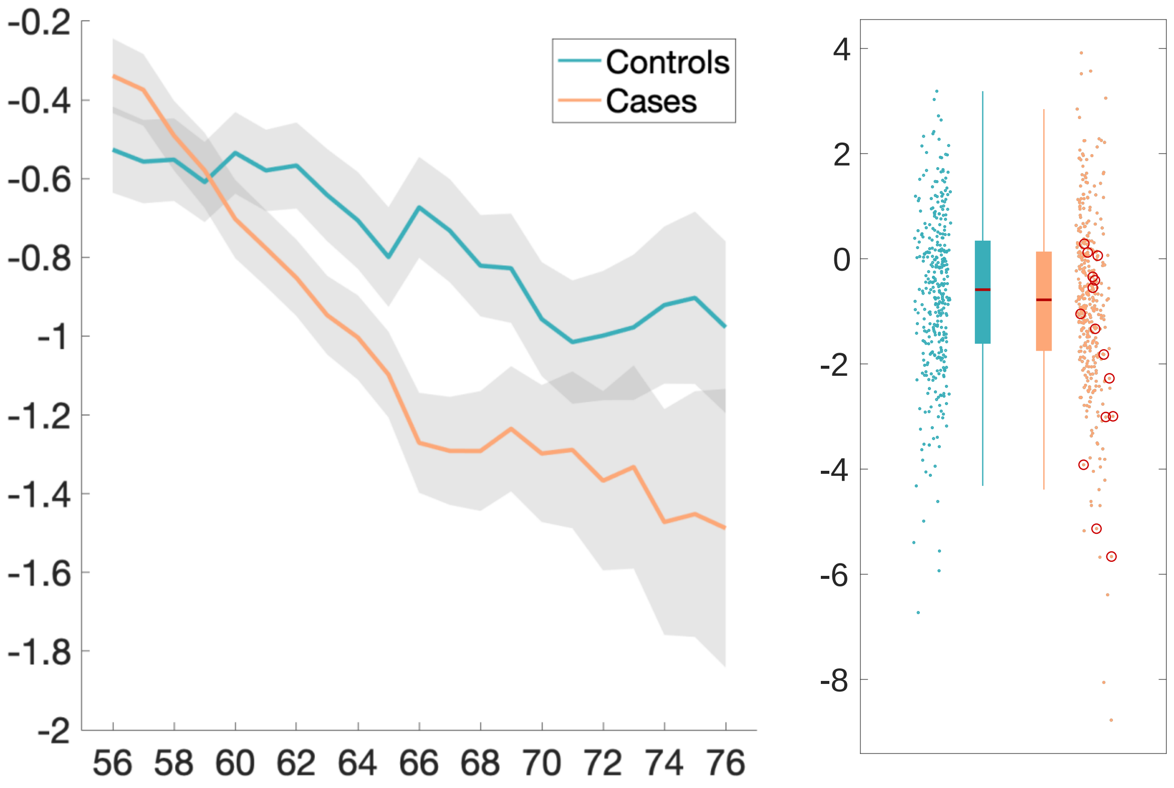

Supratentorial volume without ventricles

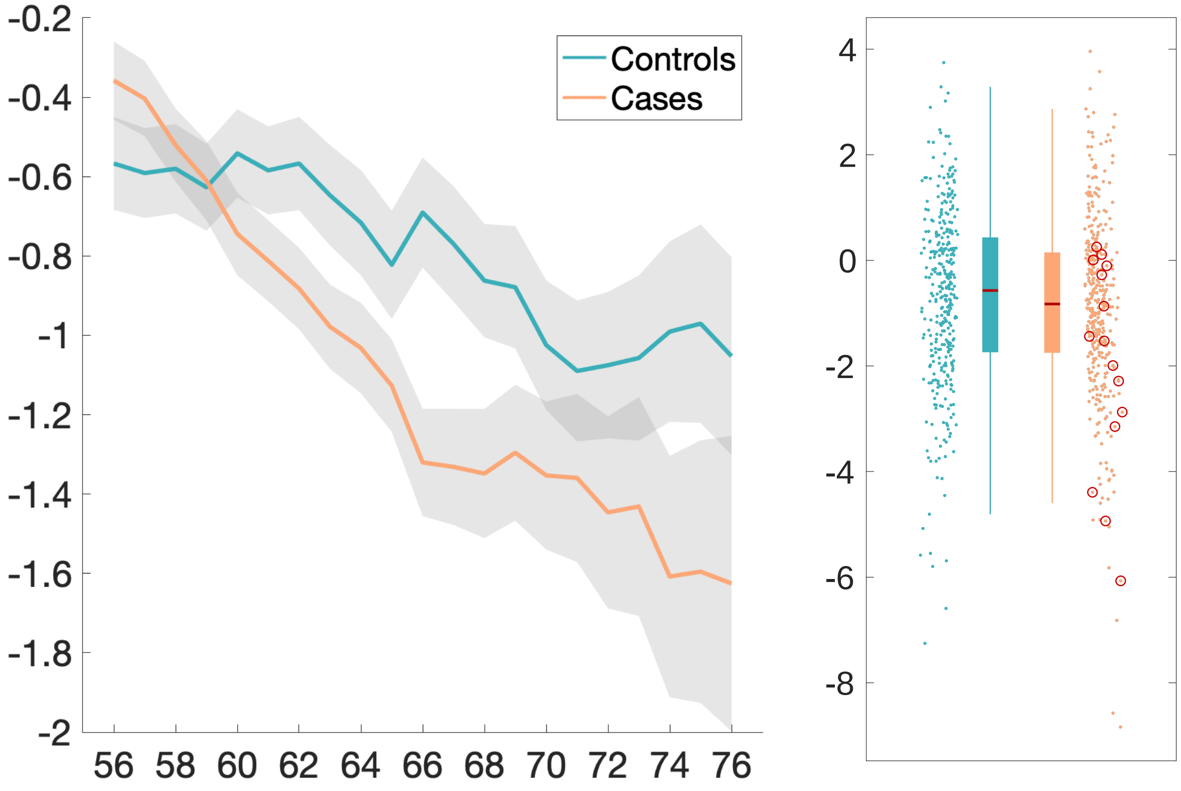

Cerebellum crus II – volume

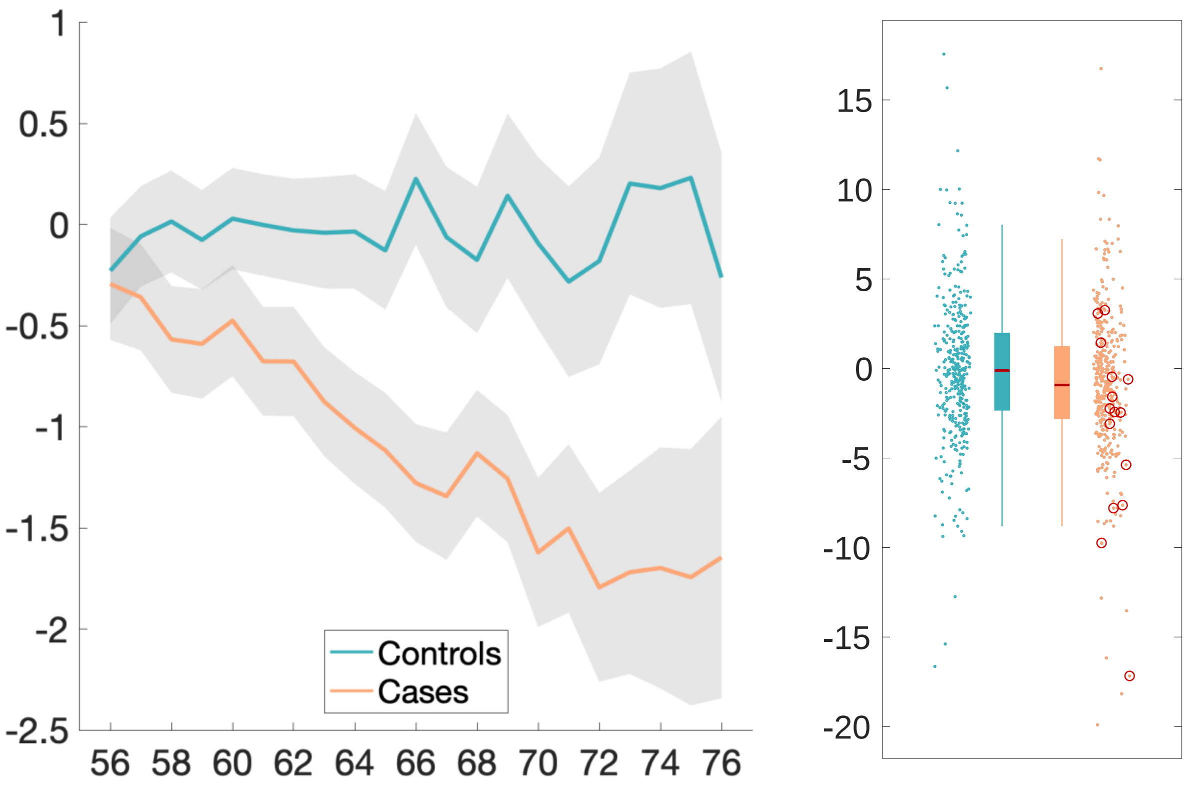

**Supplementary Baseline Plots.** Percentage difference at baseline with age plots, as well as scatter and box plots for the top 10 IDPs showing longitudinal effects using the hypothesis-driven approach, and for the top 10 longitudinal IDPs found using the exploratory approach.

### Baseline hypothesis-driven results

Temporal piriform cortex functional network – OD

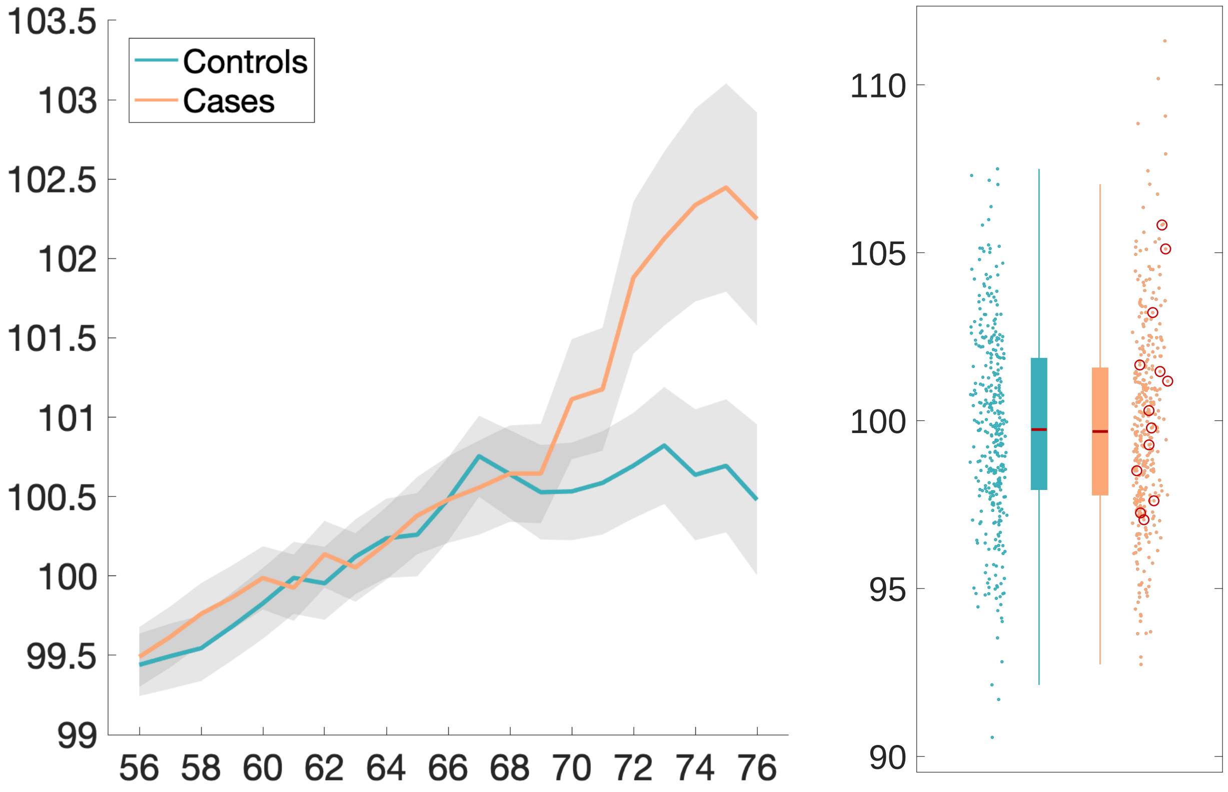

Olfactory tubercle functional network – ISOVF

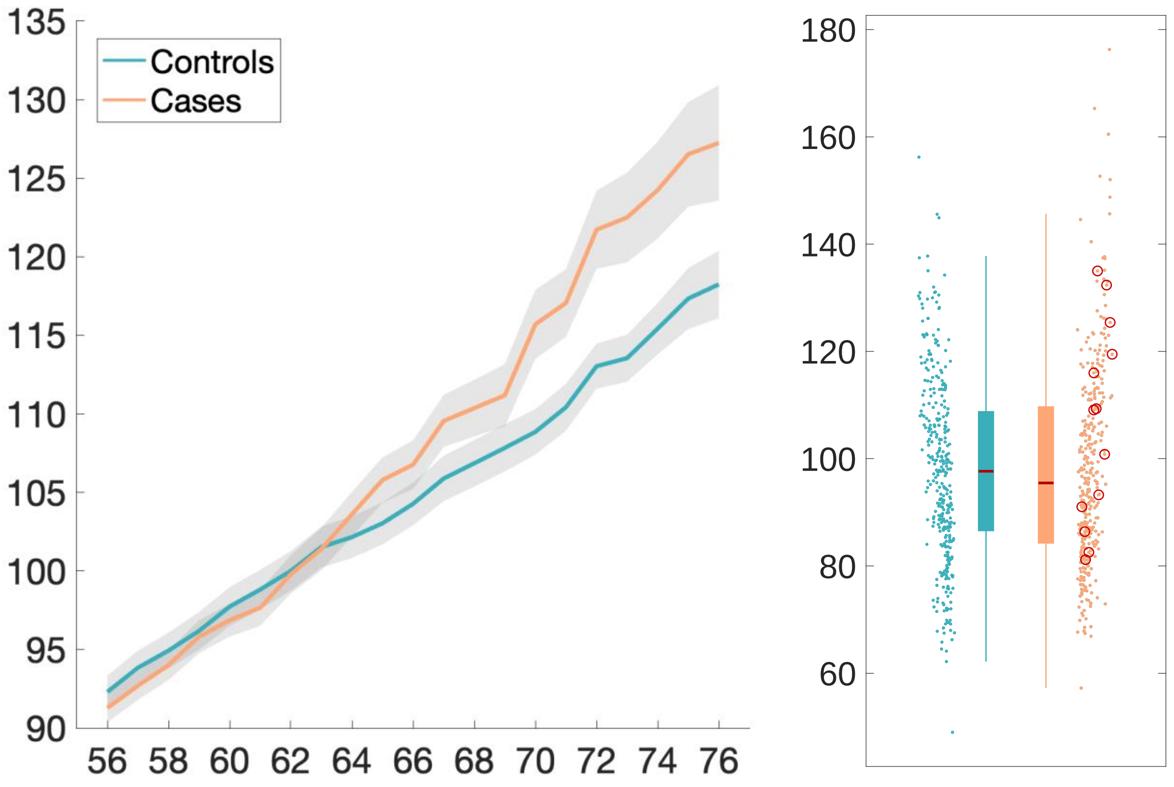

Frontal piriform cortex functional network – MD (mean)

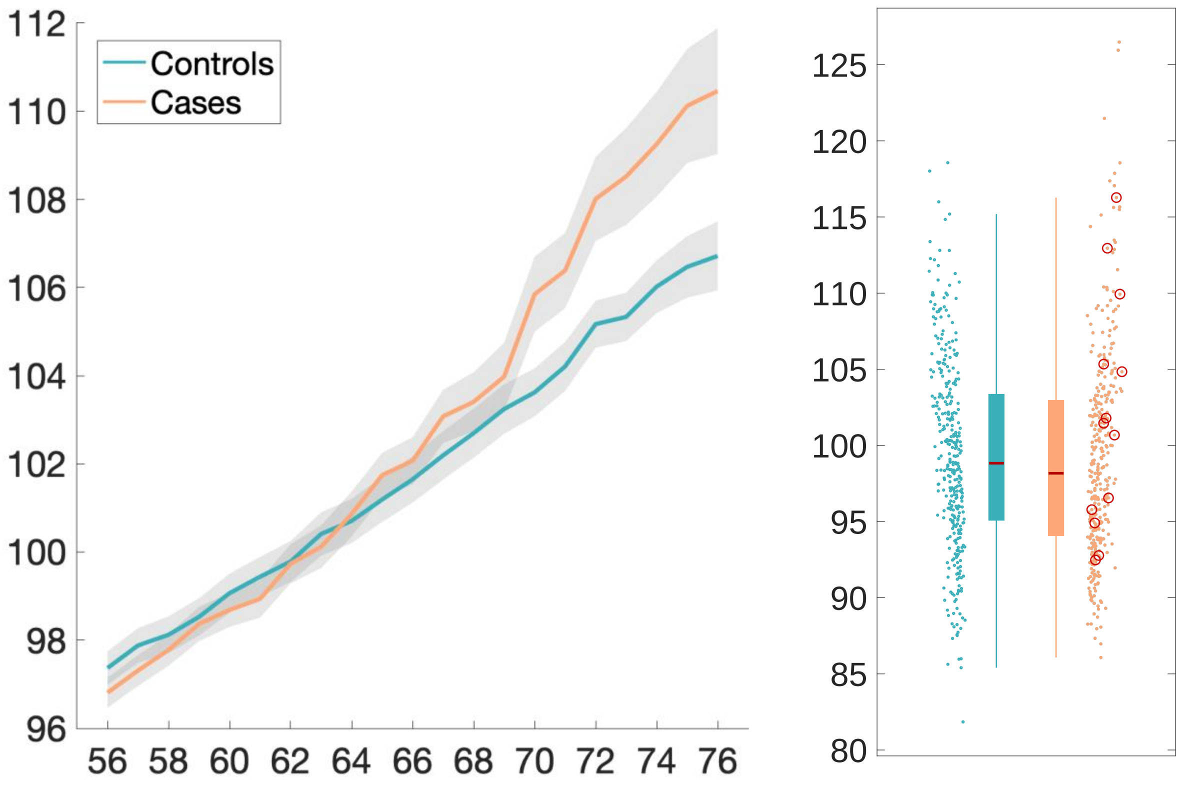

Temporal piriform cortex functional network – MD

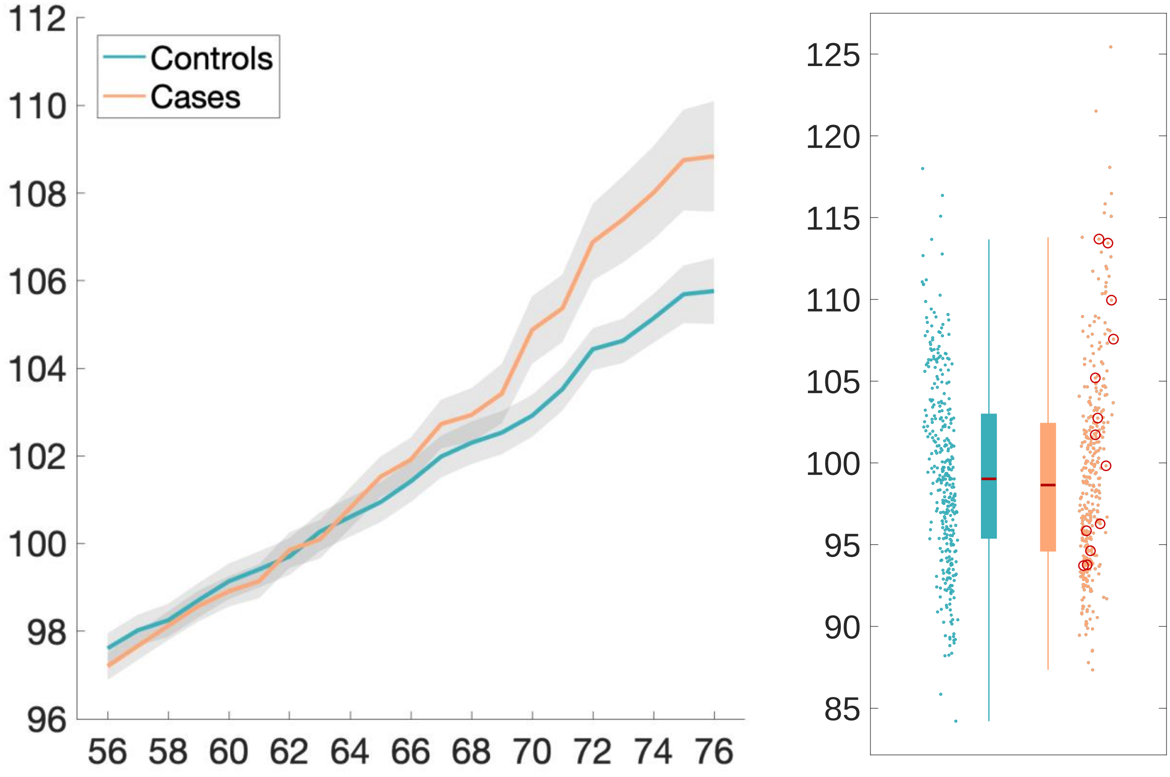

Olfactory tubercle functional network – MD

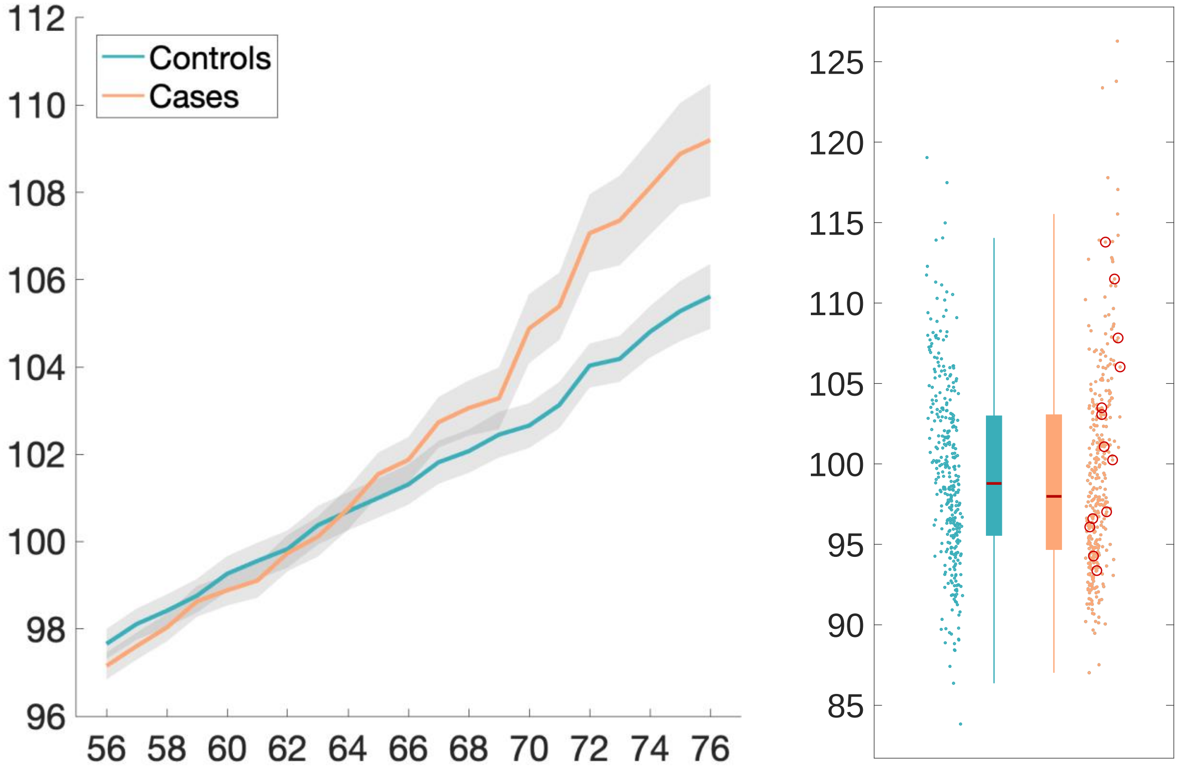

Lateral orbitofrontal cortex L – thickness (DKT atlas)

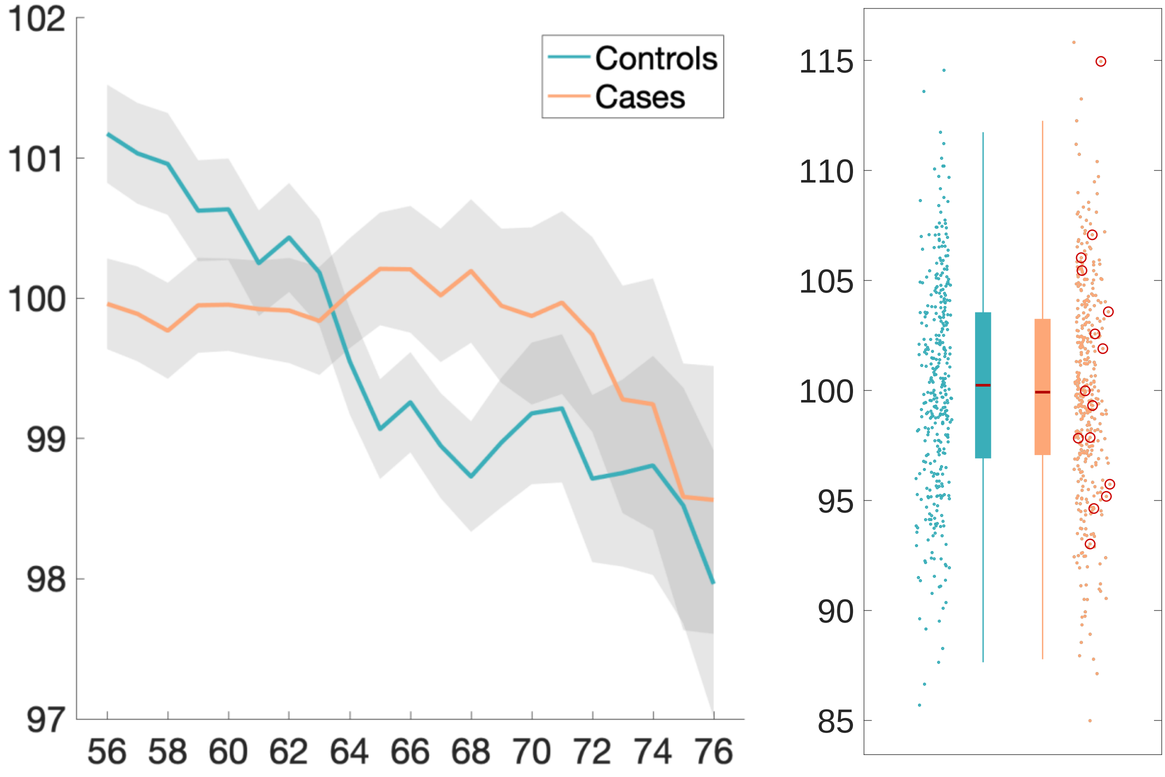

Temporal piriform cortex functional network – ISOVF

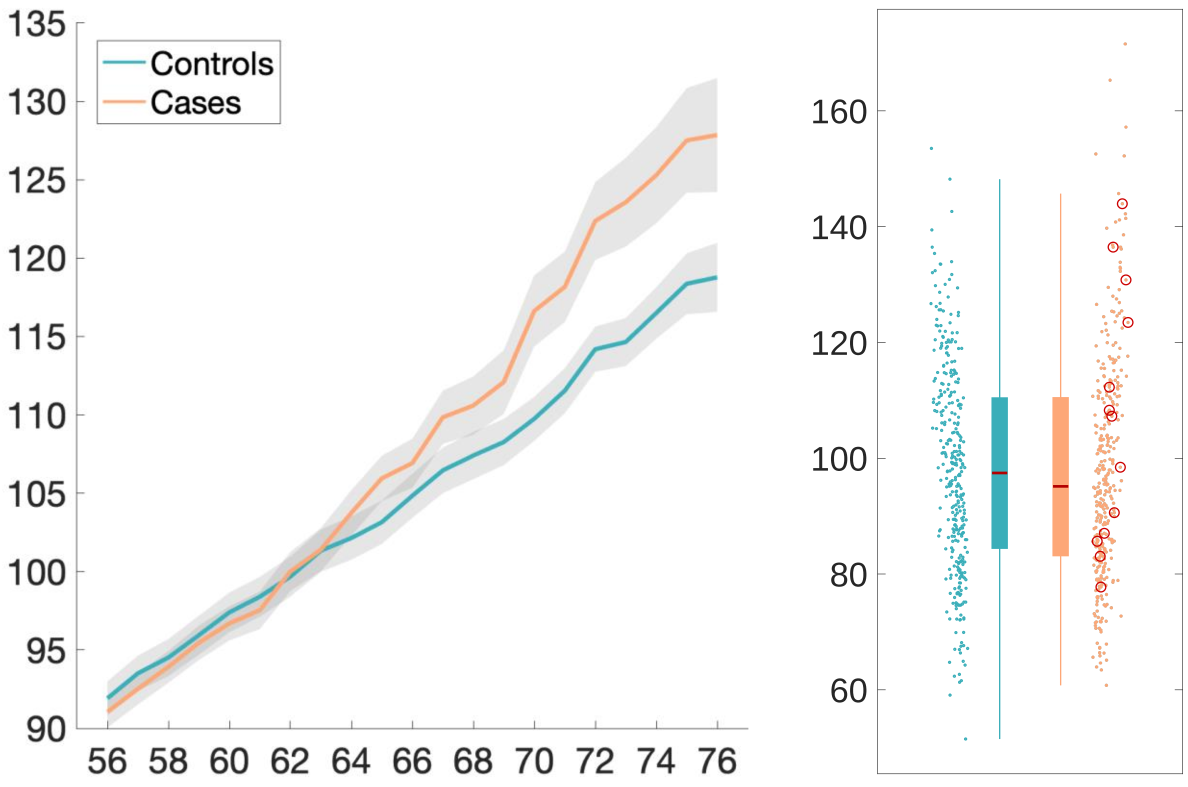

Anterior olfactory nucleus functional network – MD

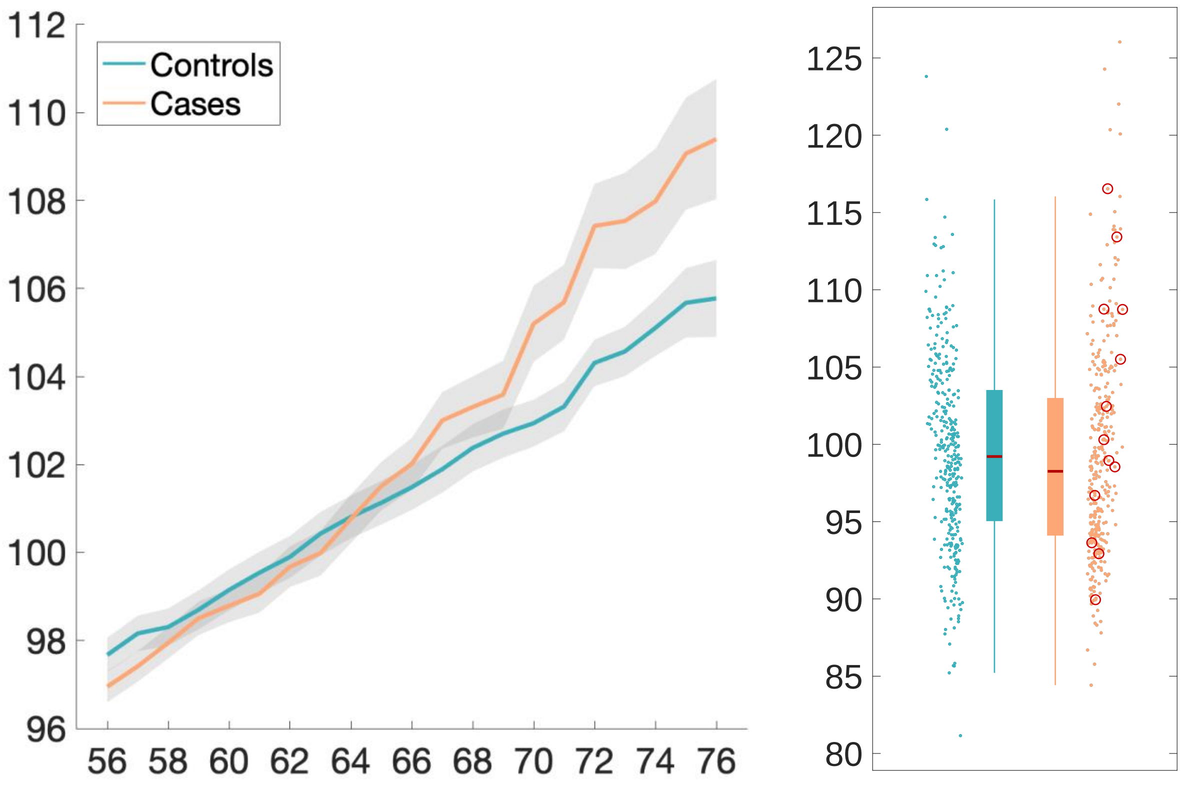

Parahippocampal gyrus L – intensity contrast (Desikan atlas)

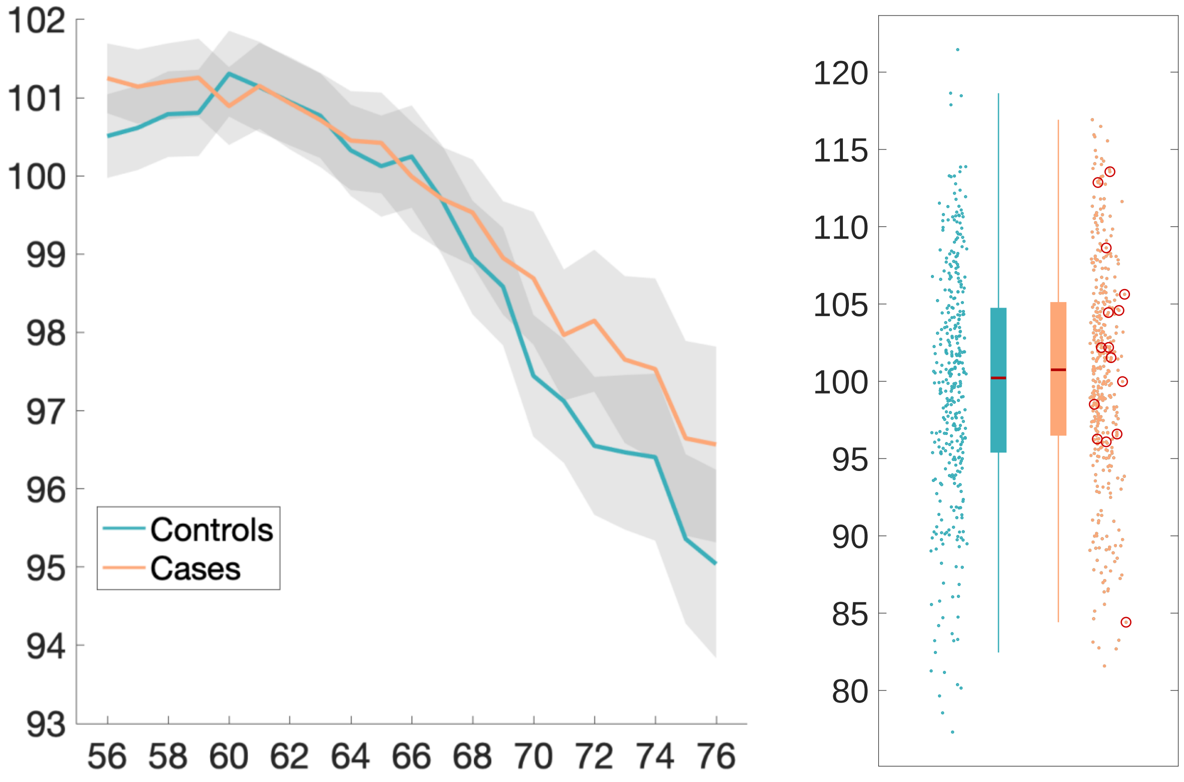

Anterior olfactory nucleus functional network – ISOVF

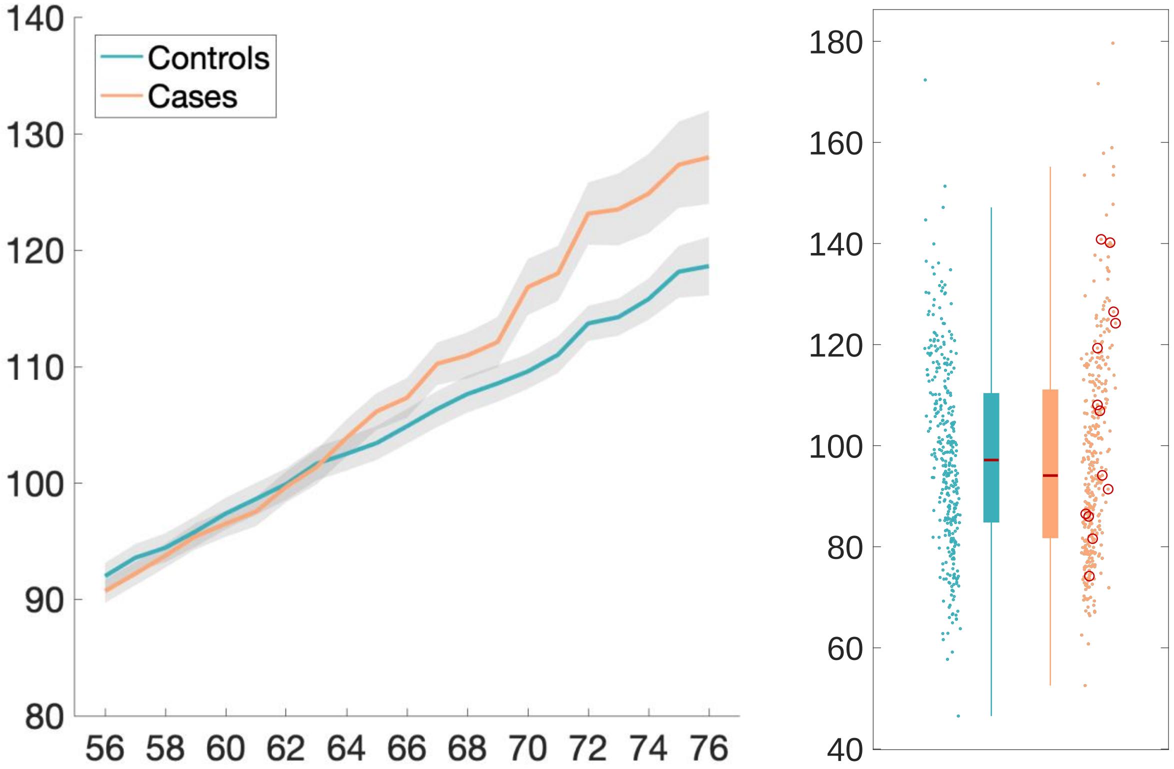

**Baseline exploratory results**

Ratio brain volume/estimated total intracranial volume

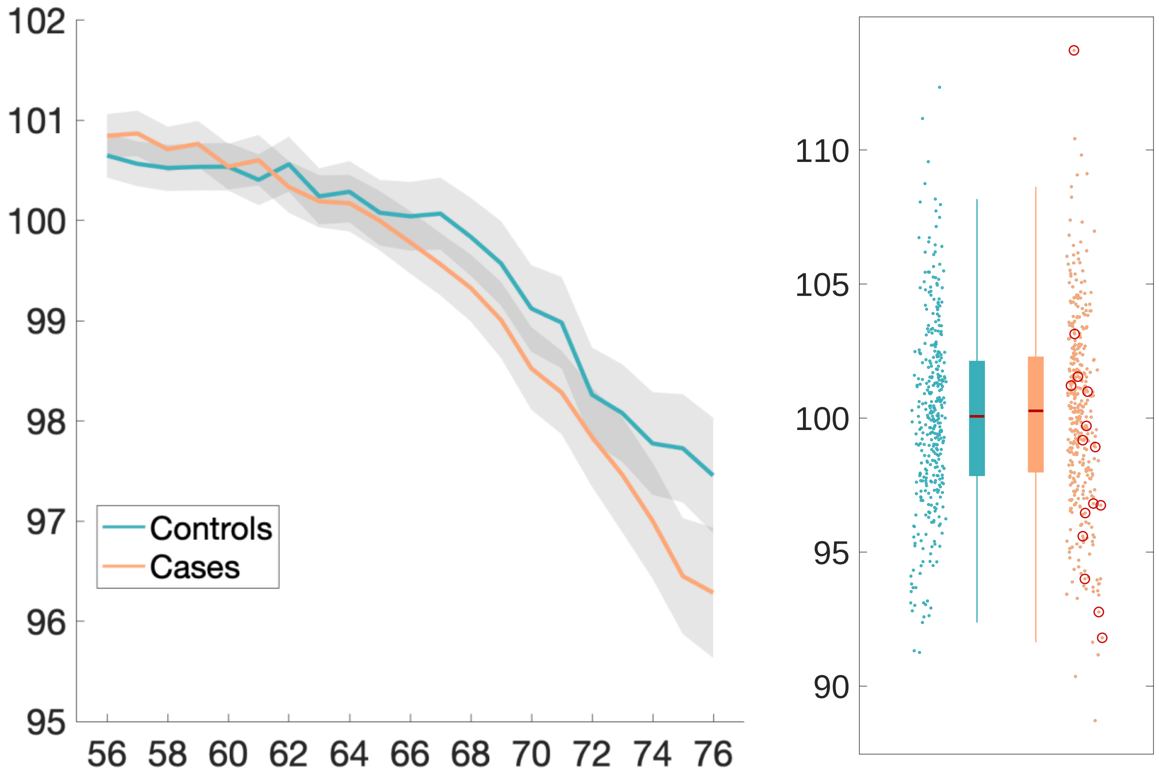

Normalised CSF – volume

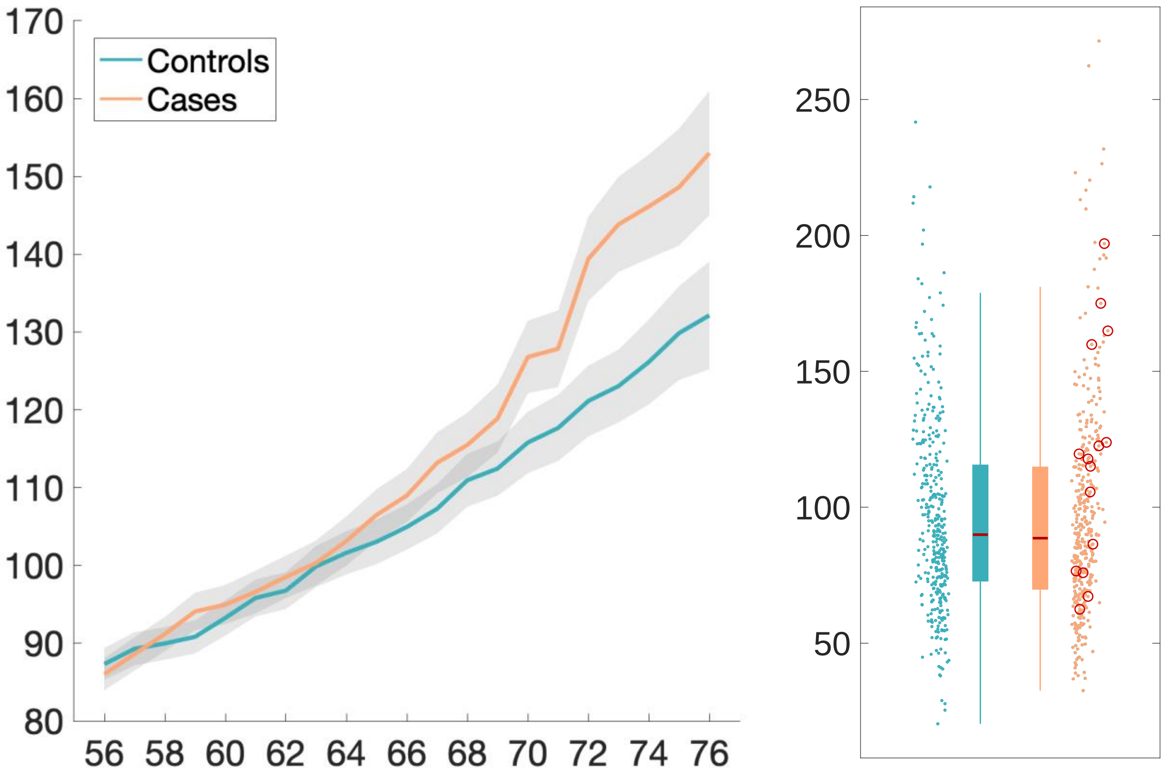

Lateral ventricle R – volume

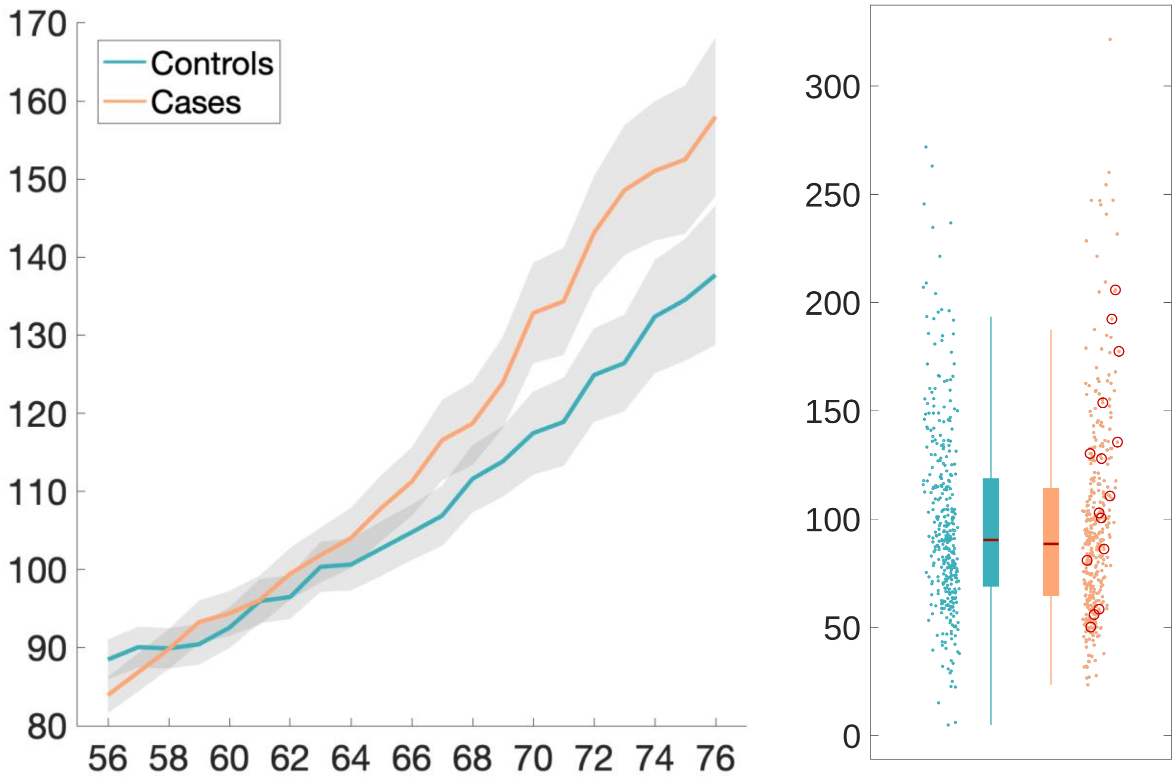

Superior fronto-occipital fasciculus R – ICVF

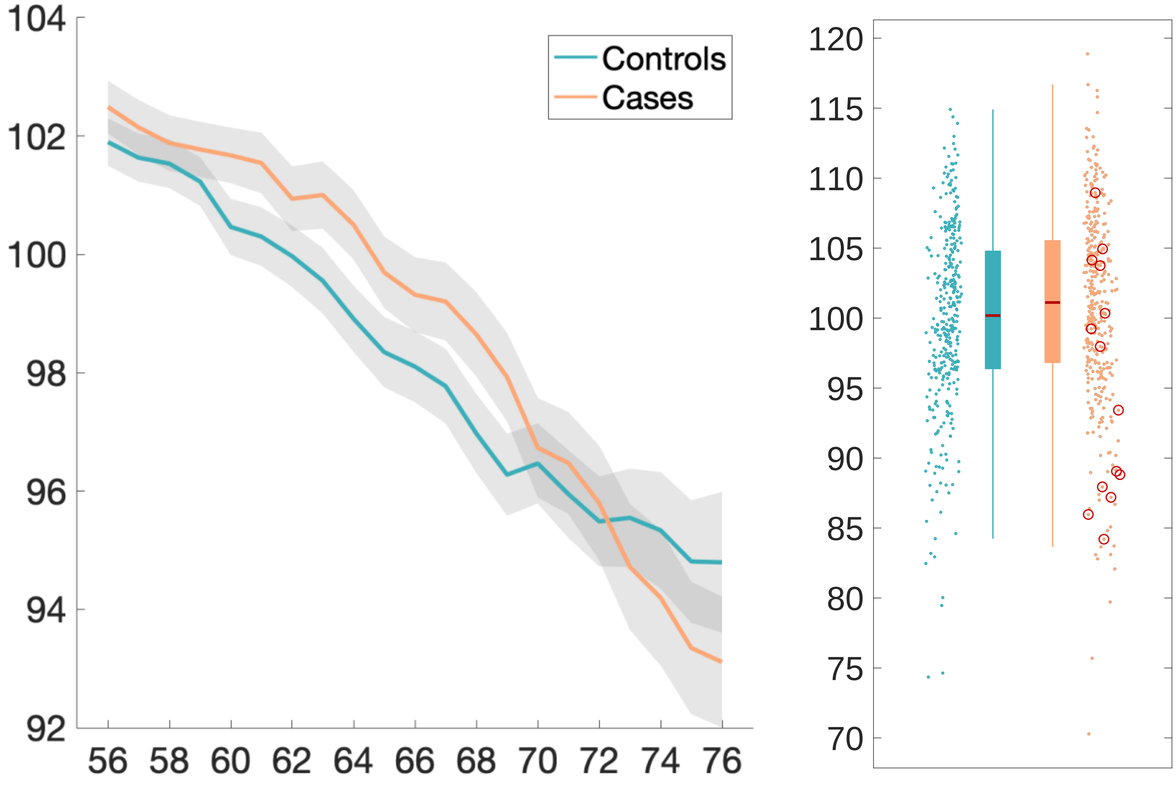

Brain volume without ventricles (surface model estimate)

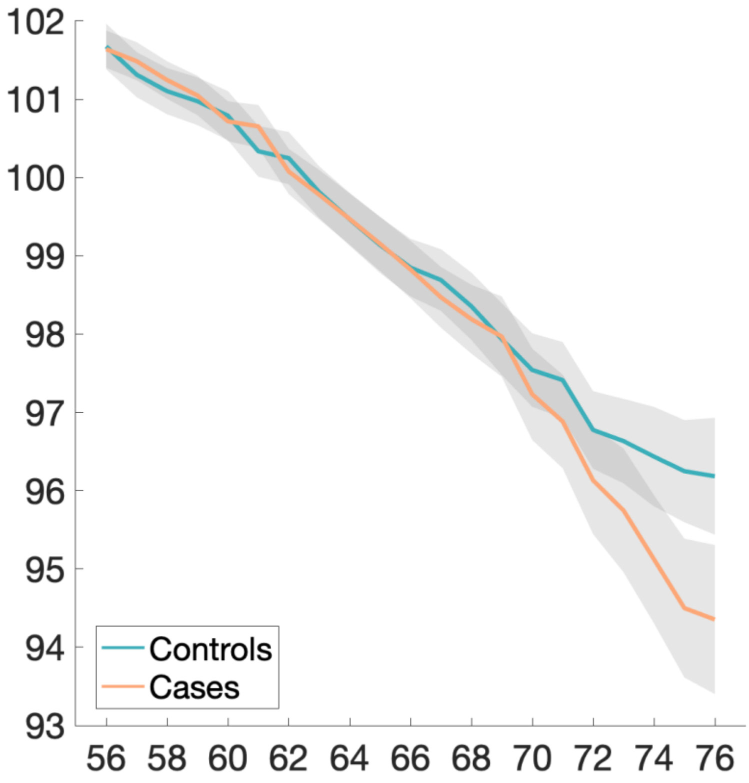


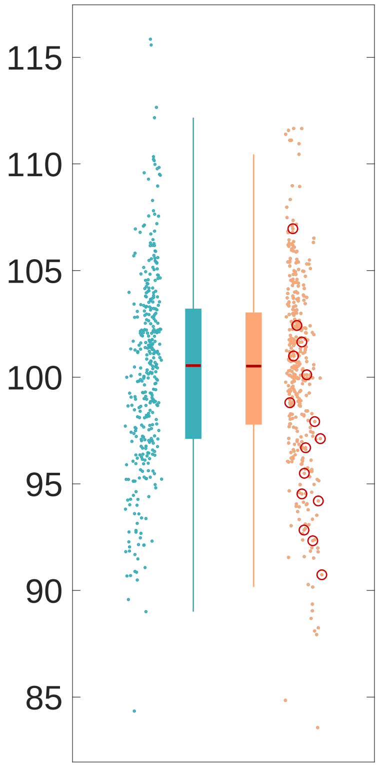

Rostral anterior cingulate cortex L – thickness (Desikan atlas)

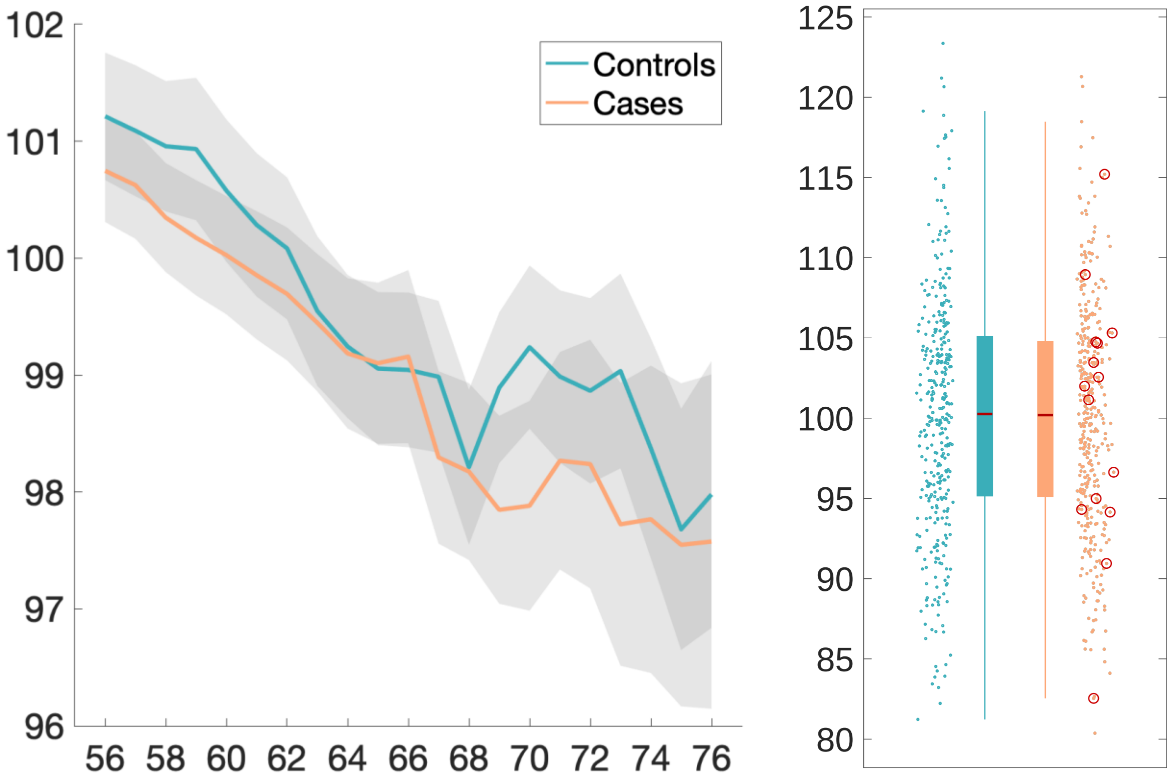

Brain volume without ventricles

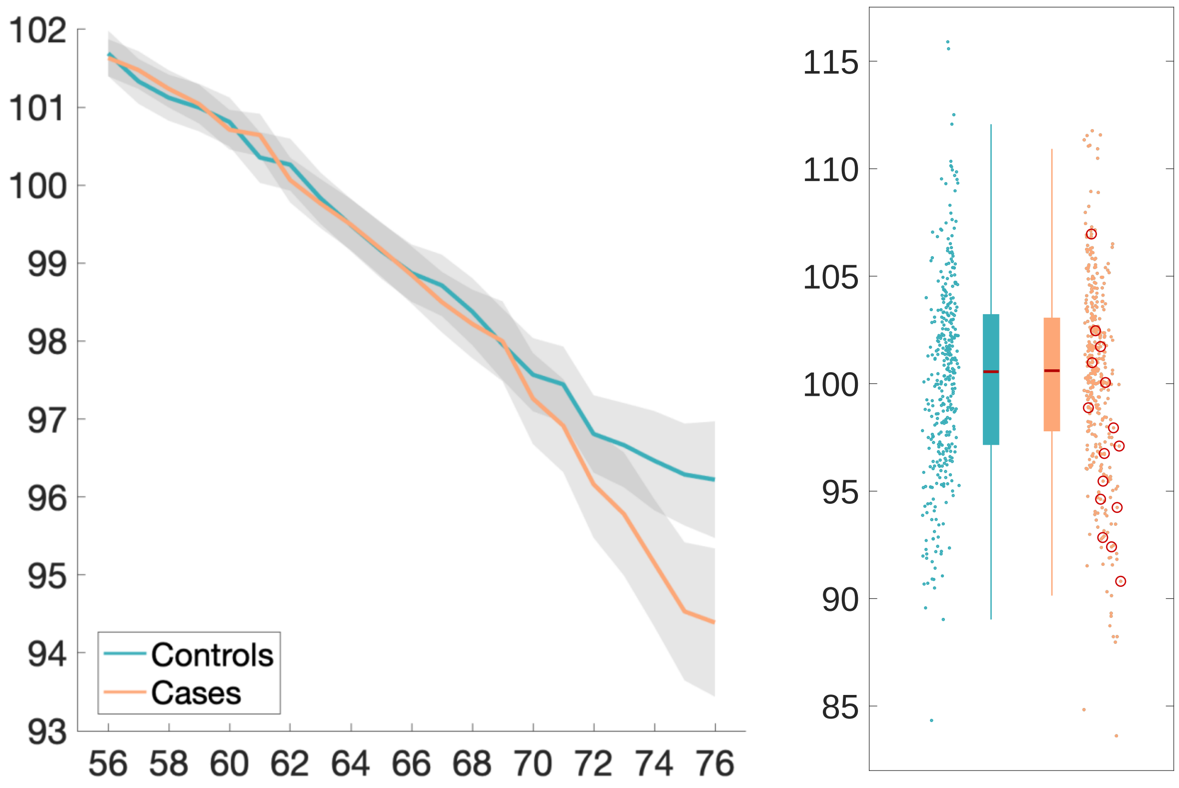

Supratentorial volume without ventricles

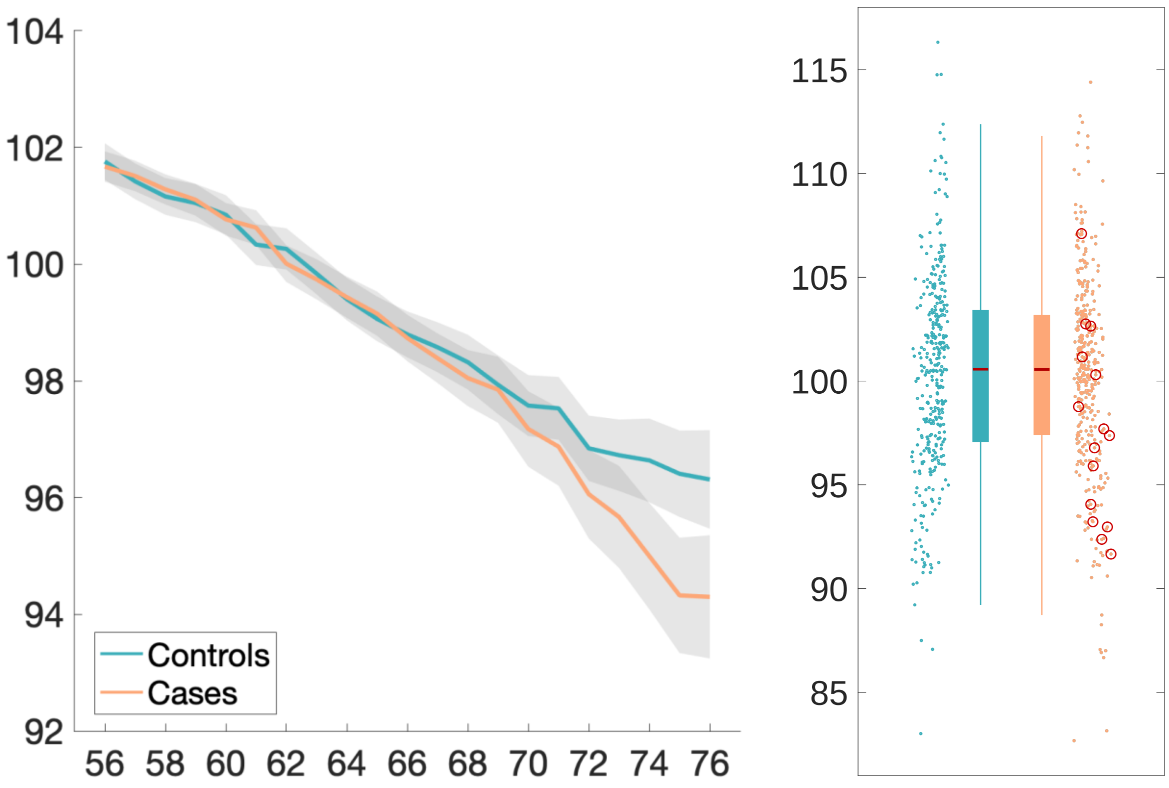

Cerebellum crus II – volume

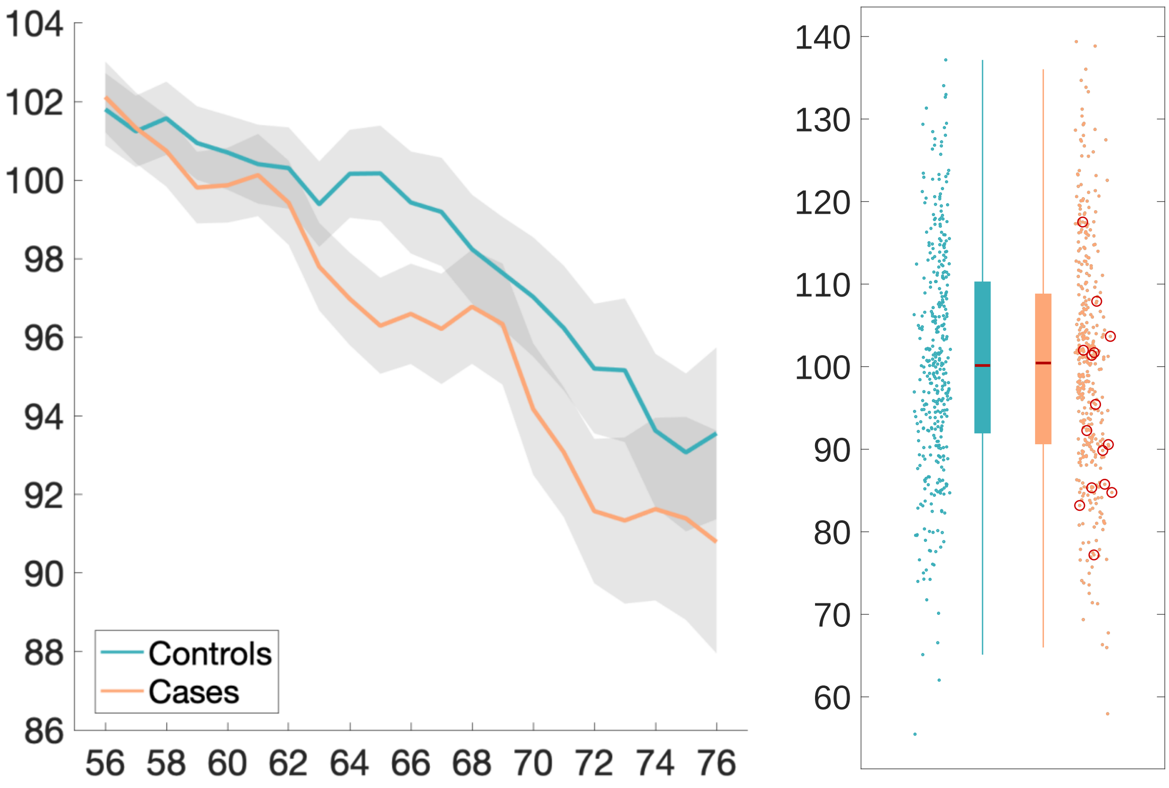

### Supplementary Analyses

#### Supplementary Analysis 1. Are nIDPs group-different at baseline, when clustering nIDPs using PCA?

#### Summary

When testing for case-vs-control differences in nIDPs at baseline, to reduce the sensitivity loss associated with multiple comparison correction across thousands of nIDPs, we first reduced the set of 6,301 nIDPs with PCA, testing a wide range of dimensionalities (1 to 700). No PCA components showed significant baseline effects. From 10 cognitive-only PCA components, 2 were significant.

#### Methods

We applied the same PCA-based reduction of the 6,301 nIDPs as described in Supplemental Analysis 1. We reduced the set of nIDPs to the top *Dmax* components, from which the top *D*<=*Dmax* can be then considered. Setting *Dmax* high (700, from a maximum rank of 785), allows for investigation of the eigenspectrum, which has a knee around *D*=10 (see **Supplementary Figure 5** below). We therefore applied the below evaluations with both *Dmax*=10 and (separately) *Dmax*=700. For all *D* from 1 to *Dmax*, we tested all components for between-group baseline difference, applying false discovery rate (FDR) and family-wise error (FWE) across components for a given *D* (while also considering the QQ plot to show potential uncorrected-P divergence from the null). To increase the chance of finding any between-group differences at baseline, no multiple comparison correction was applied across different dimensionalities *D* and different *Dmax*. We also reduced cognitive-only variables as described above, with *Dmax*=10 and *D* from 1 to 10.

#### Results

No nIDP-derived PCA components pass FDR or FWE at any *D*, for either *Dmax* (10 or 700). From the maximal set of tests, *Dmax*=700, D=700, the proportion of uncorrected P-values under 5% is 4.86%, consistent with the null; consistent with this, **Supplementary Figure 5** below shows the respective QQ plots for these uncorrected p-values. Also, when using a binary (non-age-modulated) two-group regressor and rerunning these tests, nothing passes FDR or FWE correction.

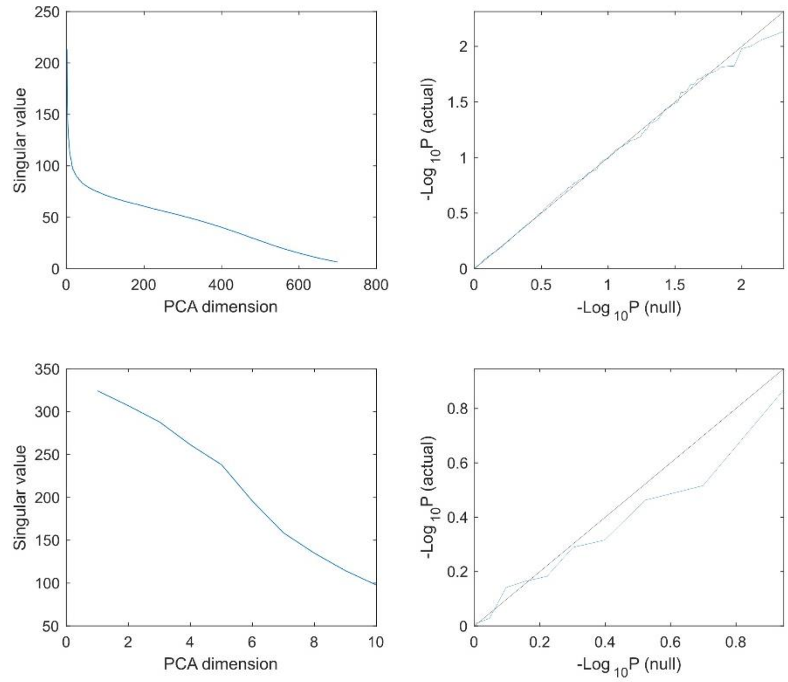


**Supplementary Figure 5. PCA eigenspectra and QQ Plots for $P_{uncorrected}$ against the theoretical null distribution**. Top: *Dmax*=700. Bottom: *Dmax*=10. Left: Eigenspectra from PCA with missing-data soft-imputation using an internal dimensionality of *Dmax*. Right: QQ plots of sorted uncorrected P-values, consistent with the null.

When applying PCA to the cognitive-only nIDPs, and testing for a group-difference at baseline, two PCs passed FDR thresholding. Within the PC, one cannot do a separate null-hypothesis statistical test (as the PCA is data-derived, and this would be circular), but we can report the nIDPs with the strongest weights in the PCA eigenvector. We thus normalised each PCA weight vector by subtracting the median and then normalising by the median absolute value, and report below the nIDPs with absolute normalised weight value greater than 5 for those two PCs.

**PCA, 10 components, uncorrected P-values:**
*0.0028* 0.6780 0.4811 0.*0039* 0.1830 0.1133 0.0981 0.2781 0.1456 0.9315
FDR threshold: 0.0039.

**nIDP PC weights (first column), along with UK Biobank variable codes and names.**

**PC 1**
-5.02 20016-1.0 Fluid intelligence score (1.0)
7.59 20156-0.0 Duration to complete numeric path (trail #1) (0.0)
7.67 20157-0.0 Duration to complete alphanumeric path (trail #2) (0.0)
-7.10 20159-0.0 Number of symbol digit matches made correctly (0.0)
-7.31 20195-0.0 Number of symbol digit matches attempted (0.0)

**PC 4**

5.42 4254-2.2 Time first key touched (2.2)
5.93 4254-2.3 Time first key touched (2.3)
5.48 4254-2.4 Time first key touched (2.4)
5.35 4255-2.2 Time last key touched (2.2)
5.91 4255-2.4 Time last key touched (2.4)
5.91 4255-2.7 Time last key touched (2.7)
6.32 4255-2.9 Time last key touched (2.9)
5.55 4256-2.2 Time elapsed (2.2)
6.10 4256-2.4 Time elapsed (2.4)
5.01 4256-2.6 Time elapsed (2.6)
6.80 4256-2.7 Time elapsed (2.7)
6.28 4256-2.9 Time elapsed (2.9)
5.45 4285-2.0 Time to complete test (2.0)
5.19 4291-1.0 Number of attempts (1.0)
5.78 20198-0.24 Test array presented (0.24)
5.82 20200-0.9 Values wanted (0.9)
5.69 20229-0.9 Values entered (0.9)

**Supplementary Analysis 2. Cognitive tests sensitive to cognitive impairment in people at risk for dementia**

**Summary**

We selected the top 10 cognitive variables that were the most sensitive to differences between 778 out-of-sample UK Biobank participants at risk of developing dementia and 778 out-of-sample matched controls.

**Methods**

We first identified all dementia cases and their diagnosis dates based on the most recently updated UK Biobank hospital inpatient records, GP records, and self-reports. Of the dementia cases identified, we selected a subgroup of 778 subjects who undertook a comprehensive set of cognitive tests, most *before* they were diagnosed for dementia: both the cognitive function tests at their initial assessment centre visit (2006-2010) and the online assessment (2014-2015). The types of cognitive function tests taken by the subjects are shown in the table below:

Initial assessment centre visit (2006-2010):

| **Cognitive Function Test** | **UKB Category ID** |
|---|---|
| Reaction time | 100032 |
| Numeric memory | 100029 |
| Fluid intelligence | 100027 |
| Prospective memory | 100031 |
| Pairs matching | 100030 |

Online (2014-2015):

| **Cognitive Function Test** | **UKB Category ID** |
|---|---|
| Numeric memory | 120 |
| Fluid intelligence | 118 |
| Trail making | 121 |
| Symbol digit substitution | 122 |
| Pairs matching | 117 |

At the time of initial assessment centre visit (2006-2010) ("Visit 0"), 15 (1.9%) out of the 778 subjects had been diagnosed with dementia. At the time of online cognitive tests, 83 (10.7%) out of the 778 subjects had been diagnosed with dementia. All the remaining cases (89.3%) were diagnosed *after* their cognitive testing. The median time difference between dementia diagnosis and cognitive tests was 6.6 years, i.e., there was a span of more than 6 years and a half between the participants undergoing their cognitive testing and their future diagnosis.

We then matched these participants with current or future dementia diagnoses with controls (**Supplementary Table 6**).

**Supplementary Table 6. Main demographics of the dementia cases and controls.** Non-parametric tests were used whenever a variable for each group was not normally distributed (Lilliefors P < 0.05). Two-sample Kolmogorov-Smirnov test was used for Age when the cognitive tests were taken, Years between diagnosis and cognitive test dates, Alcohol intake frequency, and Past tobacco smoking. Chi-square test was used for Sex, Ethnicity, and Diagnosed diabetes. Mann-Whitney U-test was used for the Systolic and Diastolic blood pressures, Weight, Waist/Hip ratio, BMI, Education, and Townsend deprivation index.

| | **Dementia** | **Controls** | **P** |
|---|---|---|---|
| **Number of subjects** | 778 | 778 | - |
| **Age at cognitive test, mean±SD (range)** | 64.0 ± 4.9 (42.2–70.3) | 63.9 ± 4.9 (42.0–70.2) | 1.00 |
| **Age at online cognitive test, mean±SD (range)** | 69.8 ± 4.9 (48.5–77.5) | 69.8 ± 4.9 (48.8–77.8) | 1.00 |
| **Sex, male/female** | 429 (55.1%) / 349 (44.9%) | 429 (55.1%) / 349 (44.9%) | 1.00 |
| **Ethnicity, white/non-white*** | 775 (97.0%) / 23 (3.0%) | 775 (97.0%) / 23 (3.0%) | 1.00 |
| **Systolic blood pressure [mmHg]** | 141.3 ± 18.4 | 141.2 ± 15.2 | 0.61 |
| **Diastolic blood pressure [mmHg]**** | 81.0 ± 10.0 | 81.7 ± 8.2 | 0.08 |
| **Diagnosed diabetes** | 69 (8.9%) | 69 (8.9%) | 1.00 |
| **Weight [kg]** | 77.1 ± 15.1 | 77.2 ± 13.9 | 0.63 |
| **Waist/Hip ratio** | 0.88 ± 0.09 | 0.88 ± 0.08 | 0.45 |
| **BMI [kg/m$^2$]** | 26.9 ± 4.5 | 26.7 ± 3.9 | 0.81 |
| **Alcohol intake frequency**** | 3.2 ± 1.6 | 3.4 ± 1.5 | 0.06 |
| **Tobacco smoking** | 0.88 ± 0.96 | 0.79 ± 0.94 | 0.19 |
| **Education** | 2.9 ± 1.2 | 2.8 ± 1.2 | 0.74 |
| **Townsend deprivation index** | -1.6 ± 2.9 | -2.0 ± 2.4 | 0.12 |

*The white/non-white distinction was made as numbers were unfortunately too low to allow for a finer distinction

**The controls show a trend towards consuming more alcohol, and having slightly higher diastolic blood pressure (rather than the other way around)

### Results

We found some significant differences across the 514 cognitive variables explored, and ranked them according to their uncorrected P values. When the same cognitive score for multiple visits was available, we selected the one with the lowest uncorrected P values. The top 10 cognitive variables are presented in **Supplementary Table 7**.

**Supplementary Table 7. Most different cognitive variables between 778 participants at risk for dementia and 778 matched controls.**

| Puncorr | N | Cognitive variable | Main test |
|---|---|---|---|
| 1.10E-34 | 1509 | Number of symbol digit matches made correctly (0.0) | Symbol digit substitution (online) |
| 3.10E-25 | 1212 | Duration to complete alphanumeric path (trail #2) (0.0) | Trail making (online) |
| 1.12E-21 | 1544 | Fluid intelligence score (0.0) | Fluid intelligence (online) |
| 1.43E-08 | 1212 | Duration to complete numeric path (trail #1) (0.0) | Trail making (online) |
| 3.32E-05 | 1546 | Time to complete round (0.1) | Pairs matching |
| 8.71E-05 | 1546 | Number of incorrect matches in round (0.1) | Pairs matching |
| 0.000815 | 639 | Total errors traversing alphanumeric path (trail #2) (0.0) | Trail making (online) |
| 0.003801 | 1506 | Number of correct matches in round (0.1) | Pairs matching (online) |
| 0.004171 | 1547 | Mean time to correctly identify matches (0.0) | Reaction time |
| 0.007522 | 176 | Maximum digits remembered correctly (0.0) | Numeric memory |

### Supplementary Analysis 3. Does pneumonia show pre-post IDP changes?

#### Summary

Analogous to our main analysis contrasting SARS-CoV-2 cases and controls, we identified 11 UK Biobank participants who are recorded as having contracted pneumonia not related to COVID-19 between their two imaging scans. Comparing these against 261 well-matched controls, we found some significant effects associated with pneumonia, but none of those overlapped with the main SARS-CoV-2 results.

#### Methods

We identified 11 UK Biobank participants who have a record of pneumonia not related to COVID-19 occurring between their two imaging visits. Four of these participants were hospitalised due to pneumonia; the others were identified on the basis of GP records (n=5) and self-reported illness conditions (n=2). We identified a set of 261 matched controls (see **Supplementary Table 8** below) who have had two imaging visits. We then carried out identical group-difference IDP-change analyses to those we carried out for the SARS-CoV-2 cases vs controls.

**Supplementary Table 8**. **Main demographics of the pneumonia patients and controls.** Non-parametric tests were used whenever a variable for each group was not normally distributed (Lilliefors P < 0.05). Two-sample Kolmogorov-Smirnov test was used for Age at Scan 1 or Scan 2, Years between Scans 1 and 2, Alcohol intake frequency, and Past tobacco smoking. Chi-square test was used for Sex, Ethnicity, and Diagnosed diabetes. Mann-Whitney U-test was used for the Systolic and Diastolic blood pressures, Weight, Waist/Hip ratio, BMI, and Townsend deprivation index.

| | **Pneumonia patients** | **Controls** | **P** |
|---|---|---|---|
| **Number of subjects** | 11 | 261 | - |
| **Age at Scan 1, mean$\pm$SD (range)** | 68.6 $\pm$ 6.6 (57.2–79.1) | 67.6 $\pm$ 5.1 (56.4–79.5) | 0.78 |
| **Age at Scan 2, mean$\pm$SD (range)** | 70.8 $\pm$ 6.7 (59.5–81.4) | 69.9 $\pm$ 5.1 (58.6–81.6) | 0.83 |
| **Sex, male/female** | 6 (54.5%) / 5 (45.5%) | 120 (46%) / 141 (54.0%) | 0.58 |
| **Ethnicity, white/non-white*** | 11 (100%) / 0 (0%) | 261 (100%) / 0 (0%) | 1.00 |
| **Years between Scans 1 and 2, mean$\pm$SD (range)** | 2.3 $\pm$ 0.1 (2.1–2.5) | 2.3 $\pm$ 0.1 (2.0–2.9) | 0.25 |
| **Systolic blood pressure [mmHg]** | 134.7 $\pm$ 11.9 | 134.9 $\pm$ 16.6 | 0.96 |
| **Diastolic blood pressure [mmHg]** | 78.1 $\pm$ 8.5 | 76.9 $\pm$ 10.5 | 0.65 |
| **Diagnosed diabetes** | 1 (9.1%) | 12 (4.6%) | 0.49 |
| **Weight [kg]** | 73.3 $\pm$ 14.8 | 73.0 $\pm$ 13.9 | 0.82 |
| **Waist/Hip ratio** | 0.74 $\pm$ 0.14 | 0.73 $\pm$ 0.11 | 0.82 |
| **BMI [kg/m$^2$]** | 26.0 $\pm$ 4.0 | 26.3 $\pm$ 4.2 | 0.85 |
| **Alcohol intake frequency** | 3.2 $\pm$ 1.3 | 3.2 $\pm$ 1.4 | 1.00 |
| **Tobacco smoking** | 0.82 $\pm$ 0.98 | 0.72 $\pm$ 0.94 | 1.00 |
| **Townsend deprivation index** | -3.0 $\pm$ 1.6 | -2.2 $\pm$ 2.6 | 0.54 |

*The white/non-white distinction was made as numbers were unfortunately too low to allow for a finer distinction

**Results**

With the full ("Exploratory") sets of IDPs, some FDR-significant differences between pneumonia cases and controls were found in the longitudinal IDP change of: the total white matter volume, cerebellar volume of lobe V, white matter lesions, and fractional anisotropy changes in the corona radiata (**Supplementary Table 9**). These results do not overlap with the main longitudinal IDP differences found with SARS-CoV-2 (**Tables 4 and 5**). Asking a similar question, but more liberally (i.e., looking at weaker associations than those passing FDR), no IDPs have |Z|>3 for both pneumonia and SARS-CoV-2. Correlation between all IDPs' Z-statistics from pneumonia and SARS-CoV-2 is very low (r=0.057).

No significant pneumonia associations were found when restricting the IDPs to the hypothesis-driven IDP set used in the main SARS-CoV-2 analyses (minimum $P_{uncorrected}$ = 0.005, no FDR significant associations).

**Supplementary Table 9. Significant longitudinal group comparison results between pneumonia cases and controls.** Significant results, surviving false discovery rate (FDR) correction (FDR threshold was 0.00017), showing where the 11 pneumonia cases and 261 controls differed over time. We report % change between the two groups, standard error (SE) on these % changes, and uncorrected and family-wise error (FWE) corrected P values. Note: the Z-statistics reflect the statistical strength of the longitudinal group-difference modelling, and are not raw data effect sizes. L is left.

| % | SE | z | p_raw | fdr | p_corr | fwe | n | IDP |
|---|---|---|---|---|---|---|---|---|
| 2.96 | 0.63 | 4.6 | 0.000004 | * | 0.0472 | * | 269 | IDP_T1_SIENAX_white_unnormalised_volume |
| 2.89 | 0.62 | 4.5 | 0.000006 | * | 0.0639 | | 269 | IDP_T1_SIENAX_white_normalised_volume |
| -1.34 | 0.32 | -4.1 | 0.000037 | * | 0.2029 | | 265 | IDP_dMRI_TBSS_FA_Superior_corona_radiata_L |
| -4.28 | 1.07 | -3.9 | 0.000080 | * | 0.3041 | | 272 | IDP_T1_FAST_ROIs_L_cerebellum_V |
| -1.42 | 0.36 | -3.9 | 0.000100 | * | 0.3454 | | 266 | IDP_dMRI_TBSS_FA_Posterior_corona_radiata_R |
| 53.18 | 13.58 | 3.8 | 0.000121 | * | 0.3766 | | 226 | IDP_T2_FLAIR_BIANCA_deepWMH_volume |
| -1.36 | 0.35 | -3.8 | 0.000133 | * | 0.3965 | | 265 | IDP_dMRI_TBSS_FA_Anterior_corona_radiata_R |
| -4.03 | 1.05 | -3.8 | 0.000156 | * | 0.4255 | | 272 | IDP_T1_FAST_ROIs_R_cerebellum_V |

**Supplementary Analysis 4. Does influenza show pre-post IDP changes?**

As carried out for pneumonia, we also tested influenza in the UK Biobank. We identified 5 cases (3 hospitalised) who had contracted influenza (either self-reported, or reported in hospital or primary care records) between their two scans and 127 matched controls (**Supplementary Table 10**), finding no significant effects, in the main two-group test. Correlation of Z-statistics between influenza and SARS-CoV-2 was low (r=0.077). There were no common results where any IDPs had Z>3 or Z<-3 for both conditions. For the hypothesis-driven set of IDPs, there were no FDR or FWE significant results. A few significant results in the 3 hospitalised cases vs controls were found, with one IDP (volume of the brainstem) overlapping with our main longitudinal results in SARS-CoV-2 (**Supplementary Table 11**).

**Supplementary Table 10. Main demographics of the influenza cases and controls.** Non-parametric tests were used whenever a variable for each group was not normally distributed (Lilliefors P < 0.05). Chi-square test was used for Sex, Ethnicity, and Diagnosed diabetes. Mann-Whitney U-test was used for all other variables.

| | **Influenza patients** | **Controls** | **P** |
|---|---|---|---|
| **Number of subjects** | 5 | 127 | - |
| **Age at Scan 1, mean±SD (range)** | 65.3 ± 8.7 (52.4–75.5) | 65.6 ± 5.6 (52.1–75.5) | 0.89 |
| **Age at Scan 2, mean±SD (range)** | 67.4 ± 8.7 (54.6–77.5) | 67.9 ± 5.6 (54.4–77.9) | 0.81 |
| **Sex, male/female** | 2 (40%) / 3 (60%) | 39 (30.7%) / 88 (69.3%) | 0.66 |
| **Ethnicity, white/non-white*** | 5 (100%) / 0 (0%) | 127 (100%) / 0 (0%) | 1.00 |
| **Years between Scans 1 and 2, mean±SD (range)** | 2.2 ± 0.1 (2.0–2.4) | 2.2 ± 0.1 (2.0–2.7) | 0.25 |
| **Systolic blood pressure [mmHg]** | 138.4 ± 14.6 | 132.9 ± 16.1 | 0.35 |
| **Diastolic blood pressure [mmHg]** | 82.0 ± 8.9 | 76.7 ± 10.9 | 0.29 |
| **Diagnosed diabetes** | 0 (0%) | 6 (4.7%) | 0.62 |
| **Weight [kg]** | 75.8 ± 15.6 | 71.7 ± 13.7 | 0.53 |
| **Waist/Hip ratio** | 0.74 ± 0.13 | 0.70 ± 0.10 | 0.47 |
| **BMI [kg/m$^2$]** | 27.1 ± 4.9 | 26.6 ± 4.8 | 0.73 |
| **Alcohol intake frequency** | 2.8 ± 2.3 | 2.9 ± 1.5 | 0.98 |
| **Tobacco smoking** | 1.0 ± 1.4 | 0.60 ± 0.95 | 0.52 |
| **Townsend deprivation index** | -3.3 ± 1.5 | -1.8 ± 2.7 | 0.18 |

*The white/non-white distinction was made as numbers were unfortunately too low to allow for a finer distinction

**Supplementary Table 11. Significant longitudinal group comparison results between hospitalised influenza cases and controls.** A few significant results, only surviving false discovery rate (FDR) correction (FDR threshold was 0.00014), were found between the 3 hospitalised influenza cases and 127 controls. We report % change between the two groups, standard error (SE) on these % changes, and uncorrected and family-wise error (FWE) corrected P values. Note: the Z-statistics reflect the statistical strength of the longitudinal group-difference modelling, and are not raw data effect sizes. R is right.

| % | SE | z | p_raw | fdr | p_corr | n | IDP |
|---|---|---|---|---|---|---|---|
| 3.44 | 0.81 | 4.1 | 0.000046 | * | 0.3975 | 124 | aseg_global_volume_Brain-Stem |
| -11.21 | 2.58 | -4.2 | 0.000028 | * | 0.3231 | 126 | aseg_lh_volume_Cerebellum-White-Matter |
| 4.48 | 1.09 | 4 | 0.000073 | * | 0.4767 | 125 | aseg_lh_volume_Cerebellum-Cortex |
| -4.66 | 1.13 | -4 | 0.000063 | * | 0.4529 | 130 | IDP_dMRI_TBSS_FA_Inferior_cerebellar_peduncle_R |
| -3.06 | 0.75 | -3.9 | 0.000081 | * | 0.498 | 130 | IDP_dMRI_TBSS_ICVF_Retrolenticular_part_of_ internal_capsule_R |
| 27.52 | 6.79 | 3.9 | 0.000089 | * | 0.5142 | 130 | rfMRI amplitudes (ICA25 node 5) |

**Supplementary Analysis 5. The form of the case-control group-difference regressor**

The age-modulation factor (see main **Methods**) accounts for more pronounced effects in older people. We chose to focus on an "objective" age model given the strong prior knowledge of a highly increased detrimental effect, at older ages, of SARS-CoV-2 infection, and a greater vulnerability of the brain with age. We therefore used an already-published data-driven curve of age-effect in SARS-CoV-2 infection[2], with no free or subjectively-chosen parameters—hence having the same degrees-of-freedom fitting power as a binary model. This dependence is $10^{age\times0.0524-3.27}$, where the constant 0.0524 has units years$^{-1}$ and determines the exact strength of the age dependence, while the constant 3.27 results in a fixed multiplicative factor which therefore has no effect on our modelling.

We have recently identified a separate set of data for age-dependent mortality rates, from the UK Office of National Statistics. This gives an almost identical exponential age dependence: males: 0.0451± 0.0008, females: 0.0475±0.0005, very close to the value of 0.0524 in the Levin study[2].

In another analysis, of hospitalisations (not mortality) due to COVID-19, the function of age has also been found to be exponential in form, with exponent dependence ~0.02[3]. If we use this age modulation instead, extremely similar results are found for the hypothesis-driven Z-statistics; correlating the Z-statistics between the two modellings gives corr(Z)=0.96, with the strongest original results (|Z|>3.5) *increasing* in strength by an average of Z=0.16.

We also tested—as a secondary analysis—the binary modelling between control and SARS-CoV-2 groups, with all other factors held the same. These two (age-modulated and non-modulated) models do not give very different primary results (hypothesis-driven results presented in **Table 1 and Supplementary Table 1**). The findings are highly similar, if a little weaker, consistent with our expectation of increased effects at higher ages. For instance, from the 10 most significant results reported in **Table 1**, 10 associations had passed FDR correction, and 6 passed FWE correction. When switching to the binary (non-modulated) group modelling, 9 out of 10 continue to pass FDR correction and 4 out of 6 pass FWE correction. The correlation between the Z-statistics from all 297 hypothesis-driven IDPs is high (r=0.88), and the mean change in |Z| across the 10 top associations was a reduction of just 0.33. Results obtained with the binary regressor in all four Models are shown in **Supplementary Table 5.**

#### Supplementary Analysis 6. Further model-fitting validity/robustness evaluations

We created diagnostic plots from the 19 IDPs obtained by combining the top 10 IDPs from the hypothesis-driven set, and the top 10 IDPs from the exploratory set (one IDP belongs to both sets)(**Supplementary Figure 6**). These show scatterplots of residuals (y axis) against model-predicted values (x axis), with one plot per IDP. There is no obvious evidence of model misspecification.

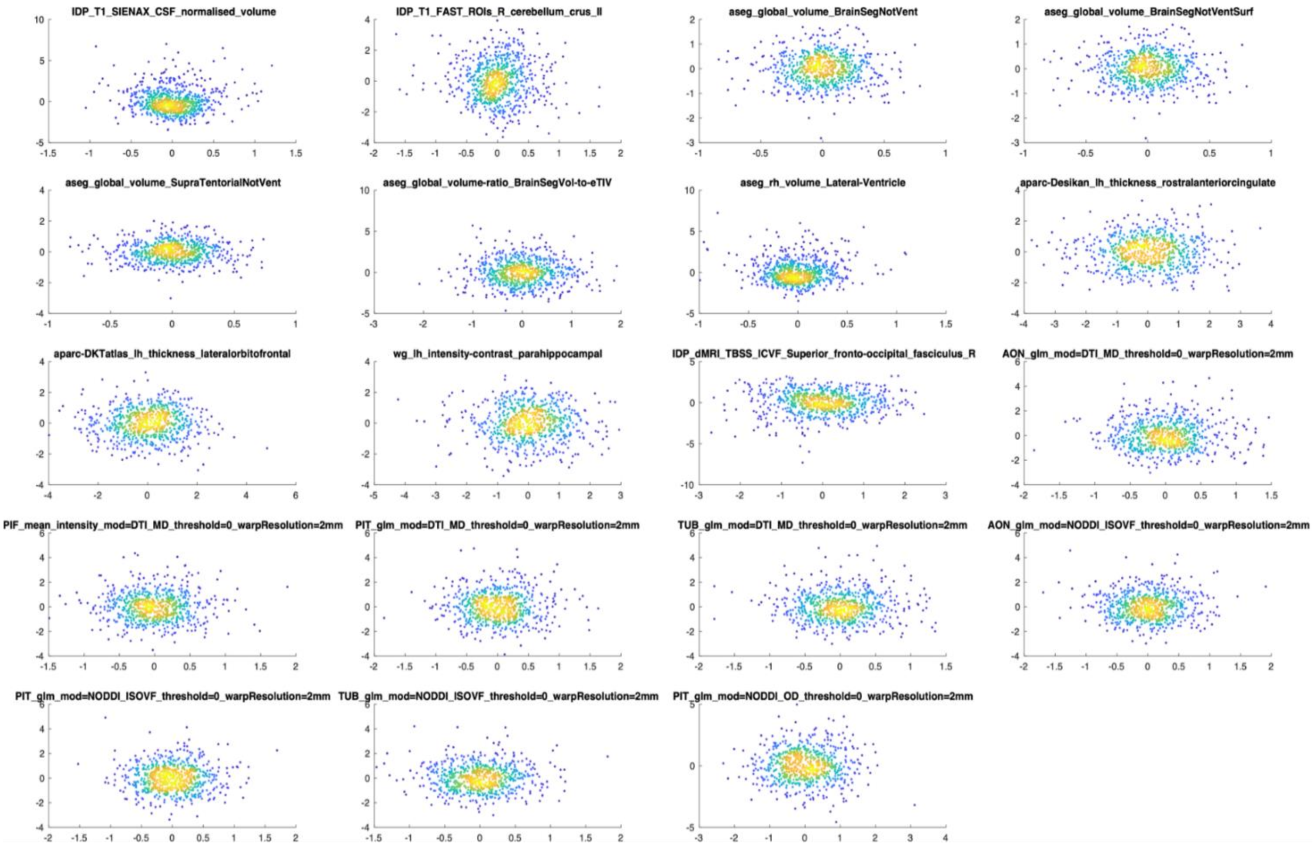


**Supplementary Figure 6. Scatterplots showing the model-fit IDP residuals (y axis) against model-predicted IDP values (x axis) for all top IDPs from both hypothesis-driven and exploratory approaches.**

Similarly, we created QQ plots from the model residuals for each of these top IDPs (**Supplementary Figure 7**). These show the ordered residuals (y axis) plotted against the standard normal quantiles, and again show no evidence of worrying violations of normality assumptions.

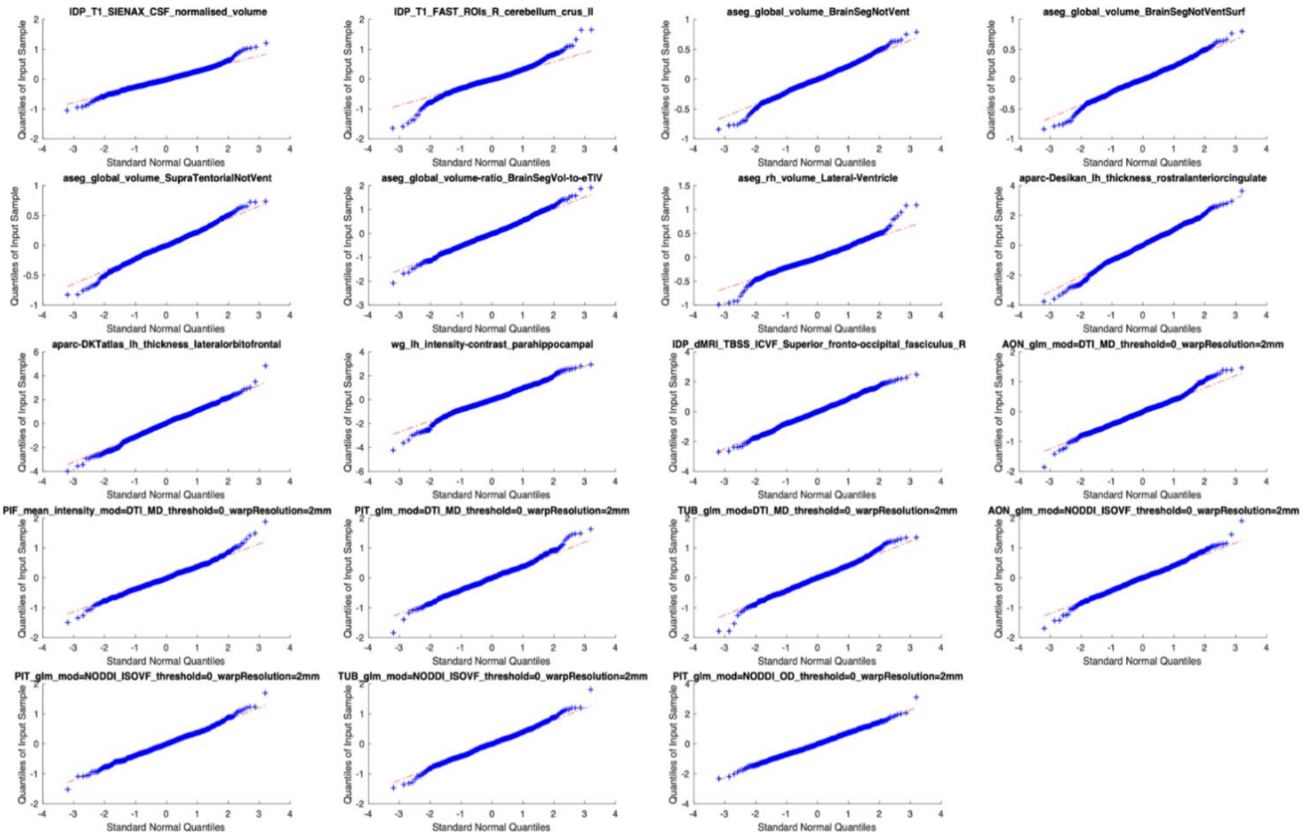


**Supplementary Figure 7. QQ Plots of model-fit IDP residuals for all top IDPs from both hypothesis-driven and exploratory approaches.**

Finally, we assessed sensitivity by evaluating model robustness, with respect to a set of additional higher-order potentially confounding effects. While the primary model considered as nuisance the main effects of age, sex, ethnicity and Townsend, there might be interactions among these variables that could alter the results. Therefore, we added an additional 15 confound variables corresponding to up-to 3-way interactions among age, age$^2$, sex, townsend and ethnicity:

age × sex, age$^2$ × sex, age × ethnicity, age$^2$ × ethnicity, age × townsend, age$^2$ × townsend,
sex × ethnicity, sex × townsend, ethnicity × townsend,
age × sex × ethnicity, age × sex × townsend, age × townsend × ethnicity,
age$^2$ × sex × ethnicity, age$^2$ × sex × townsend, age$^2$ × townsend × ethnicity.

We then reran the main case-vs-control model and compared Z-statistics, across the full ("exploratory") set of IDPs. The results are almost unchanged; comparing this Z against the original values, the correlation is r=0.98, the mean absolute difference is 0.19, and the mean absolute difference for the top 10 IDPs from Table 4 is 0.15.

**Supplementary Analysis 7. Do nIDPs explain the main (group-difference, longitudinal effect) results?**

**Summary**

We took 6,301 nIDPs (non-imaging-derived phenotypes and demographic measures), measured before, or at the time of, Scan1. We added them as confound variables in our main modelling, and found that none of these new confounds reduced any of the top main results (Z-statistics) by more than 25%. Of more potential interest, we also took the top 100 PCA components from these 6,301 nIDPs, and even when using all as 100 additional confounds in a single regression, found that none of the main results were reduced by more than 25%.

**Methods**

We considered all nIDPs that were measured at the time of, or before, Scan1. We excluded nIDPs having too little variability in their values, judged according to whether the largest group of subjects that had an identical value (to each other) was more than 97% of the total number of subjects with valid data. This resulted in 6,301 nIDPs, across all nIDP categories. We then considered, one at a time, each of the top-10 IDPs from the hypothesis-driven analyses, and the top-10 from the exploratory analyses (**Tables 4 and 5**), and for each, evaluated whether this set of 6,301 "baseline" nIDPs might reduce the strength of the main results when added as additional confounds into the longitudinal group-difference modelling.

Hence, for each imaging-derived phenotype (IDP), we initially evaluated nIDPs one at a time, considering nIDPs with <50% joint-missing-data (that is, subjects with data missing in nIDP or IDP). For each nIDP, we then estimated the (regression-based) association Z-statistic ("Z") for association of delta-IDP with the age-modulated group-difference regressor, as done for the main longitudinal analyses, but with the nIDP as an additional confound. To take into account the effect of missing data, we also estimated the association ("Z0"), without using the additional confound, but applying the same pattern of missing data. We then tested for Z/Z0<0.75 (first inverting the sign of both if Z0 was negative). This is therefore a reasonably aggressive test that only allows for 25% reduction in Z. If Z is reduced by more than this, we report the result as the nIDP potentially "explaining" the main result.

In addition, as a more powerful (aggressive) test, instead of applying nIDPs as confounds one at a time, we first estimated the top 100 components from a PCA (principal components analysis) across the 6,301 nIDPs, and then applied this entire matrix of potential confounds as a 100-variable confound matrix in the main longitudinal modelling for each of the IDPs. This therefore can potentially remove a lot more variance from the IDPs than when considering individual nIDPs, one at a time, as carried out above. Because of missing data in the nIDPs, we first removed nIDPs with more than ⅞ subjects missing, leaving 4,449, applied the same deconfounding as carried out in our main IDP modelling, normalised each nIDP, and then estimated the top 100 principal components using a soft imputation approach that iterates between imputing missing data using PCA bases, and re-estimating the PCA; this is implemented in FSLNets, and is similar to the approached described in[1]. Each IDP was then tested using the 100 additional confounds simultaneously, again testing for whether Z/Z0<0.75. We also tested each of the 100 PCA components on its own to see if Z was reduced for any IDPs by more than 25%.

Finally, we took the set of 516 nIDPs derived from cognitive testing carried out at the time of, or before, Scan1, and reduced these to the top 10 PCA components (in order to focus this variable-clustering to find latent variables from just this one, important, class of nIDPs, the cognitive scores). We then used these to test for any reductions >25% in Z for each IDP.

**Results**

None of these tests (all combinations of IDPs, nIDPs and PCA components derived from nIDPs) showed any cases of Z being reduced by more than 25%. Additionally, when using the binary (instead of age-modulated) group-difference regressor, again no reductions >25%were found. These evaluations are more directly relevant than separately asking whether baseline nIDPs correlated with IDPs, or are different between the groups; here we are in effect asking about both factors combined, which is the most direct test of whether baseline measures might account for (i.e., explain away) the main results.

### Supplementary Analyses References


1 Cai, J. F., Candes, E. J. & Shen, Z. W. A Singular Value Thresholding Algorithm for Matrix Completion. *Siam J Optimiz* **20**, 1956-1982, doi:10.1137/080738970 (2010).

2 Levin, A. T. *et al.* Assessing the age specificity of infection fatality rates for COVID-19: systematic review, meta-analysis, and public policy implications. *Eur J Epidemiol* **35**, 1123-1138, doi:10.1007/s10654-020-00698-1 (2020).

3 Palmer, S., Cunniffe, N. & Donnelly, R. COVID-19 hospitalization rates rise exponentially with age, inversely proportional to thymic T-cell production. *J R Soc Interface* **18**, 20200982, doi:10.1098/rsif.2020.0982 (2021).
